# Novel genetic loci in adolescent-onset gout derived from whole genome sequencing of a Chinese cohort

**DOI:** 10.1101/2023.04.18.23288731

**Authors:** Aichang Ji, Yang Sui, Xiaomei Xue, Xiaopeng Ji, Yongyong Shi, Robert Terkeltaub, Nicola Dalbeth, Riku Takei, Fei Yan, Mingshu Sun, Maichao Li, Jie Lu, Lingling Cui, Zhen Liu, Can Wang, Xinde Li, Lin Han, Zhanjie Fang, Wenyan Sun, Yue Liang, Yuwei He, Guangmin Zheng, Xuefeng Wang, Jiayi Wang, Hui Zhang, Lei Pang, Han Qi, Yushuang Li, Zan Cheng, Zhiqiang Li, Jingfa Xiao, Changqing Zeng, Tony R. Merriman, Hongzhu Qu, Xiangdong Fang, Changgui Li

## Abstract

**Background:** Gout is a polygenetic inflammatory disease. Although hundreds of genetic variants associated with gout and serum urate levels have been identified in studies of adults, the pathogenesis of adolescent-onset gout remains unclear. To better characterize the genetic landscape of adolescent-onset gout, a whole genome sequencing study was done in a large Chinese adolescent-onset gout cohort.

**Methods:** We conducted whole genome sequencing in a discovery adolescent-onset gout cohort of 905 individuals (gout onset 12-19 years) to discover common SNVs, uncommon SNVs, and indels associated with gout. Candidate common SNVs were replicated in an early-onset gout cohort of 2834 individuals (gout onset ≤ 30 years old). Loci associated with early-onset gout (*P* < 5.0 × 10^−8^) were identified after meta-analysis with the discovery and replication cohorts. Transcriptome and epigenomic analyses, RT-qPCR and RNA-seq in human peripheral blood leukocytes, and knock-down experiments in human THP-1 macrophage cells investigated regulation and functions of candidate gene *RCOR1*.

**Findings:** In addition to *ABCG2*, a urate transporter previously linked to pediatric-onset and early-onset gout, we identified four novel loci: *VPRBP* (rs868933181, *P*_meta_ = 6.27 × 10^−9^; OR_meta_ = 1.66), *NKILA-MIR4532* (rs72626599, *P*_meta_ = 6.48 × 10^−9^; OR_meta_ = 1.58), *RCOR1* (rs12887440, *P*_meta_ = 3.37 × 10^−8^; OR_meta_ = 1.48), and *FSTL5*-*MIR4454* (rs35213808, *P*_meta_ = 4.02 × 10^−8^; OR_meta_ = 1.49). Additionally, we found association at *ABCG2* and *SLC22A12* that was driven by low frequency SNVs. Furthermore, eight uncommon SNVs and three indels in the exome were predicted to be harmful. SNVs in *RCOR1* were linked to heightened blood leukocyte mRNA levels. THP-1 macrophage culture studies revealed the potential of decreased RCOR1 to suppress gouty inflammation.

**Interpretation:** Performing the first comprehensive characterization of adolescent-onset gout genomes identified risk loci of early-onset gout. Loci mediate inflammatory responsiveness to crystals that could mediate gouty arthritis. This study will contribute to risk prediction and therapeutic interventions to prevent adolescent-onset gout.

**Funding:** The National Natural Science Foundation of China and the National Key R&D Program of China.

**Research in context:** *Evidence before this study:* Gout is a polygenic disease and can present in adolescents and young adults. We searched PubMed for studies published as of Dec 31, 2021, without starting date or language restrictions and with the terms “adolescent-onset gout”, “early-onset gout”, “whole genome sequencing”, and “GWAS”, and no reports were found. Although GWAS have identified hundreds of genetic variants associated with gout and serum urate levels, they are all identified in adults (mean age 37.6-76.4 years old). The mechanism of early-onset gout is still unclear. The variants previously associated with early-onset gout are only in *ABCG2*. Due to the lack of large-scale genetic studies of the adolescent gout population, the mechanism of the early-onset gout is unknown.

*Added value of this study:* To the best of our knowledge, this is the first report of the comprehensive characterization of adolescent gout genomes. We identified common and uncommon risk loci of early-onset gout, most of which implicated in inflammation response, including *RCOR1*. SNVs in candidate risk gene *RCOR1* displayed expression regulation function. Knockdown of RCOR1 decreased IL-1β levels in THP-1 cells after MSU treatment. These immune-related genetic variants leading to heightened inflammatory responses to monosodium urate (MSU) crystals may contribute to early onset of gout in adolescents.

*Implications of all the available evidence:* This is the first report of the genetic landscape of adolescent-onset gout and increases our knowledge of the biological mechanisms underlying early-onset gout. The immune-related loci associated with early-onset gout discovered in this study are potential drug targets. Reducing inflammatory MSU crystal inflammatory responses to MSU crystals is a central objective in the prevention and treatment of adolescent-onset gout.

## Introduction

Gout is a polygenic inflammatory arthritis and mostly occurs in adult men, with an average age of onset > 45 years.^1^ Although rare, gout can occur in adolescents and young adults,^2,3^ and the mechanism of early-onset gout is still unknown. Adolescents with gout have higher urate levels, faster development of tophi, and are more likely to have a family history of gout compared with those who develop disease in adulthood.^4,5^ The percentage of adolescent-onset gout patients with a family history of gout is 38.7%-43.8%,^4,5^ compared to 25.9%-30% in gout with onset in early adulthood^6-8^ and 11%-23.8% in gout with the most common pattern of onset later in adulthood.^5,6,8^ This supports that the adolescent-onset gout population is more likely to have a strongest genetic background basis. The early onset of complex disease is often familial, with genetic risk factors having a relatively large impact on risk in younger patients.^9^ Therefore, genetic studies of adolescent-onset gout can provide novel insights into the mechanisms of gout and also may allow identification of new therapeutic targets for adolescents and adults with gout.

Most studies of the genetics of gout have focused on adult-onset disease. Hundreds of genetic variants associated with gout and serum urate levels have been identified by genome-wide association study (GWAS) in adults (mean age 37.6-76.4 years old).^10-12^ Other than rare mutations that cause HPRT deficiency, or PRS superactivity and major perturbations in purine metabolism,^13^ gene variants previously associated with gout with adult onset before age 40 have only been described in *ABCG2*,^14^ a high-capacity urate transporter that promotes intestinal and renal excretion of urate. Moreover, a Czechia study of 31 pediatric-onset patients, two common (p.V12M, p.Q141K) and three very rare (p.K360del, p.T421A, p.T434M) allelic *ABCG2* variants were detected.^15^

Routine array-based GWAS is common in adult gout patients, usually focusing on common variants, and imputation quality to study less common variants depends on the reference panel size and genetic distance from the target population.^16^ Deep whole-genome sequencing (WGS) is the gold standard to fully capture genetic variation, which provides the means to comprehensively analyse common variants, uncommon variants, and indels to discover etiological variants associated with disease.^17^ Here, we conducted WGS, association analysis, and replication in an adolescent-onset gout and an early-onset gout cohort (a total of 3739 individuals with onset ≤ age 30) to discover loci associated with early-onset gout.

## Methods

### Study design and participants

We did a WGS-based genetic study of adolescent-onset gout. Full details and references are provided in appendix (pp 2-6). We classified the single nucleotide variations (SNVs) into three types based on the minor allele frequency (MAF): common (MAF ≥ 0.05), low frequency (MAF < 0.05 and minor allele counts ≥ 2) and uncommon (MAF < 0.01 and minor allele counts ≥ 2). For common SNVs, we performed GWAS, Sequence Kernel association test (SKAT), and transcriptome-wide association analysis (TWAS) for the detection of adolescent gout/hyperuricemia associated loci. SKAT was also used for the identification of gout-related genes driven by low frequency SNVs. For uncommon SNVs and indels, we assessed for gout-specific functional variants or indels located in the exon regions using Fisher’s exact test. Candidate common SNVs were replicated in an independent early-onset gout. The meta-analyses were carried out with discovery and replication cohorts in METAL software to identify the significant loci associated with early-onset gout (*P* < 5 × 10^−8^).

Gout onset between 12-19 years old was defined as adolescent-onset gout. The adolescent-onset gout cohort included 280 individuals diagnosed with adolescent-onset gout, 191 individuals with hyperuricemia, and 434 normouricemic individuals. The independent early-onset gout replication cohort, defined as having gout with onset ≤ 30 years of age, comprised 824 early-onset gout, 926 hyperuricemic, and 1084 normouricemic individuals. Normouricemic individuals are controls that do not develop hyperuricemia or gout beyond the period of adolescence. All subjects in this study are male, because of gout is extremely rare in girls and young pre-menopausal women in the absence of monogenic syndromes such as Lesch-Nyhan syndrome. All subjects were recruited from the Affiliated Hospital of Qingdao University except 332 normouricemic individuals from Beijing Institute of Genomics, Chinese Academy of Sciences / China National Center for Bioinformation (BIG). All gout patients analysed in the study were examined by rheumatologists and met the 2015 American College of Rheumatology/European League Against Rheumatism classification criteria for gout.^18^ Hyperuricemia was defined as serum urate concentration > 7 mg/dL (> 420 μmol/L). The normouricemic controls were attained via site survey. Practice lists of normouricemic controls were screened for potentially suitable subjects by excluding those with hyperuricemia, diabetes, cancer and other arthritis-related illnesses. All participants provided written informed consent. In concordance with the Declaration of Helsinki, this study was reviewed and approved by the ethics committee of the Affiliated Hospital of Qingdao University (QYFYKYLL923011921) and BIG (2015H023).

### Procedures

To explore the genetic differences between gout and hyperuricemia, we classified all participants in the cohort into two comparison groups: gout vs individuals without gout (G-NG), and gout vs hyperuricemia (G-H). The GWAS was performed based on genotype data by PLINK software package (v1.9). Multivariable-adjusted logistic regression association analysis was performed with ten principal components and sequencing platform batches as covariables. SKAT on common and low frequency SNVs was performed by using the R-package SKAT (http://www.hsph.harvard.edu/skat/). SKAT CommonRare methods was used to evaluate the effect of common or low frequency SNVs separately. TWAS was performed by MetaXcan framework and the GTEx v.8 eQTL MASHR-M models (http://predictdb.org/). Considering the tissue-specificity of gene expression, TWAS was performed for data from five gout-relevant tissues - whole-blood, kidney cortex, colon sigmoid, colon transverse, and liver tissues. A meta-analysis across these tissues was performed by S-Multixcan. Candidate common SNVs for replication were selected using the following criteria: (1) Lead SNVs with a *P* ≤ 1 × 10^−4^ in either G-NG group or G-H group (LD, r^2^ < 0.2). (2) Four SNVs including rs2273406, rs41303970, rs7515191, and rs6680315, located in the *GCLM* region and three SNVs including rs8009475, rs35258120, and rs35785423 in the *RCOR1* region. Candidate common SNVs were genotyped by targeted sequencing using an Illumina Nova seq 6000. Case-control analysis of the variants was conducted using the same model as the GWAS. The meta-analyses were carried out in METAL software. The CADD scores (GRCh37-v1.6) of variant deleteriousness are available from https://cadd.gs.washington.edu/.

### Functional experiments

Total RNA from whole blood was extracted in adolescent individuals with the genotype of GG (n = 3), GA (n = 3), or AA (n = 3) at rs12887440 (*RCOR1*). Quantitative real-time polymerase chain reaction (RT-qPCR) and RNA-seq were conducted to measure the mRNA expression level of *RCOR1*, differentially expressed genes (DEGs), and enriched pathways of DEGs in different genotype of rs12887440. A human monocyte leukemia cell line (THP-1) was used for lentivirus virus transduction for a RCOR1 knockdown experiment. THP-1 cells were cultured with MSU crystals for 24 hours after being treated with PMA to induce macrophage differentiation, and supernatants and cells were harvested separately. Protein concentration of interleukin (IL)-1β in supernatants was determined by ELISA (E-EL-H0149c, Elabscience, China). Total protein was extracted from THP-1 cells to measure the protein level of RCOR1 and pro-IL-1β by Western Blot. All experimental data with a normal distribution were presented as mean ± standard deviation (s.d.) and analyzed with the aid of SPSS17.0 Statistical Software. Statistical significance was determined by one-way analysis of variance (ANOVA). *P* < 0.05 was considered to be statistically significant.

## Results

### Genomic resource of Chinese adolescent-onset gout cohort

We enrolled 905 male participants including 280 people with gout, 191 people with hyperuricemia, and 434 normouricemic individuals as Chinese adolescent-onset gout cohort. The mean ages at onset gout and hyperuricemia were 16.68 years (s.d. 1.79) and 17.38 years (s.d. 1.59), and mean age of controls was 29.38 years (s.d. 10.64) (table 1). As expected, the urate concentration in gout patients was significantly higher than that in individuals with hyperuricemia and normouricemia (figure 1A). The positive family history of gout in adolescents was 40.36% and 11.28% of individuals had palpable tophi (table 1).

**Figure 1.**
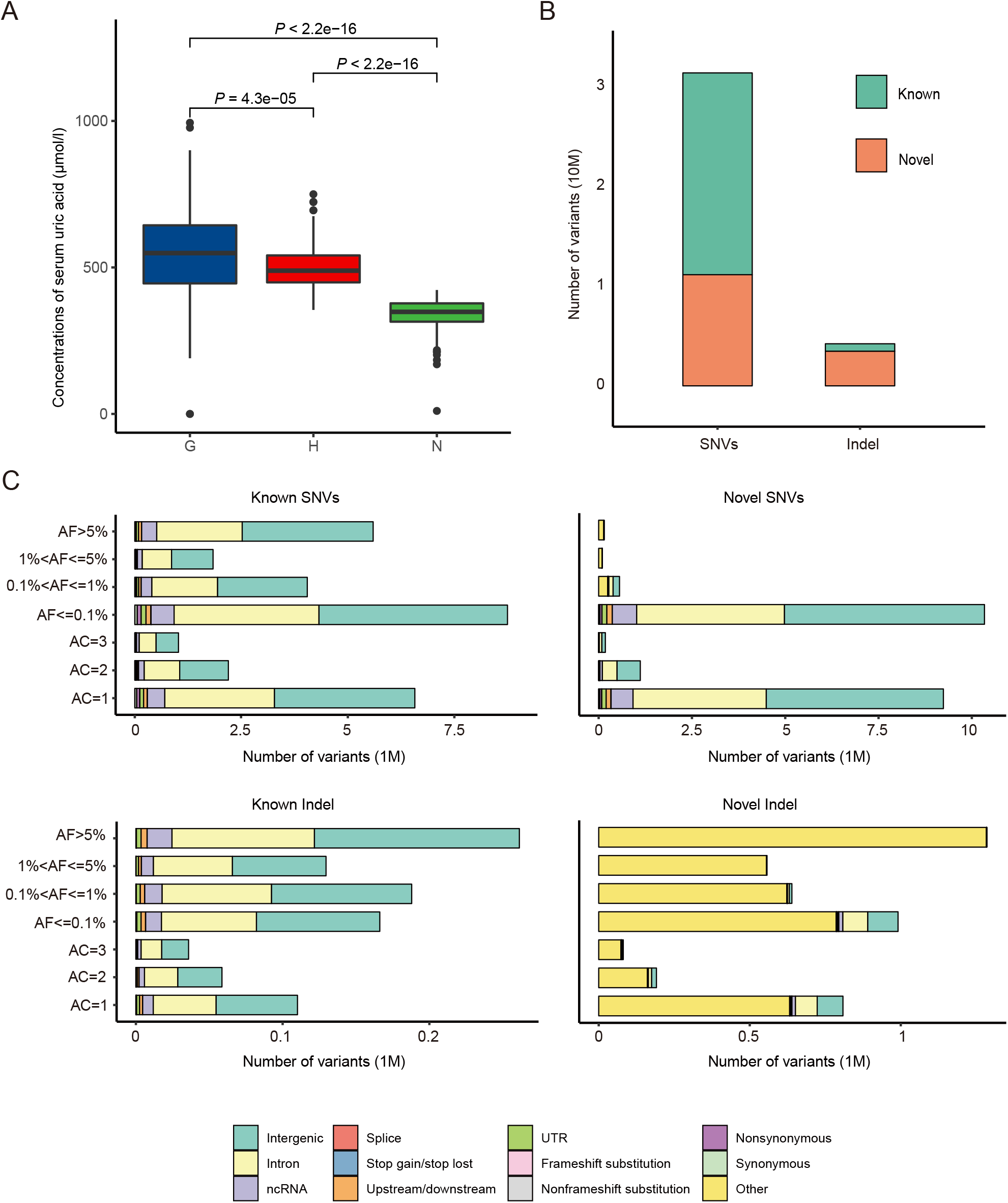
Genomic resource of adolescent gout cohort. (A) Serum urate concentrations in gout (G), hyperuricemic (H), and normouricemic (N) groups. *P* values were determined by t-test. (B) The number of known and novel SNVs and indels identified. (C) The distribution of allele frequency and genomic element of SNVs and indels in adolescent gout cohort.

We performed WGS with a median sequencing coverage of 25x in an adolescent-onset gout cohort. We identified a total of 31.36 million single nucleotide variations (SNVs) and 4.22 million indels. Compared with other public datasets, including dbSNP (v150), ExAC (v0.3), the gnomAD genome collection (v2.1.1), NHLBI-ESP project, and the 1000 Genomes Project, we obtained 11.14 million novel SNVs and 3.47 million novel indels (figure 1B). As expected, most (92.84%) novel SNVs were uncommon (allele frequency, AF ≤ 0.1%), with singletons, doubletons and tripletons accounting for 82.95%, 10.02%, 1.62% of novel SNVs, respectively (figure 1C). Of the 35.58 million genetic variants, 80.65% were located in intronic and intergenic regions, only 0.34 million SNVs were found in protein-coding regions. The distributions the genetic variants were 20.15% common SNVs (MAF ≥ 0.05, 7.17 million), 79.85% low frequency SNVs (MAF < 0.05, 28.41 million), and 72.71% uncommon SNVs (MAF < 0.01, 25.87 million).

### Identification of risk loci from common variants

We first performed GWAS on common SNVs in the G-NG group (272 gout vs 602 individuals without gout) and the G-H group (272 gout vs 185 individuals with hyperuricemia) after the quality control (appendix p 2). A total of 571 and 550 gout-associated SNVs were identified as putative associations (*P*_discovery_ < 1.0 × 10^−4^) from the G-NG and G-H groups, respectively (appendix pp 8-50). Among these, all SNVs that reached nominal genome-wide significance (*P*_discovery_ < 5.0 × 10^−8^) were located in the 4q22.1 region (figure 2A and B), which contains the *ABCG2* locus, established also in adult gout.

**Figure 2.**
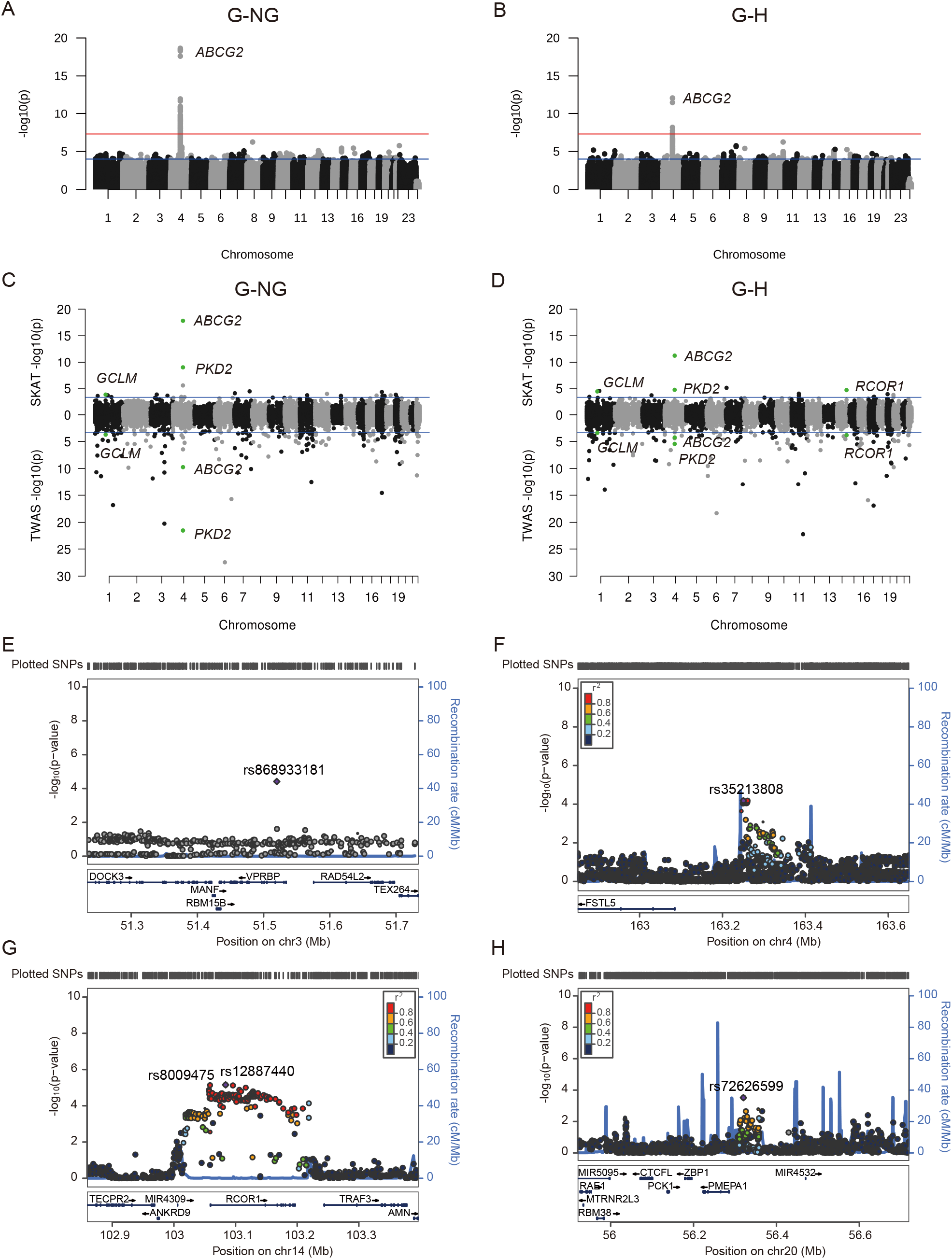
Manhattan plots and regional association plots for the loci identified from GWAS, SKAT, and TWAS. Manhattan plot of GWAS in the G-NG group (A) and the G-H group (B). The upper red horizontal line indicates the genome-wide significance threshold (*P* = 5.0 × 10^−8^). The lower blue horizontal line indicates the cut-off level for selecting SNVs for replication (*P* = 1.0 × 10^−4^). GWAS, genome-wide association study. Gene-level Manhattan plot from SKAT (upper) and TWAS (lower) of G-NG group (C) and G-H group (D). The blue solid horizontal line indicates the cut-off level for selecting genes (*P* = 5.0 × 10^−4^). TWAS, transcriptome-wide association study; SKAT, sequence kernel association test. Regional association plot for four novel loci identified from meta-analysis: *VPRBP* (E), *FSTL5*-*MIR4454* (F), *RCOR1* (G), and *NKILA-MIR4532* (H). The vertical axis represents –log10(*P* value) for the test of SNV association with gout in the GWAS stage. SNV with the lowest *P* value is depicted as a pink diamond and neighboring SNVs are colored according to the extent of linkage disequilibrium (measured in r^2^) based on 1000 Genomes phase 3. Genomic coordinates are based on NCBI human genome reference sequence build 37.

We performed both TWAS and gene-based SKAT on common SNVs to evaluate the joint effect of SNVs at the gene level. From the results of TWAS, SKAT and GWAS, four genes reached suggestive transcriptome- and genome-wide significance (*P*_TWAS_ < 5 × 10^−4^, *P*_GWAS_ < 1 × 10^−4^, and *P*_SKAT_ < 5 × 10^−4^), including the known genes *ABCG2, PKD2*, a novel gene *GCLM* significant in both the G-NG and G-H groups (figure 2C and D), and a novel gene *RCOR1* significant only in the G-H group (figure 2D).

For validation, 166 independent association signals (linkage disequilibrium, LD, r^2^ < 0.2), combined with seven SNVs in *GCLM* (rs7515191, rs6680315, rs2273406, rs41303970) and *RCOR1* (rs8009475, rs35258120, rs35785423) included in the TWAS were genotyped in an independent Chinese case-control cohort (n = 2834; gout = 824, hyperuricemia = 926 and normouricemia = 1084). Due to the low prevalence of adolescent-onset gout, we recruited gout patients with age of onset ≤ 30. The mean age of participants in the replication stage was 24.92 (s.d. = 4.44), 18.30 (s.d. = 0.54) and 16.70 (s.d. = 2.23) for gout, hyperuricemia, and normouricemia, respectively. There were 26 and 36 variants with modest evidence of association in the replication data set (*P*_replication_ < 0.05) of the G-NG and G-H group comparisons, respectively (appendix pp 51-53). Subsequently, we performed meta-analysis combining the results from the discovery and replication datasets. At the genome-wide significance level (*P* < 5.0 × 10^−8^), in addition to SNVs which had already been reported for gout-associated loci (*ABCG2* and *PKD2*, appendix p 54), we identified one novel association in the G-NG group: rs868933181 of *VPRBP* (*P*_meta_ = 6.27 × 10^−9^; OR_meta_ = 1.66), and four novel associations in the G-H group: rs35213808 of *FSTL5*-*MIR4454* (*P*_meta_ = 4.02 × 10^−8^; OR_meta_ = 1.49), rs12887440 of *RCOR1* (*P*_meta_ = 3.37 × 10^−8^; OR_meta_ = 1.48), rs8009475 of *RCOR1* (*P*_meta_ = 4.18 × 10^−8^; OR_meta_ = 1.48; r^2^ = 0.90 with rs12887440), and rs72626599 of *NKILA-MIR4532* (*P*_meta_ = 6.48 × 10^−9^; OR_meta_ = 1.58, figure 2E-H, table 2). However, the candidate gene *GCLM* detected in the discovery stage did not reach the genome-wide significance level in the replication stage (appendix p 57).

### Functional regulation of RCOR1 by identified SNVs

Since *RCOR1*, identified by the analysis of GWAS, TWAS, and SKAT in the discovery stage also reached the genome-wide significance level after meta-analysis, we functionally annotated the SNVs in *RCOR1*. The two maximally-associated SNVs were located in the promoter (rs8009475) and the second intron (rs12887440) of *RCOR1*. The region harboring rs12887440 was enriched with active histone modification markers (H3K4me1 and H3K27ac) in gout-relevant cells and tissues, including E029 (primary monocytes from peripheral blood), E124 (monocytes-CD14^+^ RO01746 primary cells), E066 (liver), E062 (primary mononuclear cells from peripheral blood) and E034 (primary T cells from peripheral blood). Furthermore, Hi-C evidence^19^ suggested that this region regulates the promoter (harboring rs8009475) by a long-range chromatin conformation interaction. In addition, we observed that rs12887440 associated with 14 DNA methylation (DNAm) sites (*P* < 1 × 10^−5^) in a whole blood DNA methylation QTL dataset.^20^ Of the 14 DNAm sites, cg10501036 (511 bp from rs8009475) was located in the *RCOR1* promoter (figure 3A). These findings suggest that rs12887440 may influence *RCOR1* expression. To confirm this, we measured the expression of *RCOR1* in the whole blood of nine individuals from the adolescent gout cohort with the genotype of GG (n = 3), GA (n = 3), or AA (n = 3) at rs12887440 through RT-qPCR. The homozygous risk allele carriers (AA allele) displayed significantly higher *RCOR1* mRNA levels in their whole blood compared to the non-carriers (GG allele; figure 3B), which suggests that rs12887440 associates with the expression of *RCOR1*.

**Figure 3.**
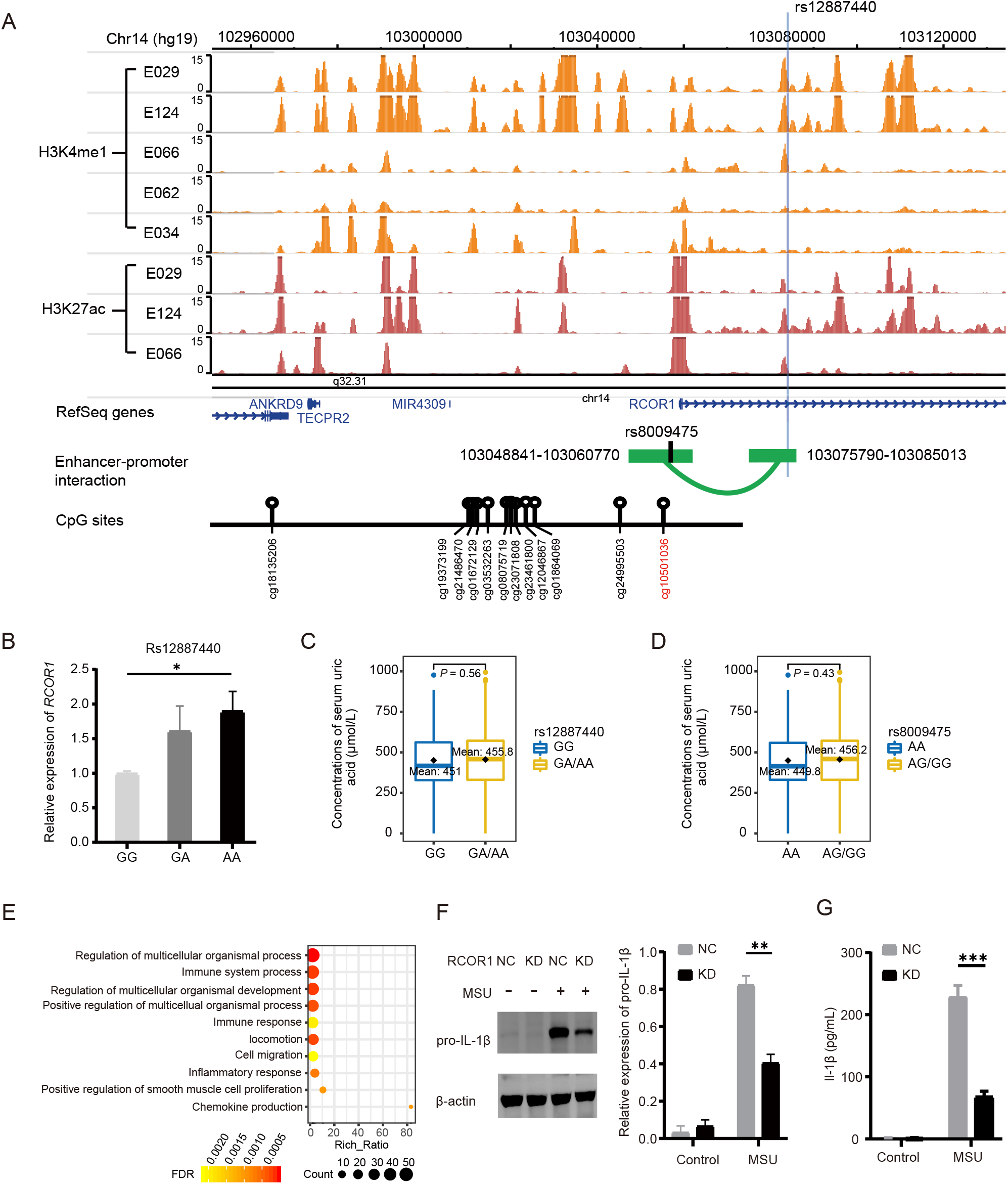
*RCOR1* is involved in the immune process of gout. (A) Functional annotation of rs12887440. The locus where rs12887440, highlighted with a blue vertical line, resides is enriched with both H3K4me1 (orange) and H3K27ac (brown) modifications, including in E029 (primary monocytes from peripheral blood), E124 (monocytes-CD14+ RO01746 primary cells), E066 (liver), E062 (primary mononuclear cells from peripheral blood) and E034 (primary T cells from peripheral blood) cell lines or tissue, suggesting an enhancer function of this region. The interaction of rs12887440-residing region with the promoter of *RCOR1* where rs8009475 were located as revealed by Hi-C schematic representation (green curve) in the GM12878, H1ESC, Left_Ventricle, MSC, Psoas, Right_Ventricle, and Troproblast sourced from 3D Genome browser. Genomic coordinates are based on NCBI human genome reference sequence build 37. The CpGs associated with rs12887440 are displayed below the interaction curve. The CpG residing in the promoter of *RCOR1* is marked in red. (B) The expression of *RCOR1* was significantly increased in the rs12887440 risk allele AA carriers compared with non-risk allele GG carriers (*P* < 0.05), whereas no significant difference was observed between GA carriers and GG carriers (*P* > 0.05). Data are mean ± s.e.m., n = 3. Association between serum urate concentration and rs12887440 alleles (C) and rs8009475 alleles (D). (E) The top 10 biological process terms enriched by DEGs in whole blood of rs12887440-AA carriers compared with that of rs12887440-GG carriers. The x-axis shows the ratio of input DEGs that are annotated in a term to all genes that are annotated in this term (rich ratio), and the y-axis represents the enriched biological process terms. The color scale represents the false discovery rate (FDR) for each enriched biological process and the dot size indicates the number of genes involved in a particular process. The protein expression of pro-IL-1β via western blotting, using β-actin as an internal control (F) and the levels of IL-1*β* secretion in the supernatant via ELISA (G) in *RCOR1*-knockdown (KD) THP-1 cells after MSU crystal treatment as indicated. -, MSU untreated; +, MUS treated. NC, Negative control THP-1 cells without *RCOR1* treatment. Data are mean ± s.e.m., n = 3. *P* values in panel b,c,d,e, f and g were determined by t-test. * *P* < 0.05, ** *P* < 0.01, *** *P* < 0.001.

### *RCOR1* is involved in the inflammatory process of gout

Since *RCOR1* was identified to be associated with gout in the G-H group, we hypothesized that its role was in the immune process and not via urate homeostasis. To confirm this, we first performed an association analysis between *RCOR1* SNVs and serum urate concentrations in all participants included in the discovery and replication stages, and found that rs12887440 and rs8009475 were not associated with serum urate (*P*_rs12887440_ **=** 0.56, *P*_rs8009475_ **=** 0.43, figure 3C and D). As mentioned above, rs12887440 AA allele carriers have higher *RCOR1* expression, thus we performed RNA-Seq on the whole blood of the same patients tested for RT-qPCR to explore the effect of higher *RCOR1* expression in patients with the risk allele (A allele). Compared with the rs12887440 GG carriers, we identified 341 DEGs in rs12887440 AA allele carriers. As the top 10 Gene Ontology (GO) biological process terms showed, these DEGs were involved in immune-related pathways such as immune system process, immune response, inflammatory response, and chemokine production (figure 3E), suggesting that *RCOR1* is possibly involved in the inflammatory process of gout.

To examine whether RCOR1 suppresses or promotes inflammation in gout development, we used THP-1-derived macrophages where RCOR1 was knocked down (RCOR1-KD) to determine the function of RCOR1 in MSU crystal-induced inflammation. The cells did not express pro-IL-1*β* and IL-1*β* when there was no inflammatory stimulus, regardless of the expression level of RCOR1. However, after exposure to MSU crystals to stimulate the immune response, the protein levels of intracellular pro-IL-1*β* and IL-1*β* levels in the supernatant were significantly reduced in RCOR1-KD cells compared with control cells (figure 3F and G). These findings demonstrate that RCOR1 could be actively involved in development of gouty inflammation.

### Gene-based SKAT for low frequency SNVs

To determine whether low frequency SNVs (MAF < 0.05) associate with gout and hyperuricemia in adolescents, we conducted gene-based SKAT on low frequency SNVs with minor allele counts ≥ 2. As anticipated, *ABCG2* reached genome-wide significance level in the G-NG group (*PABCG2* = 1.81 × 10^−8^) after Bonferroni correction (*P*_Bonferroni_ < 2.09 × 10^−6^, 23,869 gene sets). Solute carrier family 22 member 12 (*SLC22A12*), which associates with serum urate levels,^21^ was also significant in the G-NG group (*PSLC22A12* = 1.06 × 10^−8^), consistent with multiple previous reports of association of uncommon variants with hypouricemia.^12,22^ There were no genes significantly associated in the G-H group (appendix pp 58-75).

### Gout-specific exonic uncommon variants

To detect functional gout-specific uncommon variants (MAF < 0.01), we calculated the difference of mutation ratio in samples and observed eight non-synonymous, deleterious, uncommon SNVs and three uncommon indels affecting the primary structure of proteins (Fisher’s exact test, *P* value < 0.05). Eight uncommon SNVs located in *LIPH, PKHD1, SLC12A3, DDR2, PKP1, ITGAL, SCN10A*, and *MCOLN3*, and three indels located in *TRIM67, MAIP1*, and *DOC2A* were identified only in gout cases (appendix p 76).

Although not statistically significant, there were six gout-specific heterozygous, non-synonymous, uncommon SNVs located in a 1672 bp exonic region of *IL37*, a gene previously implicated in gout.^23^ Of these, the Combined Annotation Dependent Depletion (CADD) scores of rs2708943, rs2723187, rs2723192, and rs532660211 were > 10 (table 3), suggesting that the variants have a deleterious effect. Moreover, five that were in strong LD (r^2^ = 1), including rs2708943, rs2723183, rs2723187, rs2708947, and rs2723192, were detected in two adolescent-onset gout individuals simultaneously. Four of them had previously been reported in gout (rs2708943, rs2723183, rs2723187, and rs2708947).^24^

We also investigated uncommon SNVs and indels of *RCOR1*. There were 10 SNVs and 12 indels that showed significantly different allele counts in the G-NG group, and four SNVs and nine indels with significantly different allele counts in the G-H group, respectively (Fisher’s exact test, *P* value < 0.05, appendix pp 77-78).

## Discussion

This study comprehensively characterized adolescent-onset gout genomes. In addition to the gout risk locus *ABCG2* previously linked in a small Czechia study to pediatric onset gout,^15^ and to *ABCG2* and *PKD2* identified in previous GWAS of gout,^12^ genetic analyses of the WGS data allowed us to identify four novel common risk loci, uncommon variants and indels associated with gout. These results provide more insights into the genetic basis of gout than traditional array-based genotyping analyses by utilizing deep sequencing data in people with early-onset gout who are expected to have a stronger genetic contribution to disease.

Genetic variants related to immune response were identified in the adolescent-onset gout cohort. Regarding common variants, four novel candidate gout-risk loci (*RCOR1, VPRBP, NKILA-MIR4532*, and *FSTL5*-*MIR4454*) were identified, of which one (*RCOR1*) may have a role in inflammation and immunity. REST corepressor 1 (*RCOR1* or CoREST), identified to be associated with gout in the G-H group, is a protein that binds to the C-terminal domain of repressor element-1 silencing transcription factor (REST) and regulates diverse immune and inflammatory responses.^25^ *Rcor1* deletion disrupts Foxp3-dependent recruitment of the CoREST complex to the promoters of T-bet, IL-2, and IFN-γ, leading to impaired Treg function *in vivo* and promotes antitumor immunity in mice.^26^ T-bet deficiency alleviates MSU-crystal-induced inflammatory cell infiltration in peritonitis and air pouch models *in vivo*, in addition to the IL-1β levels of air pouch lavage fluid.^27^ In our study, the risk allele A of lead SNV rs12887440 in *RCOR1* had a higher allele frequency in gout cases compared with hyperuricemia and we observed that the risk allele increases the expression of *RCOR1*. The A allele of rs12887440 and the G allele of rs8009475 were not associated with serum urate levels (figure 3C and D). Functional experiments in THP-1 macrophage cells indicated *RCOR1* has a role in promoting gouty inflammation. These findings suggest that the *RCOR1* locus is pro-inflammatory and contributes to the progression from hyperuricemia to gout.

*VPRBP*, also called *DCAF1* is considered to be a general substrate-recognizing subunit of E3 ligases, and has been implicated in various cellular processes. *VPRBP* was reported to inhibit the NF-κB pathway by inhibiting the nuclear transportation of p65 in HEK293 cells.^28^ Activation of NF-κB in macrophages and monocytes by MSU crystals is of particular relevance for the initiation of the gout flare.^1^ We also identified rs72626599, a SNV in the intergenic region between *NKILA* and *MIR4532*, as a new gout locus. Studies conducted in human inflammation-stimulated breast epithelial cells have shown that the lncRNA *NKILA* (NF-kB-interacting lncRNA) binds to and blocks phosphorylation sites on IkB, thereby inhibiting IKK-induced IkB phosphorylation and NF-kB activation.^29^ NKILA was downregulated in osteoarthritic cartilage tissues, and inhibits the SP1/NF-κB signaling pathway of chondrocytes in the progression of osteoarthritis.^30^ We also identified rs35213808, a SNV in the intergenic region between *FSTL5* and *MIR4532. FSTL5* inhibits the Wnt/β-catenin signaling pathway in a hepatocellular carcinoma cell line.^31^ In this study, rs868933181 in *VPRBP*, rs72626599 in *NKILA-MIR4532*, and rs35213808 in *FSTL5*-*MIR4454* were all associated with serum urate levels (appendix p 7), suggesting that they may play dual roles by affecting serum urate levels and inflammation in the development of gout in adolescents.

Besides common variants, we also found low frequency SNVs, uncommon SNVs and indels associated with adolescent gout. Low frequency SNVs were associated with adolescent gout at *ABCG2* and *SLC22A12*, which have previously been reported to be associated with hyperuricemia and gout.^12,15,32^ The rare variant p.(n182s) (rs752113534) of IL37 causes loss of anti-inflammatory activity of IL-37 and increases the risk of developing gout in hyperuricemic individuals of Polynesian ancestry.^23^ In this study, we found six gout-specific heterozygous, non-synonymous, uncommon SNVs of IL37 in the adolescent-onset gout cohort. Our results will improve our understanding of the role of uncommon variants in gout progression.

The genetic risk variants in adolescent-onset gout patients may contribute to clinical manifestations such as faster development of tophi and the earlier onset of gout. In this adolescent-onset gout study, we discovered several novel loci, such as *RCOR1* and *NKILA*, related to immunity response. Immune-associated genetic variants for gout in adolescence may result in individuals being more sensitive to the inflammatory response to MSU crystals, leading to early onset of gout. As our experiments found the knockdown of RCOR1 caused lower IL-1β levels that showed RCOR1 acted as a role in promoting gouty inflammation in THP-1 cells after MSU treatment. Greater sensitivity to MSU crystal inflammatory responses may be a major cause of early onset of gout. This study provides insight into the pathogenesis of gout in adolescents.

However, due to the low prevalence of gout in adolescents, this study had limitations in discovering more gout-related genes and uncommon variants in adolescents. In addition, our study shows that the newly discovered adolescent-onset gout risk genes mainly have the potential to regulate gout inflammation and immunity, but the specific regulatory mechanisms remain to be elucidated. The functional RCOR1 experiments in THP-1 macrophage cells probed effects on the inflammatory phenotype of the disease. However, a much larger scope of studies would be needed to examine the etiology independent of hyperuricemia of developing gout, which requires urate crystal deposition in the joint. That process involves the joint surface lubrication by lubricin, collagen II release from cartilage, and homeostasis of articular cartilage, synovium, and other tissues in the joint, which is disrupted by several proteases released by inflammatory cells.^33^ Moreover, there are 16419 target genes of the RCOR1 transcription factor repressor in ChIP-seq datasets from the ENCODE Transcription Factor Targets dataset (http://www.ncbi.nlm.nih.gov/gene/23186). They include not only NLRP3, L-1β and IL-6, but also multiple genes that regulate lubricin proteolytic turnover (ITIH3, THBS1, ELANE, TLR2, SERPINB6).^33^ Notably, RCOR1 also modulates articular chondrocyte homeostasis and cartilage integrity.^34^

This is the first report of comprehensive characterization of adolescent-onset gout genomes, and risk loci of early-onset gout were identified. Novel loci involved in inflammatory response may contribute to early-onset gout. This study will contribute a better understanding of the mechanisms of the adolescent-onset gout and be helpful for drug development, risk prediction, and intervention to prevent adolescent-onset gout and early-onset gout.

## Data Availability

Adolescent gout GWAS summary statistics and the genotype data can be made available on request to the corresponding author.

## Contributors

CL took full responsibility for the work, had access to the data, and controlled the decision to publish. CL, XF, HQ, and TRM. conceived and supervised the project. AJ and YS analyzed the data. XJ, CW, CZ, and JX collected the samples. YS, ZF, YL, and GZ performed the RT-qPCR of rs12887440. MS and JL performed RNA-seq analysis. XX, FY, ML, LC, ZL, XL, and LH performed knockdown of *RCOR1*. WS, YH, HZ, XW, LP, HQ, YL, and ZC provided statistical support. RT and ZL offered genetic consult. AJ and YS drafted the manuscript. JW, HQ, YS, RT, ND, RT and TRM revised the manuscript. All the authors read and approved the final manuscript.

## Acknowledgments

This work was supported by the National Natural Science Foundation of China (Grant Nos. 82220108015 and 82201957) and National Key R&D Program of China (Grant Nos. 2022YFC2503300, and 2022YFE0107600).

We are grateful to all anonymous participants in adolescent cohort including part of normal control which was supported by grant of Key Program of the Chinese Academy of Sciences (KJZD-EW-L14).

## List of Tables

**Table 1 Population characteristics of adolescent-onset gout cohort**

**Table 2 Novel SNVs associated with gout at a genome-wide level of significance**

**Table 3 Six non-synonymous, uncommon SNVs of *IL37* in gout cases**

