## Supplemental materials for "Novel genetic loci in adolescent-onset gout derived from whole genome sequencing of a Chinese cohort"

**Appendix Methods**

**Study cohort**

Gout onset between 12-19 years old was defined as adolescent-onset gout. The adolescent-onset gout cohort included 280 individuals diagnosed with adolescent-onset gout, 191 individuals with hyperuricemia, and 434 normouricemic individuals. The independent early-onset gout replication cohort, defined as having gout with onset ≤ 30 years of age, comprised 824 early-onset gout, 926 hyperuricemic, and 1084 normouricemic individuals. Normouricemic individuals are controls that do not develop hyperuricemia or gout beyond the period of adolescence. All subjects in this study are male, because of gout is extremely rare in girls and young pre-menopausal women in the absence of monogenic syndromes such as Lesch-Nyhan syndrome. All subjects were recruited from the Affiliated Hospital of Qingdao University except 332 normouricemic individuals from Beijing Institute of Genomics, Chinese Academy of Sciences / China National Center for Bioinformation (BIG). All gout patients analysed in the study were examined by rheumatologists and met the 2015 American College of Rheumatology/European League Against Rheumatism classification criteria for gout.[1] Hyperuricemia was defined as serum urate concentration > 7 mg/dL (> 420 μmol/L). The normouricemic controls were attained via site survey. Practice lists of normouricemic controls were screened for potentially suitable subjects by excluding those with hyperuricemia, diabetes, cancer and other arthritis-related illnesses. All participants provided written informed consent. In concordance with the Declaration of Helsinki, this study was reviewed and approved by the ethics committee of the Affiliated Hospital of Qingdao University (QYFYKYLL923011921) and BIG (2015H023).

**Whole genome sequencing and data processing**

DNA of the GWAS cohort was extracted from whole blood by the Qiagen Blood Midi Kit (Qiagen, Hilden, Germany). Qingdao data were generated by BGISEQ-500 WGS library preparation[2] and BGISEQ-500 (PE 100) platform. Data from BIG were generated by NEXTflex Rapid DNA-seq Kit (Catalog No. 5144-08, Bioo Scientific Corporation, Austin, TX, USA) library preparation and Hi-Seq X Ten, Hi-Seq 3000, and Hi-Seq 4000 platforms. Adaptor sequences and low quality bases were removed using SOAPnuke (v1.5.6),[3] and filtered reads were aligned to human reference genome (hg19) using BWA (Burrows-Wheeler Aligner, v0.7.12) with default parameters.[4] The duplicated reads of PCR amplification were removed using Picard tool (http://picard.sourceforge.net). GATK (v3.3)[5] HaplotypeCaller was used for variants calling and variants were further filtered in confidence level 99.0 with Variant Quality Score Recalibration (VQSR) and then annotated with ANNOVAR (v2018-04-16).[6]

**GWAS**

The GWAS was performed by PLINK software package (v1.9).[7] Individuals with ambiguous sex, genotype missing rate greater than 0.05, and heterozygosity rate exceeding ± 3 standard deviations as well as outliers that greatly departed from other samples detected by principal component analysis were excluded (PC1 < -0.05 or PC2 < -0.05). Variants with genotype missing rate > 0.05, MAF < 0.05, as well as departures from Hardy-Weinberg equilibrium ($P < 1 \times 10^{-6}$) were also removed. Multivariable-adjusted logistic regression association analysis was performed with ten principal components as covariables. To correct the influence of different sequencing platforms, sequencing platform batches were also included as covariates. The online tool LocusZoom (http://locuszoom.sph.umich.edu/)[8] was performed to visualize specific regions.

**Gene-based SKAT**

Sequence Kernel association test (SKAT)[9] on common and low frequency variants was performed by using the R-package SKAT (http://www.hsph.harvard.edu/skat/). SKAT CommonRare methods was used to evaluate the effect of common (MAF ≥ 0.05) or low frequency variants (MAF < 0.05 and MAC ≥2) separately.

**TWAS**

TWAS was performed by MetaXcan framework and the GTEx v.8 eQTL MASHR-M models (http://predictdb.org/).[10] Firstly, GWAS results were harmonized and lifted over to hg38. TWAS was then performed for data from five tissues including whole-blood, kidney cortex, colon sigmoid, colon transverse, and liver tissues, separately. Finally, a meta-analysis across these tissues was performed by S-Multixcan to test the joint effect of genes on phenotype in different tissues.

**Uncommon variants and indels in gout cases**

Uncommon variants and indels (MAF < 0.01 and MAC ≥ 2) discovered in gout and with allele count/count of 0 in both hyperuricemia and normouricemia controls were defined as gout-specific. Deleterious variants were annotated "D" in 11 ANNOVAR algorithms including SIFT_pred, Polyphen2_HDIV_pred, Polyphen2_HVAR_pred, LRT_pred, fathmm.MKL_coding_pred, MetaSVM_pred, MetaLR_pred, FATHMM_pred, PROVEAN_pred, ClinPred_pred, M.CAP_pred. We conducted an association analysis using a 2×2 contingency table based on mutated samples. For each of the filtered SNVs and indels, the $P$ value of association was assessed using Fisher's exact test. Allele frequencies of *IL37* variants in different ancestral groups were taken from the gnomAD Database v3.1.2 (http://gnomad.broadinstitute.org). The CADD scores (GRCh37-v1.6) of variant deleteriousness[11] are available from https://cadd.gs.washington.edu/.

**Replication**

Candidate common SNVs for replication were selected using the following criteria: (1) Lead SNVs with a $P \leq 1 \times 10^{-4}$ in either the G-NG group or G-H group (LD, $r^2 < 0.2$). (2) Four SNVs including rs2273406, rs41303970, rs7515191, and rs6680315, located in the *GCLM* region from the TWAS analysis and three SNVs including rs8009475, rs35258120, and rs35785423 in the *RCOR1* region from the TWAS analysis. DNA of the replication cohort was extracted from whole blood by the Magnetic Whole Blood Genome Extraction Kit (NanoMagBio, Wuhan, China) and NEXTflex Rapid DNA-seq Kit (Catalog No. 5144-08, Bioo Scientific Corporation, Austin, TX, USA) was used to prepare libraries. Candidate common SNVs were genotyped by targeted sequencing using an Illumina Nova seq 6000. Case-control analysis of the variants was conducted using the same model as the GWAS. The meta-analyses were carried out in METAL software[12] using the Standard Error SE model (STDERR) which takes into account effect size estimate and the association results for each SNV across GWAS and replication data sets were combined.

**Functional annotation of *RCOR1***

Chromatin state data from the Roadmap epigenomics project[13] was used to identify areas of regulatory function or active transcription. The Washington University epigenome browser[14,15] was used to visualize the genomic area encompassing rs12887440 and mapped the regulatory overlap with H3K4me1 and H3K27ac histone modification marks. The 3D Genome browser's Hi-C data[16-19] were used to investigate chromatin interactions. The GoDMC data set[20] was used to identify DNAm of rs12887440. Location of DNAm site was annotated by DNMIVD.[21]

**Quantitative real-time polymerase chain reaction (RT-qPCR)**

Whole blood from gout patients was collected by PAXgene Blood RNA Tubes (No. 762165, Qiagen) and total RNA from whole blood was extracted by PAXgene Blood RNA Kit (No. 762174, Qiagen). RNA was reverse-transcribed into cDNA by PrimeScript RT reagent kit (No. RR047A, Takara Bio, Kusatsu, Shiga, Japan). RT-qPCR was performed to quantify the relative expression of *RCOR1* based on the SYBR Green Universal Master Mix (No. KK4601, KAPA Biosystems). The expression of each target gene was quantified by the $2^{-\Delta\Delta Ct}$ method against *GAPDH* for normalization.

**RNA-seq and data processing**

The concentration and integrity of RNA was detected by Agilent 2100 RNA nano 6000 assay kit (No. 5067-1511, Agilent Technologies, CA, USA). Sequencing libraries were generated using VAHTS Universal V6 RNA-seq Library Prep Kit for Illumina® (NR604-01/02) following the manufacturer's recommendations and index codes were added to attribute sequences to each sample. The cluster generation and sequencing were performed on Novaseq 6000 S4 platform, using NovaSeq 6000 S4 Reagent kit V1.5. Bowtie2 (v2.2.3) software[22] was used for building the genome index, and clean data was then aligned to the reference genome using HISAT2 (v2.1.0)[23]. Read counts for each gene in each sample were counted by HTSeq (v0.6.0),[24] and FPKM (Fragments Per Kilobase Millon Mapped Reads) was then calculated to estimate the expression level of genes in each sample.

Differentially expressed genes (DEGs) were determined by DESeq2 R package (v1.6.3)[25] with the criteria of adjusted p value less than 0.05 and absolute value of fold change greater than 1.5. The GO (Gene Ontology, http://geneontology.org/) enrichment analyses of DEGs was implemented by the hypergeometric test.

**Cell culture**

A human monocyte leukemia cell line (THP-1) was obtained from the Cell Bank of the Type Culture Collection of the Chinese Academy of Sciences. The cells were grown in RPMI-1640 medium supplemented with 10% heat-inactivated fetal bovine serum at 37°C and 5% $CO_2$. THP-1 cells were plated at a density of $1.0 \times 10^6$/mL in 6-well plates.

**Lentivirus Virus Production and Transduction**

A lentiviral (LV) knockdown vector containing enhanced green fluorescence protein (GFP) was constructed by GENECHEM (Shanghai, China). The cDNA sequences of RCOR1 were amplified and used to synthesize knockdown lentiviral particles for Human RCOR1 (LV-RCOR1-RNAi) and their controls (LV-NC). THP-1 was transfected with a multiplicity of infection (MOI, 30) of LV-RCOR1-RNAi or LV-NC. After infection for 72 h, puromycin was added to the culture medium for stable cell selection. The infection rate was evaluated through the expression of green fluorescent protein. PMA (500 ng/mL) was used to treat THP-1 cells in a six-well plate ($2 \times 10^6$ cells/well) for 3 h, and THP-1-derived macrophages were obtained.[26] MSU crystal suspension (200 μg/mL) stimulated THP-1-derived macrophages for 24 h.

**Cytokine ELISA assay**

THP-1 cells were cultured with MSU for 24 hours after PMA treatment, and the supernatants harvested. Protein concentrations of interleukin (IL)-1β in supernatants was determined by ELISA (E-EL-H0149c, Elabscience, China), according to the manufacturer's instructions.

**Western Blot for RCOR1 and pro-IL-1β Protein Measurement**

Total protein was extracted from THP-1 cells with RIPA buffer, and the protein concentration was determined using BCA Protein Assay Kit (Thermo Scientific, MA, USA). The total protein (μg) was mixed with sample buffer, boiled for 10 min, and electrophoresed on a 10% SDS-PAGE gel. Separated proteins on the gel were transferred to polyvinylidene fluoride (PVDF) membranes (Bio-Rad, Hercules, CA, USA). After blocking with 5% BSA for 1 h at room temperature, the primary antibody of RCOR1(#14567S, CST, USA) and pro-IL-1β (#27989SF, CST, USA) was incubated at 4°C overnight (diluted at 1:1000). Then, PVDF membranes were incubated with the secondary antibody (E-AB-1001, Elabscience, China) (diluted at 1:10,000) for 1 h at room temperature and visualized using a gel imaging system with a ECL chemiluminescence kit (CB08669050, Wuhan Chemstan Biotechnology, China). The pictures were analyzed with ImageJ software to calculate the gray scale ratio of β-actin (81115-1-RR, Proteintech, China).

**Statistical Analysis**

All experimental data with a normal distribution were presented as mean ± standard deviation (s.d.) and analyzed with the aid of SPSS17.0 Statistical Software. Statistical significance was determined by one-way analysis of variance (ANOVA). For data with equal variances assumed, ANOVA followed by the Least significance difference test was applied. For data with equal variances not assumed, ANOVA followed by Dunnett's T3 test was used. Data in bar graphs are expressed as means ± SEM. A probability of less than 0.05 was considered to be statistically significant.

**Appendix Figure**

**Appendix Figure 1. The correlation of gout risk loci with serum uric acid concentration.**

Association between serum uric acid concentration and rs868933181 (A), rs72626599 (B), and rs35213808 alleles (C).

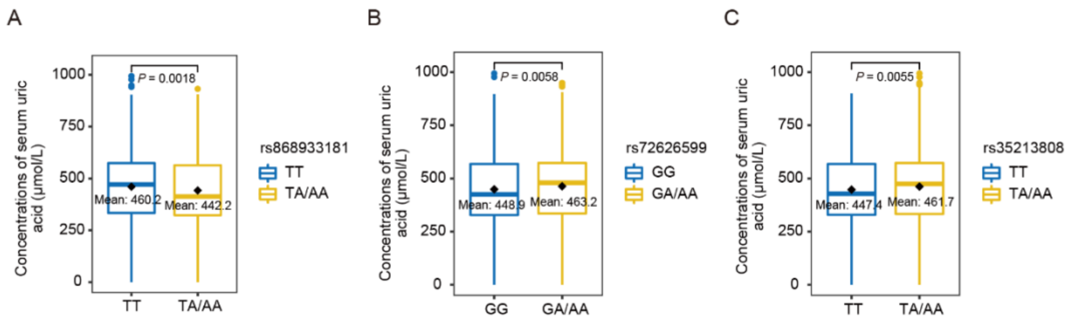

### Appendix Tables

#### Appendix Table 1 SNVs with $P < 1\times10^{-4}$ in the G-NG group of GWAS discovery stage

| | Position | | | | | Frequency of A1 | | | |
|---|---|---|---|---|---|---|---|---|---|
| Rs id | Chr | Bp[a] | A1/A2[b] | Gene | Locus | Cases | Controls | OR (95% CI) | P value |
| rs3737624 | 1 | 1620904 | A/G | *CDK11B - SLC35E2B* | 1p36.33 | 0.543 | 0.47 | 1.64 ( 1.28 - 2.09 ) | 8.74E-05 |
| rs4484948 | 1 | 20041054 | G/A | *TMCO4* | 1p36.13 | 0.911 | 0.834 | 2.26 ( 1.53 - 3.34 ) | 4.46E-05 |
| rs57247989 | 1 | 43414370 | G/C | *SLC2A1* | 1p34.2 | 0.211 | 0.151 | 2.13 ( 1.49 - 3.05 ) | 3.17E-05 |
| rs879688782 | 1 | 52348092 | C/A | *NRDC - RAB3B* | 1p32.3 | 0.795 | 0.727 | 2.03 ( 1.45 - 2.85 ) | 3.54E-05 |
| rs7536714 | 1 | 94381597 | C/G | *GCLM - ABCA4* | 1p22.1 | 0.896 | 0.815 | 2.05 ( 1.43 - 2.94 ) | 8.62E-05 |
| rs116759146 | 1 | 94384955 | T/C | *GCLM - ABCA4* | 1p22.1 | 0.894 | 0.812 | 2.09 ( 1.46 - 2.98 ) | 5.18E-05 |
| rs1473711 | 1 | 94385591 | G/A | *GCLM - ABCA4* | 1p22.1 | 0.894 | 0.812 | 2.09 ( 1.46 - 2.98 ) | 5.18E-05 |
| rs11165034 | 1 | 94387055 | A/G | *GCLM - ABCA4* | 1p22.1 | 0.896 | 0.812 | 2.13 ( 1.49 - 3.05 ) | 3.40E-05 |
| rs2391324 | 1 | 94387597 | C/T | *GCLM - ABCA4* | 1p22.1 | 0.896 | 0.812 | 2.13 ( 1.49 - 3.05 ) | 3.40E-05 |
| rs761889 | 1 | 94387799 | A/G | *GCLM - ABCA4* | 1p22.1 | 0.894 | 0.811 | 2.1 ( 1.47 - 2.99 ) | 4.28E-05 |
| rs2017736 | 1 | 94388005 | T/C | *GCLM - ABCA4* | 1p22.1 | 0.894 | 0.812 | 2.06 ( 1.44 - 2.94 ) | 7.19E-05 |
| rs10874811 | 1 | 94388143 | C/T | *GCLM - ABCA4* | 1p22.1 | 0.894 | 0.812 | 2.09 ( 1.46 - 2.98 ) | 5.18E-05 |
| rs12135991 | 1 | 94389196 | C/T | *GCLM - ABCA4* | 1p22.1 | 0.894 | 0.812 | 2.05 ( 1.43 - 2.93 ) | 7.89E-05 |
| rs6699912 | 1 | 94390174 | A/C | *GCLM - ABCA4* | 1p22.1 | 0.894 | 0.812 | 2.06 ( 1.44 - 2.94 ) | 7.28E-05 |
| rs10874812 | 1 | 94392613 | T/A | *GCLM - ABCA4* | 1p22.1 | 0.894 | 0.812 | 2.09 ( 1.46 - 2.98 ) | 5.18E-05 |
| rs6696758 | 1 | 94394079 | G/A | *GCLM - ABCA4* | 1p22.1 | 0.894 | 0.812 | 2.09 ( 1.46 - 2.98 ) | 5.18E-05 |
| rs10159000 | 1 | 94394420 | G/C | *GCLM - ABCA4* | 1p22.1 | 0.894 | 0.812 | 2.09 ( 1.46 - 2.98 ) | 5.18E-05 |
| rs1989021 | 1 | 94396070 | A/G | *GCLM - ABCA4* | 1p22.1 | 0.894 | 0.811 | 2.1 ( 1.47 - 2.99 ) | 4.07E-05 |
| rs1989020 | 1 | 94396525 | C/A | *GCLM - ABCA4* | 1p22.1 | 0.894 | 0.812 | 2.09 ( 1.46 - 2.98 ) | 5.18E-05 |
| rs114907547 | 1 | 94397001 | G/A | *GCLM - ABCA4* | 1p22.1 | 0.894 | 0.812 | 2.09 ( 1.46 - 2.98 ) | 5.18E-05 |
| rs115601346 | 1 | 94398028 | C/T | *GCLM - ABCA4* | 1p22.1 | 0.896 | 0.814 | 2.09 ( 1.46 - 2.99 ) | 5.97E-05 |
| rs12140097 | 1 | 94398945 | C/T | *GCLM - ABCA4* | 1p22.1 | 0.893 | 0.811 | 2.03 ( 1.42 - 2.89 ) | 8.63E-05 |

| | | | | | | | | | |
|---|---|---|---|---|---|---|---|---|---|
| rs12129986 | 1 | 94399210 | A/G | *GCLM - ABCA4* | 1p22.1 | 0.9 | 0.812 | 2.19 ( 1.53 - 3.14 ) | 2.07E-05 |
| rs10489607 | 1 | 94400446 | T/C | *GCLM - ABCA4* | 1p22.1 | 0.894 | 0.812 | 2.09 ( 1.46 - 2.98 ) | 5.18E-05 |
| rs12131247 | 1 | 94400986 | A/G | *GCLM - ABCA4* | 1p22.1 | 0.894 | 0.812 | 2.09 ( 1.46 - 2.98 ) | 5.18E-05 |
| rs4573532 | 1 | 94405290 | A/C | *GCLM - ABCA4* | 1p22.1 | 0.894 | 0.812 | 2.09 ( 1.46 - 2.98 ) | 5.18E-05 |
| rs1190101854 | 1 | 154971204 | T/* | . | . | 0.821 | 0.804 | 2.07 ( 1.44 - 2.97 ) | 8.40E-05 |
| rs10918897 | 1 | 168438268 | C/T | *LOC101928565* | 1q24.2 | 0.776 | 0.678 | 1.74 ( 1.32 - 2.29 ) | 9.28E-05 |
| rs6698005 | 1 | 221560501 | T/C | *C1orf140 - DUSP10* | 1q41 | 0.298 | 0.21 | 1.9 ( 1.4 - 2.57 ) | 3.12E-05 |
| rs143936649 | 1 | 221561328 | G/A | *C1orf140 - DUSP10* | 1q41 | 0.296 | 0.21 | 1.9 ( 1.41 - 2.57 ) | 2.98E-05 |
| rs1459221 | 1 | 221562934 | G/T | *C1orf140 - DUSP10* | 1q41 | 0.298 | 0.21 | 1.9 ( 1.4 - 2.57 ) | 3.12E-05 |
| rs1551672 | 1 | 221564323 | C/T | *C1orf140 - DUSP10* | 1q41 | 0.298 | 0.21 | 1.9 ( 1.4 - 2.57 ) | 3.12E-05 |
| rs1380250 | 1 | 221566510 | A/G | *C1orf140 - DUSP10* | 1q41 | 0.211 | 0.15 | 2.2 ( 1.53 - 3.15 ) | 1.74E-05 |
| rs755104 | 1 | 221569202 | G/A | *C1orf140 - DUSP10* | 1q41 | 0.298 | 0.209 | 1.93 ( 1.43 - 2.61 ) | 1.88E-05 |
| rs9729478 | 1 | 221569322 | T/C | *C1orf140 - DUSP10* | 1q41 | 0.211 | 0.15 | 2.2 ( 1.53 - 3.15 ) | 1.74E-05 |
| rs12041580 | 1 | 221576856 | C/A | *C1orf140 - DUSP10* | 1q41 | 0.298 | 0.21 | 1.9 ( 1.4 - 2.57 ) | 3.12E-05 |
| rs4550052 | 1 | 221584840 | T/C | *C1orf140 - DUSP10* | 1q41 | 0.298 | 0.21 | 1.9 ( 1.4 - 2.57 ) | 3.12E-05 |
| rs12410403 | 1 | 221587450 | G/A | *C1orf140 - DUSP10* | 1q41 | 0.296 | 0.21 | 1.85 ( 1.37 - 2.49 ) | 5.86E-05 |
| rs1459233 | 1 | 221594969 | G/A | *C1orf140 - DUSP10* | 1q41 | 0.298 | 0.21 | 1.9 ( 1.4 - 2.57 ) | 3.12E-05 |
| rs994606 | 1 | 221602192 | G/T | *C1orf140 - DUSP10* | 1q41 | 0.294 | 0.213 | 1.84 ( 1.37 - 2.48 ) | 6.34E-05 |
| rs9727891 | 1 | 221603164 | G/A | *C1orf140 - DUSP10* | 1q41 | 0.338 | 0.248 | 1.77 ( 1.34 - 2.34 ) | 6.30E-05 |
| rs1031675 | 1 | 221605304 | A/G | *C1orf140 - DUSP10* | 1q41 | 0.296 | 0.214 | 1.84 ( 1.36 - 2.48 ) | 6.58E-05 |
| rs10495163 | 1 | 221605832 | T/A | *C1orf140 - DUSP10* | 1q41 | 0.337 | 0.251 | 1.77 ( 1.33 - 2.36 ) | 7.79E-05 |
| rs12032381 | 1 | 221617076 | C/A | *C1orf140 - DUSP10* | 1q41 | 0.389 | 0.317 | 1.77 ( 1.34 - 2.33 ) | 5.05E-05 |
| rs56321125 | 1 | 226492280 | T/C | *LIN9* | 1q42.12 | 0.715 | 0.636 | 1.79 ( 1.36 - 2.37 ) | 4.25E-05 |
| rs7546310 | 1 | 232061820 | A/C | *DISC1* | 1q42.2 | 0.841 | 0.733 | 1.91 ( 1.4 - 2.61 ) | 4.93E-05 |
| rs1341555 | 1 | 232069057 | T/C | *DISC1* | 1q42.2 | 0.841 | 0.734 | 1.89 ( 1.38 - 2.58 ) | 6.70E-05 |
| rs6697193 | 1 | 249182887 | C/T | *ZNF692 - PGBD2* | 1q44 | 0.824 | 0.762 | 1.86 ( 1.38 - 2.5 ) | 4.38E-05 |

| | | | | | | | | | |
|---|---|---|---|---|---|---|---|---|---|
| rs4335411 | 1 | 249191706 | A/G | *ZNF692 - PGBD2* | 1q44 | 0.809 | 0.748 | 1.88 ( 1.4 - 2.52 ) | 2.42E-05 |
| rs6731724 | 2 | 29546758 | G/T | *ALK* | 2p23.2 | 0.387 | 0.315 | 1.73 ( 1.33 - 2.26 ) | 5.32E-05 |
| rs1212605325 | 2 | 37562188 | */A | . | . | 0.916 | 0.847 | 2.39 ( 1.57 - 3.65 ) | 4.88E-05 |
| rs74613934 | 2 | 167198680 | G/A | *SCN9A* | 2q24.3 | 0.969 | 0.927 | 3.45 ( 1.85 - 6.43 ) | 9.54E-05 |
| rs79684957 | 2 | 167200980 | A/C | *SCN9A* | 2q24.3 | 0.969 | 0.927 | 3.45 ( 1.85 - 6.43 ) | 9.54E-05 |
| rs77154470 | 2 | 167212046 | C/T | *SCN9A* | 2q24.3 | 0.952 | 0.894 | 2.98 ( 1.79 - 4.94 ) | 2.40E-05 |
| rs148362057 | 2 | 167232446 | A/G | *SCN9A* | 2q24.3 | 0.959 | 0.918 | 3 ( 1.73 - 5.22 ) | 9.78E-05 |
| rs140991639 | 2 | 226294958 | A/G | *NYAP2* | 2q36.3 | 0.078 | 0.039 | 3.94 ( 2.02 - 7.68 ) | 5.69E-05 |
| rs11917223 | 3 | 39323423 | C/G | *CX3CR1* | 3p22.2 | 0.409 | 0.335 | 1.7 ( 1.3 - 2.23 ) | 9.15E-05 |
| rs868933181 | 3 | 51519910 | T/A | *VPRBP* | 3p21.2 | 0.918 | 0.89 | 2.46 ( 1.61 - 3.77 ) | 3.41E-05 |
| rs603059 | 3 | 54639970 | A/G | *CACNA2D3* | 3p14.3 | 0.606 | 0.506 | 1.67 ( 1.3 - 2.14 ) | 4.58E-05 |
| rs717228 | 3 | 60602895 | T/C | *FHIT* | 3p14.2 | 0.544 | 0.485 | 1.76 ( 1.36 - 2.29 ) | 2.34E-05 |
| rs2660768 | 3 | 87079981 | C/T | *VGLL3 - LINC00506* | 3p12.1 | 0.943 | 0.901 | 2.56 ( 1.6 - 4.09 ) | 8.83E-05 |
| rs1461274 | 3 | 174874317 | G/A | *NAALADL2* | 3q26.31 | 0.872 | 0.789 | 1.93 ( 1.39 - 2.68 ) | 9.50E-05 |
| rs13320132 | 3 | 174874373 | T/C | *NAALADL2* | 3q26.31 | 0.872 | 0.789 | 1.93 ( 1.39 - 2.68 ) | 9.50E-05 |
| rs1461273 | 3 | 174874389 | G/A | *NAALADL2* | 3q26.31 | 0.872 | 0.789 | 1.93 ( 1.39 - 2.68 ) | 9.50E-05 |
| rs13323546 | 3 | 174874555 | A/C | *NAALADL2* | 3q26.31 | 0.872 | 0.789 | 1.93 ( 1.39 - 2.68 ) | 9.50E-05 |
| rs9860693 | 3 | 174874634 | A/G | *NAALADL2* | 3q26.31 | 0.872 | 0.789 | 1.93 ( 1.39 - 2.68 ) | 9.50E-05 |
| rs9290541 | 3 | 174874675 | G/A | *NAALADL2* | 3q26.31 | 0.872 | 0.789 | 1.93 ( 1.39 - 2.68 ) | 9.50E-05 |
| rs9860857 | 3 | 174874709 | A/C | *NAALADL2* | 3q26.31 | 0.872 | 0.789 | 1.93 ( 1.39 - 2.68 ) | 9.50E-05 |
| rs2861997 | 3 | 174875574 | A/G | *NAALADL2* | 3q26.31 | 0.872 | 0.789 | 1.95 ( 1.4 - 2.71 ) | 7.83E-05 |
| rs73041397 | 3 | 174875925 | A/G | *NAALADL2* | 3q26.31 | 0.874 | 0.789 | 1.98 ( 1.42 - 2.75 ) | 5.74E-05 |
| rs11921295 | 3 | 174876429 | A/T | *NAALADL2* | 3q26.31 | 0.872 | 0.789 | 1.95 ( 1.4 - 2.71 ) | 7.83E-05 |
| rs10470502 | 3 | 174876464 | A/G | *NAALADL2* | 3q26.31 | 0.872 | 0.788 | 1.95 ( 1.4 - 2.71 ) | 7.83E-05 |
| rs9858268 | 3 | 174879055 | A/G | *NAALADL2* | 3q26.31 | 0.874 | 0.788 | 1.98 ( 1.42 - 2.75 ) | 5.74E-05 |
| rs1381132 | 3 | 174896492 | T/C | *NAALADL2* | 3q26.31 | 0.852 | 0.763 | 1.87 ( 1.37 - 2.56 ) | 9.18E-05 |

| | | | | | | | | | |
|---|---|---|---|---|---|---|---|---|---|
| rs2861994 | 3 | 174896751 | C/A | *NAALADL2* | 3q26.31 | 0.854 | 0.764 | 1.87 ( 1.37 - 2.57 ) | 8.99E-05 |
| rs12638990 | 3 | 174896874 | C/T | *NAALADL2* | 3q26.31 | 0.852 | 0.763 | 1.87 ( 1.37 - 2.56 ) | 9.18E-05 |
| rs11712270 | 3 | 193714121 | A/C | *LINC02026* | 3q29 | 0.794 | 0.72 | 1.76 ( 1.33 - 2.34 ) | 7.73E-05 |
| rs35594721 | 3 | 193717926 | T/C | *LINC02026* | 3q29 | 0.793 | 0.718 | 1.74 ( 1.32 - 2.31 ) | 1.00E-04 |
| rs76621524 | 4 | 69888130 | T/G | *UGT2B10 - UGT2B7* | 4q13.2 | 0.824 | 0.737 | 1.83 ( 1.36 - 2.48 ) | 8.12E-05 |
| rs11730928 | 4 | 69957915 | C/T | *UGT2B7* | 4q13.2 | 0.824 | 0.736 | 1.85 ( 1.37 - 2.5 ) | 6.81E-05 |
| rs6837285 | 4 | 70006617 | G/T | *UGT2B7 - LOC105377267* | 4q13.2 | 0.807 | 0.716 | 1.78 ( 1.33 - 2.38 ) | 9.87E-05 |
| rs2227306 | 4 | 74607055 | T/C | *CXCL8* | 4q13.3 | 0.422 | 0.351 | 1.78 ( 1.36 - 2.34 ) | 2.72E-05 |
| rs10938092 | 4 | 74609715 | A/G | *CXCL8* | 4q13.3 | 0.42 | 0.354 | 1.74 ( 1.32 - 2.27 ) | 6.82E-05 |
| rs6532039 | 4 | 88900362 | G/A | *SPP1* | 4q22.1 | 0.428 | 0.29 | 1.76 ( 1.37 - 2.28 ) | 1.15E-05 |
| rs1126616 | 4 | 88903853 | C/T | *SPP1* | 4q22.1 | 0.417 | 0.291 | 1.7 ( 1.3 - 2.21 ) | 9.45E-05 |
| rs7655182 | 4 | 88905903 | G/A | *SPP1 - PKD2* | 4q22.1 | 0.654 | 0.508 | 1.68 ( 1.31 - 2.16 ) | 4.47E-05 |
| rs4345141 | 4 | 88913121 | A/G | *SPP1 - PKD2* | 4q22.1 | 0.376 | 0.234 | 1.79 ( 1.37 - 2.35 ) | 2.41E-05 |
| rs56917667 | 4 | 88913938 | C/T | *SPP1 - PKD2* | 4q22.1 | 0.417 | 0.288 | 1.73 ( 1.32 - 2.25 ) | 5.80E-05 |
| rs58408285 | 4 | 88913941 | T/G | *SPP1 - PKD2* | 4q22.1 | 0.417 | 0.289 | 1.7 ( 1.31 - 2.22 ) | 8.36E-05 |
| rs60249214 | 4 | 88913954 | C/T | *SPP1 - PKD2* | 4q22.1 | 0.417 | 0.289 | 1.7 ( 1.31 - 2.22 ) | 8.36E-05 |
| rs57929326 | 4 | 88913999 | C/T | *SPP1 - PKD2* | 4q22.1 | 0.417 | 0.289 | 1.7 ( 1.31 - 2.22 ) | 8.36E-05 |
| rs116863899 | 4 | 88914319 | G/A | *SPP1 - PKD2* | 4q22.1 | 0.733 | 0.588 | 1.75 ( 1.35 - 2.27 ) | 2.51E-05 |
| rs13137113 | 4 | 88914436 | C/T | *SPP1 - PKD2* | 4q22.1 | 0.44 | 0.3 | 1.78 ( 1.35 - 2.37 ) | 5.70E-05 |
| rs4376106 | 4 | 88914666 | A/G | *SPP1 - PKD2* | 4q22.1 | 0.418 | 0.29 | 1.69 ( 1.3 - 2.19 ) | 9.57E-05 |
| rs4282132 | 4 | 88914780 | C/T | *SPP1 - PKD2* | 4q22.1 | 0.417 | 0.289 | 1.7 ( 1.31 - 2.22 ) | 8.36E-05 |
| rs72871575 | 4 | 88915273 | T/C | *SPP1 - PKD2* | 4q22.1 | 0.711 | 0.572 | 1.74 ( 1.34 - 2.26 ) | 3.06E-05 |
| rs72871576 | 4 | 88915288 | T/G | *SPP1 - PKD2* | 4q22.1 | 0.711 | 0.572 | 1.74 ( 1.34 - 2.26 ) | 3.06E-05 |
| rs72871579 | 4 | 88915612 | T/C | *SPP1 - PKD2* | 4q22.1 | 0.713 | 0.572 | 1.76 ( 1.35 - 2.28 ) | 2.39E-05 |
| rs78099378 | 4 | 88915731 | A/C | *SPP1 - PKD2* | 4q22.1 | 0.711 | 0.571 | 1.76 ( 1.35 - 2.28 ) | 2.35E-05 |
| rs72871581 | 4 | 88915780 | G/A | *SPP1 - PKD2* | 4q22.1 | 0.711 | 0.571 | 1.76 ( 1.35 - 2.28 ) | 2.35E-05 |

| | | | | | | | | | |
|---|---|---|---|---|---|---|---|---|---|
| rs72871583 | 4 | 88916031 | C/T | *SPP1 - PKD2* | 4q22.1 | 0.711 | 0.571 | 1.76 ( 1.35 - 2.28 ) | 2.35E-05 |
| rs72871585 | 4 | 88916130 | C/T | *SPP1 - PKD2* | 4q22.1 | 0.711 | 0.571 | 1.76 ( 1.35 - 2.28 ) | 2.35E-05 |
| rs12650232 | 4 | 88916501 | T/G | *SPP1 - PKD2* | 4q22.1 | 0.711 | 0.571 | 1.76 ( 1.35 - 2.28 ) | 2.35E-05 |
| rs17013702 | 4 | 88916715 | A/G | *SPP1 - PKD2* | 4q22.1 | 0.707 | 0.57 | 1.71 ( 1.32 - 2.21 ) | 4.34E-05 |
| rs12647183 | 4 | 88917078 | G/C | *SPP1 - PKD2* | 4q22.1 | 0.711 | 0.572 | 1.72 ( 1.33 - 2.23 ) | 4.13E-05 |
| rs12641146 | 4 | 88917266 | A/T | *SPP1 - PKD2* | 4q22.1 | 0.711 | 0.574 | 1.74 ( 1.34 - 2.26 ) | 2.90E-05 |
| rs72871588 | 4 | 88917447 | G/T | *SPP1 - PKD2* | 4q22.1 | 0.711 | 0.571 | 1.74 ( 1.34 - 2.26 ) | 3.15E-05 |
| rs76158295 | 4 | 88917535 | C/T | *SPP1 - PKD2* | 4q22.1 | 0.711 | 0.571 | 1.76 ( 1.35 - 2.28 ) | 2.35E-05 |
| rs76923321 | 4 | 88917723 | C/T | *SPP1 - PKD2* | 4q22.1 | 0.711 | 0.57 | 1.76 ( 1.36 - 2.29 ) | 1.93E-05 |
| rs17013705 | 4 | 88917735 | T/C | *SPP1 - PKD2* | 4q22.1 | 0.711 | 0.571 | 1.76 ( 1.35 - 2.28 ) | 2.35E-05 |
| rs17013707 | 4 | 88917836 | C/T | *SPP1 - PKD2* | 4q22.1 | 0.709 | 0.572 | 1.7 ( 1.31 - 2.2 ) | 5.92E-05 |
| rs2725237 | 4 | 88918786 | T/C | *SPP1 - PKD2* | 4q22.1 | 0.376 | 0.233 | 1.81 ( 1.38 - 2.38 ) | 1.93E-05 |
| rs77972559 | 4 | 88919101 | T/C | *SPP1 - PKD2* | 4q22.1 | 0.71 | 0.572 | 1.73 ( 1.33 - 2.24 ) | 3.71E-05 |
| rs59961674 | 4 | 88919362 | C/G | *SPP1 - PKD2* | 4q22.1 | 0.715 | 0.571 | 1.8 ( 1.38 - 2.33 ) | 1.21E-05 |
| rs57588100 | 4 | 88919528 | T/A | *SPP1 - PKD2* | 4q22.1 | 0.711 | 0.571 | 1.76 ( 1.35 - 2.28 ) | 2.35E-05 |
| rs57009586 | 4 | 88919536 | A/G | *SPP1 - PKD2* | 4q22.1 | 0.711 | 0.571 | 1.76 ( 1.35 - 2.28 ) | 2.35E-05 |
| rs56983082 | 4 | 88919548 | T/C | *SPP1 - PKD2* | 4q22.1 | 0.713 | 0.571 | 1.77 ( 1.37 - 2.3 ) | 1.72E-05 |
| rs61580992 | 4 | 88919572 | G/A | *SPP1 - PKD2* | 4q22.1 | 0.715 | 0.571 | 1.79 ( 1.38 - 2.33 ) | 1.20E-05 |
| rs57007910 | 4 | 88919651 | T/A | *SPP1 - PKD2* | 4q22.1 | 0.711 | 0.571 | 1.74 ( 1.34 - 2.25 ) | 3.21E-05 |
| rs12649741 | 4 | 88920124 | C/T | *SPP1 - PKD2* | 4q22.1 | 0.711 | 0.571 | 1.76 ( 1.35 - 2.28 ) | 2.35E-05 |
| rs12643720 | 4 | 88920172 | A/G | *SPP1 - PKD2* | 4q22.1 | 0.711 | 0.572 | 1.76 ( 1.35 - 2.28 ) | 2.35E-05 |
| rs148294817 | 4 | 88920508 | G/T | *SPP1 - PKD2* | 4q22.1 | 0.713 | 0.571 | 1.77 ( 1.36 - 2.29 ) | 1.91E-05 |
| rs28674686 | 4 | 88921277 | A/G | *SPP1 - PKD2* | 4q22.1 | 0.713 | 0.576 | 1.76 ( 1.36 - 2.29 ) | 1.97E-05 |
| rs72873405 | 4 | 88921369 | G/C | *SPP1 - PKD2* | 4q22.1 | 0.711 | 0.571 | 1.76 ( 1.35 - 2.28 ) | 2.35E-05 |
| rs72873408 | 4 | 88921446 | G/C | *SPP1 - PKD2* | 4q22.1 | 0.711 | 0.571 | 1.76 ( 1.35 - 2.28 ) | 2.35E-05 |
| rs72873410 | 4 | 88921512 | G/A | *SPP1 - PKD2* | 4q22.1 | 0.711 | 0.571 | 1.76 ( 1.35 - 2.28 ) | 2.35E-05 |

| | | | | | | | | | |
|---|---|---|---|---|---|---|---|---|---|
| rs60179294 | 4 | 88921927 | A/G | *SPP1 - PKD2* | 4q22.1 | 0.713 | 0.571 | 1.77 ( 1.37 - 2.3 ) | 1.69E-05 |
| rs61108048 | 4 | 88922391 | C/T | *SPP1 - PKD2* | 4q22.1 | 0.711 | 0.571 | 1.76 ( 1.35 - 2.28 ) | 2.35E-05 |
| rs80083821 | 4 | 88922832 | C/T | *SPP1 - PKD2* | 4q22.1 | 0.711 | 0.57 | 1.77 ( 1.37 - 2.31 ) | 1.73E-05 |
| rs79080030 | 4 | 88922897 | A/G | *SPP1 - PKD2* | 4q22.1 | 0.711 | 0.572 | 1.74 ( 1.34 - 2.25 ) | 3.26E-05 |
| rs61566498 | 4 | 88922968 | C/T | *SPP1 - PKD2* | 4q22.1 | 0.713 | 0.572 | 1.77 ( 1.36 - 2.3 ) | 1.82E-05 |
| rs60515196 | 4 | 88923093 | T/A | *SPP1 - PKD2* | 4q22.1 | 0.711 | 0.57 | 1.77 ( 1.36 - 2.3 ) | 1.86E-05 |
| rs2728131 | 4 | 88924344 | T/C | *SPP1 - PKD2* | 4q22.1 | 0.374 | 0.23 | 1.82 ( 1.38 - 2.39 ) | 1.79E-05 |
| rs2725234 | 4 | 88932272 | T/C | *PKD2* | 4q22.1 | 0.382 | 0.23 | 1.89 ( 1.44 - 2.48 ) | 4.97E-06 |
| rs74970203 | 4 | 88938067 | A/G | *PKD2* | 4q22.1 | 0.702 | 0.569 | 1.68 ( 1.3 - 2.18 ) | 7.94E-05 |
| rs79485111 | 4 | 88938074 | T/C | *PKD2* | 4q22.1 | 0.702 | 0.569 | 1.68 ( 1.3 - 2.18 ) | 7.94E-05 |
| rs4484262 | 4 | 88939000 | C/A | *PKD2* | 4q22.1 | 0.702 | 0.567 | 1.73 ( 1.33 - 2.25 ) | 4.07E-05 |
| rs2728113 | 4 | 88939736 | G/A | *PKD2* | 4q22.1 | 0.391 | 0.231 | 1.98 ( 1.5 - 2.61 ) | 1.18E-06 |
| rs10516801 | 4 | 88941549 | C/T | *PKD2* | 4q22.1 | 0.659 | 0.52 | 1.66 ( 1.29 - 2.14 ) | 9.04E-05 |
| rs7700163 | 4 | 88942584 | G/T | *PKD2* | 4q22.1 | 0.659 | 0.52 | 1.66 ( 1.29 - 2.14 ) | 9.04E-05 |
| rs4312723 | 4 | 88947869 | G/A | *PKD2* | 4q22.1 | 0.663 | 0.521 | 1.68 ( 1.31 - 2.17 ) | 5.76E-05 |
| rs6850829 | 4 | 88954247 | G/C | *PKD2* | 4q22.1 | 0.661 | 0.52 | 1.67 ( 1.3 - 2.16 ) | 6.97E-05 |
| rs13119877 | 4 | 88954408 | G/A | *PKD2* | 4q22.1 | 0.661 | 0.52 | 1.67 ( 1.3 - 2.15 ) | 7.23E-05 |
| rs28634790 | 4 | 88954523 | A/G | *PKD2* | 4q22.1 | 0.393 | 0.231 | 2 ( 1.52 - 2.64 ) | 8.26E-07 |
| rs60621874 | 4 | 88955603 | C/G | *PKD2* | 4q22.1 | 0.7 | 0.565 | 1.74 ( 1.34 - 2.26 ) | 3.38E-05 |
| rs6532045 | 4 | 88956521 | A/G | *PKD2* | 4q22.1 | 0.663 | 0.52 | 1.68 ( 1.31 - 2.17 ) | 5.77E-05 |
| rs2728109 | 4 | 88957723 | A/C | *PKD2* | 4q22.1 | 0.387 | 0.218 | 2.04 ( 1.54 - 2.7 ) | 6.12E-07 |
| rs2725222 | 4 | 88958492 | A/G | *PKD2* | 4q22.1 | 0.398 | 0.225 | 2.08 ( 1.58 - 2.75 ) | 2.55E-07 |
| rs17013735 | 4 | 88959745 | A/G | *PKD2* | 4q22.1 | 0.711 | 0.569 | 1.68 ( 1.3 - 2.16 ) | 5.89E-05 |
| rs2725220 | 4 | 88959922 | C/G | *PKD2* | 4q22.1 | 0.42 | 0.223 | 2.27 ( 1.72 - 3 ) | 9.24E-09 |
| rs2725219 | 4 | 88959961 | C/T | *PKD2* | 4q22.1 | 0.42 | 0.218 | 2.34 ( 1.76 - 3.09 ) | 3.18E-09 |
| rs2725218 | 4 | 88959968 | C/G | *PKD2* | 4q22.1 | 0.422 | 0.224 | 2.28 ( 1.72 - 3.01 ) | 7.44E-09 |

| | | | | | | | | | |
|---|---|---|---|---|---|---|---|---|---|
| rs2725217 | 4 | 88960258 | T/A | *PKD2* | 4q22.1 | 0.428 | 0.218 | 2.43 ( 1.83 - 3.22 ) | 8.38E-10 |
| rs2725216 | 4 | 88960579 | G/A | *PKD2* | 4q22.1 | 0.437 | 0.246 | 2.1 ( 1.59 - 2.77 ) | 1.50E-07 |
| rs2725215 | 4 | 88961571 | T/C | *PKD2* | 4q22.1 | 0.43 | 0.218 | 2.46 ( 1.85 - 3.26 ) | 5.66E-10 |
| rs2728108 | 4 | 88961736 | C/A | *PKD2* | 4q22.1 | 0.437 | 0.246 | 2.1 ( 1.59 - 2.77 ) | 1.50E-07 |
| rs17013739 | 4 | 88963255 | G/A | *PKD2* | 4q22.1 | 0.904 | 0.765 | 2.68 ( 1.84 - 3.91 ) | 2.79E-07 |
| rs10031265 | 4 | 88968343 | A/G | *PKD2* | 4q22.1 | 0.898 | 0.724 | 3.24 ( 2.26 - 4.65 ) | 1.50E-10 |
| rs2728107 | 4 | 88968488 | T/C | *PKD2* | 4q22.1 | 0.435 | 0.22 | 2.54 ( 1.91 - 3.38 ) | 1.77E-10 |
| rs2725211 | 4 | 88970375 | T/C | *PKD2* | 4q22.1 | 0.435 | 0.22 | 2.54 ( 1.91 - 3.38 ) | 1.77E-10 |
| rs2728104 | 4 | 88973006 | C/T | *PKD2* | 4q22.1 | 0.441 | 0.243 | 2.22 ( 1.68 - 2.93 ) | 2.51E-08 |
| rs2728099 | 4 | 88975738 | C/T | *PKD2* | 4q22.1 | 0.433 | 0.217 | 2.47 ( 1.86 - 3.28 ) | 4.11E-10 |
| rs2728134 | 4 | 88978139 | G/T | *PKD2* | 4q22.1 | 0.433 | 0.219 | 2.48 ( 1.87 - 3.3 ) | 3.37E-10 |
| rs74901820 | 4 | 88981537 | G/A | *PKD2* | 4q22.1 | 0.906 | 0.771 | 2.62 ( 1.79 - 3.83 ) | 6.50E-07 |
| rs74668466 | 4 | 88981837 | A/G | *PKD2* | 4q22.1 | 0.906 | 0.764 | 2.69 ( 1.84 - 3.92 ) | 3.18E-07 |
| rs117007073 | 4 | 88987915 | C/T | *PKD2* | 4q22.1 | 0.906 | 0.761 | 2.77 ( 1.89 - 4.04 ) | 1.42E-07 |
| rs74933714 | 4 | 88990386 | T/G | *PKD2* | 4q22.1 | 0.906 | 0.77 | 2.62 ( 1.8 - 3.82 ) | 4.75E-07 |
| rs2725204 | 4 | 88992813 | G/T | *PKD2* | 4q22.1 | 0.437 | 0.221 | 2.43 ( 1.83 - 3.21 ) | 6.82E-10 |
| rs2725202 | 4 | 88997876 | A/G | *PKD2* | 4q22.1 | 0.437 | 0.22 | 2.48 ( 1.87 - 3.29 ) | 3.42E-10 |
| rs2728126 | 4 | 88999222 | A/T | *PKD2* | 4q22.1 | 0.443 | 0.221 | 2.5 ( 1.88 - 3.31 ) | 2.21E-10 |
| rs17013802 | 4 | 89001185 | T/A | *PKD2 - ABCG2* | 4q22.1 | 0.904 | 0.77 | 2.6 ( 1.79 - 3.79 ) | 5.94E-07 |
| rs2728125 | 4 | 89001893 | G/A | *PKD2 - ABCG2* | 4q22.1 | 0.443 | 0.223 | 2.5 ( 1.88 - 3.32 ) | 2.56E-10 |
| rs2728124 | 4 | 89006160 | A/T | *PKD2 - ABCG2* | 4q22.1 | 0.456 | 0.242 | 2.38 ( 1.8 - 3.14 ) | 9.83E-10 |
| rs17013810 | 4 | 89008506 | A/G | *PKD2 - ABCG2* | 4q22.1 | 0.909 | 0.766 | 2.9 ( 1.97 - 4.27 ) | 6.33E-08 |
| rs2725269 | 4 | 89009006 | T/C | *PKD2 - ABCG2* | 4q22.1 | 0.435 | 0.234 | 2.23 ( 1.7 - 2.93 ) | 5.89E-09 |
| rs142474948 | 4 | 89010900 | A/G | *ABCG2* | 4q22.1 | 0.915 | 0.78 | 2.93 ( 1.98 - 4.34 ) | 8.28E-08 |
| rs2725268 | 4 | 89010983 | G/A | *ABCG2* | 4q22.1 | 0.444 | 0.236 | 2.27 ( 1.73 - 2.99 ) | 3.89E-09 |
| rs1448784 | 4 | 89012320 | A/G | *ABCG2* | 4q22.1 | 0.878 | 0.718 | 2.59 ( 1.83 - 3.66 ) | 6.71E-08 |

| | | | | | | | | | |
|---|---|---|---|---|---|---|---|---|---|
| rs4148160 | 4 | 89015090 | C/T | *ABCG2* | 4q22.1 | 0.909 | 0.76 | 3.13 ( 2.11 - 4.66 ) | 1.56E-08 |
| rs2231164 | 4 | 89015857 | C/T | *ABCG2* | 4q22.1 | 0.646 | 0.463 | 1.95 ( 1.48 - 2.57 ) | 2.02E-06 |
| rs1383585 | 4 | 89019735 | G/A | *ABCG2* | 4q22.1 | 0.472 | 0.228 | 2.75 ( 2.08 - 3.64 ) | 1.06E-12 |
| rs2231156 | 4 | 89020427 | A/C | *ABCG2* | 4q22.1 | 0.472 | 0.238 | 2.62 ( 1.98 - 3.46 ) | 1.46E-11 |
| rs4148157 | 4 | 89020934 | A/G | *ABCG2* | 4q22.1 | 0.472 | 0.238 | 2.62 ( 1.98 - 3.46 ) | 1.46E-11 |
| rs4693924 | 4 | 89023224 | A/G | *ABCG2* | 4q22.1 | 0.472 | 0.238 | 2.62 ( 1.98 - 3.46 ) | 1.46E-11 |
| rs34455506 | 4 | 89024220 | G/A | *ABCG2* | 4q22.1 | 0.909 | 0.759 | 3.13 ( 2.11 - 4.66 ) | 1.56E-08 |
| rs76979899 | 4 | 89025241 | T/C | *ABCG2* | 4q22.1 | 0.47 | 0.235 | 2.62 ( 1.99 - 3.47 ) | 1.17E-11 |
| rs2725263 | 4 | 89026428 | C/A | *ABCG2* | 4q22.1 | 0.737 | 0.537 | 2.36 ( 1.8 - 3.09 ) | 3.94E-10 |
| rs7681519 | 4 | 89027840 | G/C | *ABCG2* | 4q22.1 | 0.909 | 0.759 | 3.13 ( 2.11 - 4.66 ) | 1.56E-08 |
| rs2231148 | 4 | 89028478 | T/A | *ABCG2* | 4q22.1 | 0.909 | 0.76 | 3.13 ( 2.11 - 4.66 ) | 1.56E-08 |
| rs2054576 | 4 | 89028775 | G/A | *ABCG2* | 4q22.1 | 0.47 | 0.238 | 2.59 ( 1.96 - 3.42 ) | 2.11E-11 |
| rs34472643 | 4 | 89029866 | G/A | *ABCG2* | 4q22.1 | 0.894 | 0.745 | 2.81 ( 1.93 - 4.1 ) | 7.72E-08 |
| rs12505410 | 4 | 89030841 | T/G | *ABCG2* | 4q22.1 | 0.876 | 0.67 | 3.21 ( 2.29 - 4.49 ) | 1.00E-11 |
| rs2622621 | 4 | 89030920 | G/C | *ABCG2* | 4q22.1 | 0.778 | 0.563 | 2.63 ( 1.98 - 3.49 ) | 2.61E-11 |
| rs11943824 | 4 | 89031519 | A/G | *ABCG2* | 4q22.1 | 0.876 | 0.674 | 3.07 ( 2.2 - 4.29 ) | 4.05E-11 |
| rs11939579 | 4 | 89031586 | T/G | *ABCG2* | 4q22.1 | 0.876 | 0.674 | 3.08 ( 2.21 - 4.3 ) | 3.89E-11 |
| rs200184409 | 4 | 89031978 | T/* | . | . | 0.856 | 0.75 | 2.03 ( 1.45 - 2.84 ) | 3.23E-05 |
| rs1481013 | 4 | 89032679 | C/T | *ABCG2* | 4q22.1 | 0.818 | 0.616 | 2.78 ( 2.05 - 3.76 ) | 4.89E-11 |
| rs2725261 | 4 | 89036353 | T/C | *ABCG2* | 4q22.1 | 0.786 | 0.57 | 2.47 ( 1.89 - 3.24 ) | 5.56E-11 |
| rs1481012 | 4 | 89039082 | G/A | *ABCG2* | 4q22.1 | 0.598 | 0.284 | 3.92 ( 2.89 - 5.33 ) | 2.49E-18 |
| rs2231146 | 4 | 89039500 | T/C | *ABCG2* | 4q22.1 | 0.93 | 0.789 | 3.39 ( 2.23 - 5.16 ) | 1.03E-08 |
| rs2231145 | 4 | 89039584 | T/C | *ABCG2* | 4q22.1 | 0.93 | 0.789 | 3.39 ( 2.23 - 5.16 ) | 1.03E-08 |
| rs1871744 | 4 | 89039629 | T/C | *ABCG2* | 4q22.1 | 0.9 | 0.72 | 3.32 ( 2.3 - 4.79 ) | 1.65E-10 |
| rs2448795 | 4 | 89040850 | A/G | *ABCG2* | 4q22.1 | 0.83 | 0.702 | 2.06 ( 1.52 - 2.79 ) | 3.27E-06 |
| rs117195876 | 4 | 89041399 | T/C | *ABCG2* | 4q22.1 | 0.93 | 0.789 | 3.39 ( 2.23 - 5.16 ) | 1.03E-08 |

| | | | | | | | | | |
|---|---|---|---|---|---|---|---|---|---|
| rs117815474 | 4 | 89041878 | C/T | *ABCG2* | 4q22.1 | 0.93 | 0.789 | 3.39 ( 2.23 - 5.16 ) | 1.03E-08 |
| rs45499402 | 4 | 89043634 | C/G | *ABCG2* | 4q22.1 | 0.613 | 0.29 | 4.11 ( 3.02 - 5.59 ) | 2.30E-19 |
| rs2170290 | 4 | 89044067 | T/C | *ABCG2* | 4q22.1 | 0.9 | 0.72 | 3.32 ( 2.3 - 4.79 ) | 1.65E-10 |
| rs149027545 | 4 | 89044180 | C/G | *ABCG2* | 4q22.1 | 0.613 | 0.29 | 4.09 ( 3 - 5.56 ) | 3.30E-19 |
| rs138409370 | 4 | 89044312 | T/A | *ABCG2* | 4q22.1 | 0.613 | 0.29 | 4.09 ( 3 - 5.56 ) | 3.30E-19 |
| rs75544042 | 4 | 89045331 | A/G | *ABCG2* | 4q22.1 | 0.613 | 0.291 | 4.09 ( 3 - 5.56 ) | 3.30E-19 |
| rs3102038 | 4 | 89045426 | G/A | *ABCG2* | 4q22.1 | 0.83 | 0.702 | 2.08 ( 1.54 - 2.82 ) | 2.34E-06 |
| rs141471965 | 4 | 89046202 | T/C | *ABCG2* | 4q22.1 | 0.613 | 0.29 | 4.09 ( 3 - 5.56 ) | 3.30E-19 |
| rs117607915 | 4 | 89046617 | G/A | *ABCG2* | 4q22.1 | 0.931 | 0.791 | 3.26 ( 2.14 - 4.96 ) | 3.95E-08 |
| rs74904971 | 4 | 89050026 | A/C | *ABCG2* | 4q22.1 | 0.613 | 0.291 | 4.07 ( 2.99 - 5.53 ) | 4.36E-19 |
| rs2725256 | 4 | 89050998 | A/G | *ABCG2* | 4q22.1 | 0.83 | 0.703 | 2.08 ( 1.54 - 2.82 ) | 2.34E-06 |
| rs2231142 | 4 | 89052323 | T/G | *ABCG2* | 4q22.1 | 0.613 | 0.29 | 4.09 ( 3 - 5.56 ) | 3.30E-19 |
| rs4148155 | 4 | 89054667 | G/A | *ABCG2* | 4q22.1 | 0.611 | 0.29 | 4.02 ( 2.96 - 5.46 ) | 5.14E-19 |
| rs3114017 | 4 | 89055194 | C/T | *ABCG2* | 4q22.1 | 0.824 | 0.692 | 2.09 ( 1.55 - 2.81 ) | 1.56E-06 |
| rs4148153 | 4 | 89056715 | G/A | *ABCG2* | 4q22.1 | 0.928 | 0.779 | 3.46 ( 2.28 - 5.23 ) | 4.48E-09 |
| rs2725254 | 4 | 89057664 | C/T | *ABCG2* | 4q22.1 | 0.832 | 0.705 | 2.08 ( 1.54 - 2.82 ) | 2.38E-06 |
| rs2929060 | 4 | 89058220 | A/C | *ABCG2* | 4q22.1 | 0.832 | 0.704 | 2.06 ( 1.52 - 2.78 ) | 2.92E-06 |
| rs12641369 | 4 | 89059917 | G/A | *ABCG2* | 4q22.1 | 0.811 | 0.652 | 2.04 ( 1.53 - 2.72 ) | 1.34E-06 |
| rs4148152 | 4 | 89060909 | T/C | *ABCG2* | 4q22.1 | 0.817 | 0.667 | 2.01 ( 1.51 - 2.69 ) | 2.06E-06 |
| rs2231137 | 4 | 89061114 | C/T | *ABCG2* | 4q22.1 | 0.818 | 0.668 | 2.03 ( 1.52 - 2.71 ) | 1.60E-06 |
| rs1564481 | 4 | 89061265 | C/T | *ABCG2* | 4q22.1 | 0.83 | 0.704 | 2.08 ( 1.54 - 2.82 ) | 2.29E-06 |
| rs77377473 | 4 | 89061802 | A/G | *ABCG2* | 4q22.1 | 0.937 | 0.805 | 3.47 ( 2.24 - 5.38 ) | 2.32E-08 |
| rs2725252 | 4 | 89061910 | C/A | *ABCG2* | 4q22.1 | 0.793 | 0.613 | 2.37 ( 1.77 - 3.17 ) | 5.70E-09 |
| rs4148150 | 4 | 89062189 | C/T | *ABCG2* | 4q22.1 | 0.826 | 0.693 | 1.95 ( 1.45 - 2.63 ) | 1.05E-05 |
| rs4148149 | 4 | 89062285 | T/G | *ABCG2* | 4q22.1 | 0.82 | 0.674 | 1.99 ( 1.48 - 2.67 ) | 5.08E-06 |
| rs72554039 | 4 | 89063354 | G/A | *ABCG2* | 4q22.1 | 0.937 | 0.805 | 3.4 ( 2.2 - 5.26 ) | 3.54E-08 |

| | | | | | | | | | |
|---|---|---|---|---|---|---|---|---|---|
| rs2622620 | 4 | 89063851 | A/C | *ABCG2* | 4q22.1 | 0.789 | 0.61 | 2.12 ( 1.59 - 2.81 ) | 2.30E-07 |
| rs2725250 | 4 | 89063985 | A/G | *ABCG2* | 4q22.1 | 0.815 | 0.674 | 1.93 ( 1.45 - 2.57 ) | 6.60E-06 |
| rs3114018 | 4 | 89064581 | C/A | *ABCG2* | 4q22.1 | 0.787 | 0.619 | 2.05 ( 1.54 - 2.73 ) | 7.33E-07 |
| rs3109823 | 4 | 89064602 | T/C | *ABCG2* | 4q22.1 | 0.926 | 0.783 | 3.37 ( 2.23 - 5.11 ) | 9.80E-09 |
| rs2725249 | 4 | 89065868 | C/A | *ABCG2* | 4q22.1 | 0.798 | 0.664 | 1.81 ( 1.35 - 2.42 ) | 7.94E-05 |
| rs2725248 | 4 | 89068007 | A/C | *ABCG2* | 4q22.1 | 0.926 | 0.794 | 2.97 ( 1.99 - 4.44 ) | 1.17E-07 |
| rs2622625 | 4 | 89068737 | C/T | *ABCG2* | 4q22.1 | 0.924 | 0.8 | 3.01 ( 1.99 - 4.55 ) | 1.58E-07 |
| rs6821607 | 4 | 89074808 | G/A | *ABCG2* | 4q22.1 | 0.922 | 0.789 | 3.12 ( 2.07 - 4.7 ) | 5.17E-08 |
| rs3109822 | 4 | 89075223 | C/T | *ABCG2* | 4q22.1 | 0.92 | 0.787 | 2.98 ( 1.99 - 4.46 ) | 1.04E-07 |
| rs145778965 | 4 | 89075239 | C/T | *ABCG2* | 4q22.1 | 0.422 | 0.202 | 3 ( 2.21 - 4.08 ) | 1.98E-12 |
| rs2725239 | 4 | 89075623 | A/C | *ABCG2* | 4q22.1 | 0.793 | 0.648 | 1.82 ( 1.36 - 2.43 ) | 5.66E-05 |
| rs2622604 | 4 | 89078924 | C/T | *ABCG2* | 4q22.1 | 0.922 | 0.792 | 3 ( 2 - 4.52 ) | 1.34E-07 |
| rs3114019 | 4 | 89081441 | T/C | *ABCG2* | 4q22.1 | 0.922 | 0.788 | 3.05 ( 2.03 - 4.59 ) | 8.96E-08 |
| rs3114020 | 4 | 89083666 | C/T | *ABCG2* | 4q22.1 | 0.793 | 0.649 | 1.8 ( 1.34 - 2.4 ) | 7.35E-05 |
| rs2622606 | 4 | 89084381 | T/A | *ABCG2* | 4q22.1 | 0.922 | 0.789 | 3.05 ( 2.03 - 4.59 ) | 8.96E-08 |
| rs2725226 | 4 | 89085331 | A/G | *ABCG2* | 4q22.1 | 0.791 | 0.65 | 1.78 ( 1.33 - 2.38 ) | 9.25E-05 |
| rs2622609 | 4 | 89088475 | C/A | *ABCG2* | 4q22.1 | 0.789 | 0.646 | 1.77 ( 1.33 - 2.35 ) | 7.54E-05 |
| rs28395359 | 4 | 89090448 | A/G | *ABCG2* | 4q22.1 | 0.894 | 0.779 | 2.36 ( 1.63 - 3.41 ) | 5.68E-06 |
| rs76368528 | 4 | 89092524 | G/T | *ABCG2* | 4q22.1 | 0.9 | 0.783 | 2.37 ( 1.62 - 3.45 ) | 7.13E-06 |
| rs66530127 | 4 | 89093013 | G/C | *ABCG2* | 4q22.1 | 0.898 | 0.781 | 2.37 ( 1.63 - 3.45 ) | 6.09E-06 |
| rs66704028 | 4 | 89093036 | C/T | *ABCG2* | 4q22.1 | 0.898 | 0.78 | 2.37 ( 1.63 - 3.45 ) | 6.09E-06 |
| rs55930652 | 4 | 89095690 | G/A | *ABCG2* | 4q22.1 | 0.896 | 0.781 | 2.34 ( 1.61 - 3.4 ) | 8.09E-06 |
| rs183997620 | 4 | 89100289 | C/G | *ABCG2* | 4q22.1 | 0.328 | 0.156 | 2.49 ( 1.82 - 3.41 ) | 1.05E-08 |
| rs72659675 | 4 | 89101603 | G/A | *ABCG2* | 4q22.1 | 0.889 | 0.752 | 2.36 ( 1.67 - 3.33 ) | 1.25E-06 |
| rs10023393 | 4 | 89106527 | C/A | *ABCG2* | 4q22.1 | 0.882 | 0.727 | 2.45 ( 1.74 - 3.43 ) | 2.12E-07 |
| rs72659682 | 4 | 89107539 | A/G | *ABCG2* | 4q22.1 | 0.885 | 0.734 | 2.46 ( 1.75 - 3.45 ) | 2.17E-07 |

| | | | | | | | | | |
|---|---|---|---|---|---|---|---|---|---|
| rs12649505 | 4 | 89108106 | A/G | *ABCG2* | 4q22.1 | 0.891 | 0.751 | 2.22 ( 1.58 - 3.13 ) | 4.74E-06 |
| rs12645232 | 4 | 89110122 | G/A | *ABCG2* | 4q22.1 | 0.885 | 0.735 | 2.41 ( 1.72 - 3.39 ) | 3.76E-07 |
| rs55976258 | 4 | 89117850 | C/T | *ABCG2* | 4q22.1 | 0.872 | 0.724 | 2.33 ( 1.66 - 3.25 ) | 7.64E-07 |
| rs6532055 | 4 | 89118387 | C/T | *ABCG2* | 4q22.1 | 0.868 | 0.723 | 2.31 ( 1.66 - 3.21 ) | 5.64E-07 |
| rs114086764 | 4 | 89118728 | G/A | *ABCG2* | 4q22.1 | 0.878 | 0.723 | 2.62 ( 1.86 - 3.68 ) | 3.08E-08 |
| rs10022955 | 4 | 89119011 | A/G | *ABCG2* | 4q22.1 | 0.763 | 0.634 | 1.78 ( 1.35 - 2.36 ) | 4.91E-05 |
| rs143861620 | 4 | 89121226 | C/T | *ABCG2* | 4q22.1 | 0.837 | 0.654 | 2.43 ( 1.78 - 3.31 ) | 1.96E-08 |
| rs115232777 | 4 | 89121335 | A/C | *ABCG2* | 4q22.1 | 0.837 | 0.656 | 2.37 ( 1.74 - 3.22 ) | 4.42E-08 |
| rs116549154 | 4 | 89121743 | C/T | *ABCG2* | 4q22.1 | 0.837 | 0.655 | 2.38 ( 1.75 - 3.25 ) | 4.06E-08 |
| rs4546214 | 4 | 89122311 | G/C | *ABCG2* | 4q22.1 | 0.857 | 0.665 | 2.32 ( 1.69 - 3.17 ) | 1.54E-07 |
| rs4693930 | 4 | 89122833 | A/G | *ABCG2* | 4q22.1 | 0.835 | 0.651 | 2.44 ( 1.79 - 3.33 ) | 1.90E-08 |
| rs13108819 | 4 | 89123925 | T/C | *ABCG2* | 4q22.1 | 0.835 | 0.651 | 2.44 ( 1.79 - 3.33 ) | 1.90E-08 |
| rs12504163 | 4 | 89124373 | C/T | *ABCG2* | 4q22.1 | 0.728 | 0.579 | 1.72 ( 1.32 - 2.25 ) | 7.30E-05 |
| rs12500185 | 4 | 89124563 | T/C | *ABCG2* | 4q22.1 | 0.835 | 0.651 | 2.44 ( 1.79 - 3.33 ) | 1.90E-08 |
| rs11097184 | 4 | 89125311 | C/T | *ABCG2* | 4q22.1 | 0.835 | 0.65 | 2.44 ( 1.79 - 3.33 ) | 1.90E-08 |
| rs144292084 | 4 | 89125424 | T/A | *ABCG2* | 4q22.1 | 0.837 | 0.654 | 2.41 ( 1.77 - 3.29 ) | 2.90E-08 |
| rs74943514 | 4 | 89125932 | G/C | *ABCG2* | 4q22.1 | 0.837 | 0.655 | 2.38 ( 1.74 - 3.24 ) | 4.36E-08 |
| rs12511059 | 4 | 89126193 | T/C | *ABCG2* | 4q22.1 | 0.833 | 0.652 | 2.36 ( 1.74 - 3.22 ) | 4.54E-08 |
| rs77180571 | 4 | 89126434 | C/T | *ABCG2* | 4q22.1 | 0.837 | 0.655 | 2.38 ( 1.74 - 3.24 ) | 4.36E-08 |
| rs7672396 | 4 | 89126767 | T/C | *ABCG2* | 4q22.1 | 0.835 | 0.651 | 2.38 ( 1.75 - 3.24 ) | 3.57E-08 |
| rs66508773 | 4 | 89127082 | T/C | *ABCG2* | 4q22.1 | 0.835 | 0.651 | 2.41 ( 1.77 - 3.28 ) | 2.87E-08 |
| rs118013835 | 4 | 89128183 | A/G | *ABCG2* | 4q22.1 | 0.174 | 0.07 | 2.74 ( 1.8 - 4.17 ) | 2.36E-06 |
| rs11935352 | 4 | 89128413 | C/T | *ABCG2* | 4q22.1 | 0.835 | 0.652 | 2.41 ( 1.77 - 3.28 ) | 2.87E-08 |
| rs12646307 | 4 | 89129256 | A/T | *ABCG2* | 4q22.1 | 0.837 | 0.655 | 2.38 ( 1.74 - 3.24 ) | 4.36E-08 |
| rs10023457 | 4 | 89134704 | A/T | *ABCG2* | 4q22.1 | 0.726 | 0.573 | 1.75 ( 1.34 - 2.29 ) | 4.35E-05 |
| rs147510135 | 4 | 89135089 | T/C | *ABCG2* | 4q22.1 | 0.226 | 0.109 | 2.23 ( 1.57 - 3.15 ) | 6.33E-06 |

| | | | | | | | | | |
|---|---|---|---|---|---|---|---|---|---|
| rs13127017 | 4 | 89135288 | T/C | *ABCG2* | 4q22.1 | 0.839 | 0.646 | 2.51 ( 1.84 - 3.41 ) | 4.84E-09 |
| rs6819328 | 4 | 89137318 | T/C | *ABCG2* | 4q22.1 | 0.726 | 0.573 | 1.73 ( 1.33 - 2.26 ) | 5.75E-05 |
| rs1986013 | 4 | 89137901 | G/T | *ABCG2* | 4q22.1 | 0.839 | 0.65 | 2.44 ( 1.79 - 3.32 ) | 1.60E-08 |
| rs10019314 | 4 | 89138621 | G/T | *ABCG2* | 4q22.1 | 0.728 | 0.573 | 1.75 ( 1.34 - 2.3 ) | 4.23E-05 |
| rs10007603 | 4 | 89138878 | G/C | *ABCG2* | 4q22.1 | 0.726 | 0.573 | 1.74 ( 1.33 - 2.28 ) | 5.52E-05 |
| rs4693935 | 4 | 89139275 | G/A | *ABCG2* | 4q22.1 | 0.839 | 0.649 | 2.44 ( 1.79 - 3.32 ) | 1.60E-08 |
| rs4693207 | 4 | 89139659 | T/A | *ABCG2* | 4q22.1 | 0.841 | 0.651 | 2.45 ( 1.8 - 3.35 ) | 1.48E-08 |
| rs4264794 | 4 | 89139826 | A/T | *ABCG2* | 4q22.1 | 0.728 | 0.572 | 1.75 ( 1.34 - 2.29 ) | 4.23E-05 |
| rs2904185 | 4 | 89139832 | T/C | *ABCG2* | 4q22.1 | 0.728 | 0.571 | 1.75 ( 1.34 - 2.29 ) | 4.23E-05 |
| rs6840723 | 4 | 89140833 | A/C | *ABCG2* | 4q22.1 | 0.72 | 0.55 | 1.72 ( 1.34 - 2.22 ) | 2.54E-05 |
| rs1383587 | 4 | 89141574 | T/C | *ABCG2* | 4q22.1 | 0.791 | 0.581 | 2.48 ( 1.86 - 3.31 ) | 6.59E-10 |
| rs13119417 | 4 | 89143112 | G/A | *ABCG2* | 4q22.1 | 0.793 | 0.583 | 2.51 ( 1.88 - 3.34 ) | 4.67E-10 |
| rs1904903 | 4 | 89145274 | T/A | *ABCG2* | 4q22.1 | 0.791 | 0.583 | 2.48 ( 1.86 - 3.32 ) | 6.47E-10 |
| rs1074839 | 4 | 89145548 | T/C | *ABCG2* | 4q22.1 | 0.789 | 0.583 | 2.44 ( 1.83 - 3.25 ) | 1.15E-09 |
| rs2046132 | 4 | 89149026 | G/C | *ABCG2* | 4q22.1 | 0.789 | 0.584 | 2.44 ( 1.84 - 3.26 ) | 1.03E-09 |
| rs13107048 | 4 | 89150075 | C/T | *ABCG2* | 4q22.1 | 0.791 | 0.581 | 2.49 ( 1.87 - 3.32 ) | 5.09E-10 |
| rs111955262 | 4 | 89150226 | T/C | *ABCG2* | 4q22.1 | 0.832 | 0.645 | 2.38 ( 1.76 - 3.23 ) | 1.87E-08 |
| rs72554040 | 4 | 89152324 | G/A | *ABCG2* | 4q22.1 | 0.804 | 0.604 | 2.47 ( 1.84 - 3.3 ) | 1.29E-09 |
| rs61046194 | 4 | 89154025 | A/T | *ABCG2 - PPM1K* | 4q22.1 | 0.943 | 0.854 | 2.95 ( 1.85 - 4.69 ) | 5.04E-06 |
| rs117104615 | 4 | 89154065 | G/T | *ABCG2 - PPM1K* | 4q22.1 | 0.839 | 0.664 | 2.31 ( 1.7 - 3.13 ) | 8.91E-08 |
| rs6846256 | 4 | 89154111 | G/T | *ABCG2 - PPM1K* | 4q22.1 | 0.809 | 0.616 | 2.46 ( 1.83 - 3.31 ) | 2.95E-09 |
| rs12641384 | 4 | 89155237 | C/G | *ABCG2 - PPM1K* | 4q22.1 | 0.837 | 0.65 | 2.47 ( 1.82 - 3.37 ) | 7.69E-09 |
| rs13328043 | 4 | 89155971 | A/C | *ABCG2 - PPM1K* | 4q22.1 | 0.788 | 0.584 | 2.36 ( 1.78 - 3.13 ) | 2.01E-09 |
| rs28883356 | 4 | 89156061 | C/T | *ABCG2 - PPM1K* | 4q22.1 | 0.794 | 0.588 | 2.5 ( 1.87 - 3.35 ) | 6.50E-10 |
| rs4693941 | 4 | 89156899 | T/A | *ABCG2 - PPM1K* | 4q22.1 | 0.793 | 0.588 | 2.46 ( 1.84 - 3.28 ) | 1.18E-09 |
| rs60190393 | 4 | 89156914 | T/A | *ABCG2 - PPM1K* | 4q22.1 | 0.837 | 0.65 | 2.47 ( 1.82 - 3.36 ) | 8.22E-09 |

| | | | | | | | | | |
|---|---|---|---|---|---|---|---|---|---|
| rs7437679 | 4 | 89157124 | A/C | *ABCG2 - PPM1K* | 4q22.1 | 0.811 | 0.61 | 2.54 ( 1.89 - 3.42 ) | 6.92E-10 |
| rs13120254 | 4 | 89160561 | A/C | *ABCG2 - PPM1K* | 4q22.1 | 0.793 | 0.588 | 2.48 ( 1.86 - 3.31 ) | 8.14E-10 |
| rs13120819 | 4 | 89160677 | A/G | *ABCG2 - PPM1K* | 4q22.1 | 0.793 | 0.589 | 2.43 ( 1.83 - 3.25 ) | 1.40E-09 |
| rs79659488 | 4 | 89161140 | G/T | *ABCG2 - PPM1K* | 4q22.1 | 0.837 | 0.649 | 2.45 ( 1.8 - 3.32 ) | 9.65E-09 |
| rs77787810 | 4 | 89161195 | A/G | *ABCG2 - PPM1K* | 4q22.1 | 0.837 | 0.649 | 2.45 ( 1.8 - 3.32 ) | 9.65E-09 |
| rs78685931 | 4 | 89161399 | A/T | *ABCG2 - PPM1K* | 4q22.1 | 0.839 | 0.65 | 2.48 ( 1.82 - 3.37 ) | 6.92E-09 |
| rs1481018 | 4 | 89162388 | A/G | *ABCG2 - PPM1K* | 4q22.1 | 0.794 | 0.589 | 2.48 ( 1.85 - 3.31 ) | 8.72E-10 |
| rs7664939 | 4 | 89162700 | A/G | *ABCG2 - PPM1K* | 4q22.1 | 0.837 | 0.651 | 2.45 ( 1.8 - 3.32 ) | 9.65E-09 |
| rs80179435 | 4 | 89163127 | C/T | *ABCG2 - PPM1K* | 4q22.1 | 0.837 | 0.649 | 2.45 ( 1.8 - 3.32 ) | 9.65E-09 |
| rs997630 | 4 | 89163853 | G/A | *ABCG2 - PPM1K* | 4q22.1 | 0.794 | 0.589 | 2.45 ( 1.83 - 3.27 ) | 1.30E-09 |
| rs922674 | 4 | 89164722 | G/A | *ABCG2 - PPM1K* | 4q22.1 | 0.794 | 0.588 | 2.48 ( 1.85 - 3.31 ) | 8.72E-10 |
| rs6532060 | 4 | 89165494 | T/C | *ABCG2 - PPM1K* | 4q22.1 | 0.783 | 0.583 | 2.42 ( 1.81 - 3.23 ) | 2.09E-09 |
| rs6532061 | 4 | 89165613 | T/C | *ABCG2 - PPM1K* | 4q22.1 | 0.782 | 0.585 | 2.36 ( 1.77 - 3.14 ) | 5.24E-09 |
| rs7656113 | 4 | 89166536 | C/A | *ABCG2 - PPM1K* | 4q22.1 | 0.832 | 0.664 | 2.14 ( 1.57 - 2.9 ) | 1.13E-06 |
| rs17013965 | 4 | 89170730 | G/A | *ABCG2 - PPM1K* | 4q22.1 | 0.809 | 0.669 | 1.88 ( 1.4 - 2.53 ) | 2.48E-05 |
| rs4693943 | 4 | 89171935 | T/G | *ABCG2 - PPM1K* | 4q22.1 | 0.809 | 0.667 | 1.88 ( 1.4 - 2.53 ) | 2.48E-05 |
| rs2045797 | 4 | 103353074 | C/G | *LOC105377621* | 4q24 | 0.433 | 0.34 | 1.77 ( 1.37 - 2.3 ) | 1.70E-05 |
| rs6533015 | 4 | 103355533 | G/C | *LOC105377621* | 4q24 | 0.428 | 0.335 | 1.77 ( 1.37 - 2.3 ) | 1.47E-05 |
| rs62328509 | 4 | 103357740 | T/G | *LOC105377621* | 4q24 | 0.411 | 0.304 | 1.87 ( 1.44 - 2.44 ) | 3.22E-06 |
| rs1811810 | 4 | 103360830 | A/G | *LOC105377621* | 4q24 | 0.428 | 0.335 | 1.77 ( 1.37 - 2.3 ) | 1.53E-05 |
| rs1609992 | 4 | 103361105 | G/A | *LOC105377621* | 4q24 | 0.406 | 0.299 | 1.89 ( 1.44 - 2.46 ) | 3.44E-06 |
| rs12710964 | 4 | 103361790 | A/G | *LOC105377621* | 4q24 | 0.43 | 0.337 | 1.76 ( 1.36 - 2.28 ) | 2.02E-05 |
| rs1037999 | 4 | 103362252 | T/C | *LOC105377621* | 4q24 | 0.43 | 0.337 | 1.76 ( 1.36 - 2.28 ) | 2.02E-05 |
| rs66521590 | 4 | 103363446 | A/G | *LOC105377621* | 4q24 | 0.404 | 0.298 | 1.86 ( 1.43 - 2.44 ) | 4.90E-06 |
| rs6851440 | 4 | 103363638 | C/T | *LOC105377621* | 4q24 | 0.406 | 0.298 | 1.9 ( 1.45 - 2.49 ) | 2.60E-06 |
| rs2169598 | 4 | 103365842 | T/C | *LOC105377621* | 4q24 | 0.426 | 0.337 | 1.74 ( 1.34 - 2.25 ) | 3.10E-05 |

| | | | | | | | | | |
|---|---|---|---|---|---|---|---|---|---|
| rs2169597 | 4 | 103368598 | T/C | *LOC105377621* | 4q24 | 0.428 | 0.337 | 1.75 ( 1.35 - 2.27 ) | 2.44E-05 |
| rs1973871 | 4 | 103371426 | T/C | *LOC105377621* | 4q24 | 0.428 | 0.336 | 1.75 ( 1.35 - 2.27 ) | 2.44E-05 |
| rs75810487 | 4 | 103372494 | */A | . | . | 0.426 | 0.337 | 1.75 ( 1.35 - 2.28 ) | 2.73E-05 |
| rs1313924 | 4 | 103375303 | A/C | *LOC105377621* | 4q24 | 0.459 | 0.368 | 1.69 ( 1.31 - 2.18 ) | 4.62E-05 |
| rs1313925 | 4 | 103377576 | C/T | *LOC105377621* | 4q24 | 0.468 | 0.371 | 1.73 ( 1.34 - 2.23 ) | 2.17E-05 |
| rs1314336 | 4 | 103378866 | T/C | *LOC105377621* | 4q24 | 0.467 | 0.368 | 1.73 ( 1.34 - 2.23 ) | 2.22E-05 |
| rs230490 | 4 | 103387419 | G/A | *LOC105377621* | 4q24 | 0.465 | 0.371 | 1.72 ( 1.33 - 2.22 ) | 3.01E-05 |
| rs4467509 | 4 | 156196646 | A/G | *NPY2R - MAP9* | 4q32.1 | 0.232 | 0.187 | 1.96 ( 1.4 - 2.74 ) | 9.02E-05 |
| rs1947068 | 4 | 156197412 | C/T | *NPY2R - MAP9* | 4q32.1 | 0.232 | 0.187 | 1.96 ( 1.4 - 2.74 ) | 9.02E-05 |
| rs141876887 | 4 | 181972398 | C/T | *NONE - LINC00290* | 4q34.3 | 0.118 | 0.075 | 2.67 ( 1.63 - 4.38 ) | 9.94E-05 |
| rs28456058 | 4 | 187414343 | T/C | *F11-AS1* | 4q35.2 | 0.304 | 0.233 | 1.9 ( 1.4 - 2.57 ) | 3.17E-05 |
| rs28755472 | 4 | 187414790 | A/G | *F11-AS1* | 4q35.2 | 0.298 | 0.229 | 1.85 ( 1.37 - 2.5 ) | 6.46E-05 |
| rs349583 | 5 | 436665 | G/A | *AHRR* | 5p15.33 | 0.539 | 0.466 | 1.67 ( 1.29 - 2.15 ) | 7.68E-05 |
| rs230258 | 6 | 8553118 | T/C | *LOC100506207* | 6p24.3 | 0.322 | 0.274 | 1.79 ( 1.35 - 2.39 ) | 6.53E-05 |
| rs850195 | 6 | 8558497 | C/T | *LOC100506207* | 6p24.3 | 0.32 | 0.276 | 1.79 ( 1.34 - 2.39 ) | 8.15E-05 |
| rs850193 | 6 | 8559060 | T/G | *LOC100506207* | 6p24.3 | 0.32 | 0.276 | 1.79 ( 1.34 - 2.39 ) | 8.15E-05 |
| rs850192 | 6 | 8560888 | T/G | *LOC100506207* | 6p24.3 | 0.32 | 0.273 | 1.81 ( 1.36 - 2.42 ) | 5.93E-05 |
| rs850188 | 6 | 8561815 | C/A | *LOC100506207* | 6p24.3 | 0.32 | 0.273 | 1.81 ( 1.36 - 2.42 ) | 5.93E-05 |
| rs6932780 | 6 | 33835652 | T/C | *MLN - LINC01016* | 6p21.31 | 0.883 | 0.838 | 2.05 ( 1.43 - 2.94 ) | 9.90E-05 |
| rs234480 | 6 | 124355296 | T/C | *NKAIN2* | 6q22.31 | 0.682 | 0.576 | 1.73 ( 1.33 - 2.26 ) | 4.05E-05 |
| rs117464851 | 6 | 152109935 | C/T | *ESR1* | 6q25.1 | 0.793 | 0.699 | 1.83 ( 1.38 - 2.44 ) | 3.15E-05 |
| rs9371227 | 6 | 152133617 | C/A | *ESR1* | 6q25.1 | 0.794 | 0.701 | 1.81 ( 1.36 - 2.4 ) | 4.37E-05 |
| rs9371557 | 6 | 152140209 | A/G | *ESR1* | 6q25.1 | 0.793 | 0.702 | 1.76 ( 1.33 - 2.34 ) | 9.25E-05 |
| rs9456488 | 6 | 160396737 | A/G | *IGF2R* | 6q25.3 | 0.45 | 0.385 | 1.65 ( 1.3 - 2.09 ) | 3.45E-05 |
| rs77869950 | 7 | 16996425 | G/A | *AGR3 - AHR* | 7p21.1 | 0.924 | 0.881 | 2.41 ( 1.57 - 3.7 ) | 5.32E-05 |
| rs2731568 | 7 | 18113008 | C/T | *PRPS1L1 - HDAC9* | 7p21.1 | 0.424 | 0.324 | 1.75 ( 1.34 - 2.3 ) | 4.51E-05 |

| | | | | | | | | | |
|---|---|---|---|---|---|---|---|---|---|
| rs58705226 | 7 | 18120806 | A/C | *PRPS1L1 - HDAC9* | 7p21.1 | 0.498 | 0.404 | 1.74 ( 1.34 - 2.24 ) | 2.60E-05 |
| rs12700326 | 7 | 21955738 | G/A | *CDCA7L* | 7p15.3 | 0.911 | 0.865 | 2.27 ( 1.52 - 3.4 ) | 6.96E-05 |
| rs75682841 | 7 | 28463166 | C/T | *CREB5* | 7p15.1 | 0.106 | 0.064 | 2.82 ( 1.7 - 4.69 ) | 6.19E-05 |
| rs2160000 | 7 | 36513617 | A/C | *ANLN - AOAH* | 7p14.2 | 0.12 | 0.069 | 2.82 ( 1.71 - 4.65 ) | 4.52E-05 |
| rs6943424 | 7 | 77641545 | T/G | *PHTF2 - MAGI2* | 7q21.11 | 0.672 | 0.589 | 1.65 ( 1.28 - 2.11 ) | 8.39E-05 |
| rs56963120 | 7 | 88351391 | T/A | *STEAP4 - ZNF804B* | 7q21.13 | 0.382 | 0.297 | 1.73 ( 1.31 - 2.28 ) | 9.74E-05 |
| rs17163911 | 7 | 88352832 | C/T | *STEAP4 - ZNF804B* | 7q21.13 | 0.387 | 0.298 | 1.74 ( 1.32 - 2.29 ) | 9.20E-05 |
| rs9641035 | 7 | 88421333 | T/C | *ZNF804B* | 7q21.13 | 0.378 | 0.288 | 1.83 ( 1.38 - 2.42 ) | 2.29E-05 |
| rs888604 | 7 | 131295022 | C/A | *PODXL - LOC101928782* | 7q32.3 | 0.285 | 0.212 | 1.87 ( 1.38 - 2.54 ) | 6.20E-05 |
| rs2971750 | 7 | 131301170 | A/G | *PODXL - LOC101928782* | 7q32.3 | 0.317 | 0.243 | 1.8 ( 1.36 - 2.4 ) | 5.24E-05 |
| rs75119659 | 7 | 131301405 | A/G | *PODXL - LOC101928782* | 7q32.3 | 0.311 | 0.236 | 1.77 ( 1.33 - 2.35 ) | 9.19E-05 |
| rs67337609 | 7 | 131339495 | T/C | *PODXL - LOC101928782* | 7q32.3 | 0.957 | 0.844 | 3.14 ( 1.9 - 5.19 ) | 8.50E-06 |
| rs73159568 | 7 | 131339498 | G/T | *PODXL - LOC101928782* | 7q32.3 | 0.957 | 0.848 | 3.14 ( 1.89 - 5.22 ) | 9.64E-06 |
| rs73158732 | 7 | 136606172 | A/C | *LOC349160* | 7q33 | 0.709 | 0.609 | 1.81 ( 1.37 - 2.38 ) | 3.04E-05 |
| rs1364407 | 7 | 136609925 | C/T | *LOC349160* | 7q33 | 0.676 | 0.569 | 1.77 ( 1.34 - 2.33 ) | 4.83E-05 |
| rs1364409 | 7 | 136612033 | T/A | *LOC349160* | 7q33 | 0.674 | 0.568 | 1.75 ( 1.33 - 2.31 ) | 6.78E-05 |
| rs978437 | 7 | 136614178 | C/T | *LOC349160* | 7q33 | 0.676 | 0.568 | 1.78 ( 1.35 - 2.35 ) | 3.92E-05 |
| rs7792449 | 7 | 136617722 | G/A | *LOC349160* | 7q33 | 0.678 | 0.57 | 1.78 ( 1.35 - 2.35 ) | 4.17E-05 |
| rs7795766 | 7 | 136617796 | C/* | . | . | 0.707 | 0.603 | 1.77 ( 1.34 - 2.34 ) | 6.32E-05 |
| rs10267979 | 7 | 136618188 | T/A | *LOC349160* | 7q33 | 0.706 | 0.603 | 1.76 ( 1.33 - 2.33 ) | 7.41E-05 |
| rs35246768 | 7 | 136619959 | T/C | *LOC349160* | 7q33 | 0.676 | 0.57 | 1.77 ( 1.35 - 2.34 ) | 4.84E-05 |
| rs7806357 | 7 | 136620152 | C/T | *LOC349160* | 7q33 | 0.706 | 0.604 | 1.75 ( 1.32 - 2.31 ) | 8.83E-05 |
| rs7782965 | 7 | 136624133 | T/C | *LOC349160* | 7q33 | 0.676 | 0.569 | 1.79 ( 1.36 - 2.36 ) | 3.46E-05 |
| rs58394792 | 7 | 136626027 | G/A | *LOC349160* | 7q33 | 0.707 | 0.607 | 1.74 ( 1.32 - 2.3 ) | 9.01E-05 |
| rs17168819 | 7 | 136628299 | T/C | *LOC349160* | 7q33 | 0.706 | 0.604 | 1.74 ( 1.32 - 2.3 ) | 8.59E-05 |
| rs201752458 | 7 | 136629104 | G/* | . | . | 0.702 | 0.598 | 1.75 ( 1.32 - 2.31 ) | 9.13E-05 |

| | | | | | | | | | |
|---|---|---|---|---|---|---|---|---|---|
| rs12374823 | 7 | 150563978 | A/C | *AOC1 - KCNH2* | 7q36.1 | 0.73 | 0.653 | 1.74 ( 1.32 - 2.3 ) | 7.46E-05 |
| rs10236839 | 7 | 150564287 | C/T | *AOC1 - KCNH2* | 7q36.1 | 0.689 | 0.604 | 1.72 ( 1.31 - 2.24 ) | 8.06E-05 |
| rs10254616 | 7 | 150565220 | G/A | *AOC1 - KCNH2* | 7q36.1 | 0.689 | 0.604 | 1.72 ( 1.31 - 2.24 ) | 8.06E-05 |
| rs78506532 | 8 | 5476076 | T/G | *CSMD1 - LOC100287015* | 8p23.2 | 0.831 | 0.719 | 1.82 ( 1.36 - 2.44 ) | 5.47E-05 |
| rs1477926733 | 8 | 51993276 | */T | . | . | 0.891 | 0.729 | 2.75 ( 1.85 - 4.09 ) | 5.73E-07 |
| rs60355343 | 9 | 4434012 | C/A | *GLIS3 - SLC1A1* | 9p24.2 | 0.459 | 0.393 | 1.69 ( 1.31 - 2.18 ) | 5.90E-05 |
| rs57649467 | 9 | 101632854 | A/G | *GALNT12 - COL15A1* | 9q22.33 | 0.541 | 0.447 | 1.7 ( 1.31 - 2.19 ) | 5.95E-05 |
| rs13283283 | 9 | 101633504 | G/T | *GALNT12 - COL15A1* | 9q22.33 | 0.593 | 0.492 | 1.66 ( 1.29 - 2.14 ) | 8.24E-05 |
| rs13289251 | 9 | 101633530 | T/C | *GALNT12 - COL15A1* | 9q22.33 | 0.593 | 0.492 | 1.66 ( 1.29 - 2.14 ) | 8.24E-05 |
| rs2295930 | 9 | 101634511 | T/C | *GALNT12 - COL15A1* | 9q22.33 | 0.541 | 0.447 | 1.71 ( 1.33 - 2.21 ) | 3.68E-05 |
| rs2295931 | 9 | 101634619 | T/G | *GALNT12 - COL15A1* | 9q22.33 | 0.543 | 0.447 | 1.75 ( 1.35 - 2.26 ) | 2.03E-05 |
| rs62562032 | 9 | 101636230 | T/C | *GALNT12 - COL15A1* | 9q22.33 | 0.543 | 0.448 | 1.73 ( 1.34 - 2.24 ) | 2.71E-05 |
| rs7466147 | 9 | 101636786 | A/G | *GALNT12 - COL15A1* | 9q22.33 | 0.541 | 0.445 | 1.74 ( 1.35 - 2.25 ) | 2.20E-05 |
| rs4135168 | 9 | 113016885 | T/C | *TXN* | 9q31.3 | 0.97 | 0.925 | 3.65 ( 1.95 - 6.83 ) | 4.92E-05 |
| rs10124196 | 9 | 132417991 | C/T | *ASB6 - PRRX2* | 9q34.11 | 0.783 | 0.72 | 1.79 ( 1.35 - 2.38 ) | 4.83E-05 |
| rs603261 | 10 | 14393233 | T/C | *FRMD4A - MIR4293* | 10p13 | 0.67 | 0.566 | 1.76 ( 1.36 - 2.28 ) | 1.88E-05 |
| rs601853 | 10 | 14393558 | C/T | *FRMD4A - MIR4293* | 10p13 | 0.691 | 0.582 | 1.77 ( 1.36 - 2.3 ) | 2.11E-05 |
| rs11259026 | 10 | 14395591 | G/C | *FRMD4A - MIR4293* | 10p13 | 0.606 | 0.515 | 1.65 ( 1.28 - 2.13 ) | 9.90E-05 |
| rs482468 | 10 | 27671644 | G/C | *ARMC4P1 - PTCHD3* | 10p12.1 | 0.444 | 0.369 | 1.67 ( 1.29 - 2.15 ) | 8.33E-05 |
| rs10829253 | 10 | 27675276 | T/C | *ARMC4P1 - PTCHD3* | 10p12.1 | 0.42 | 0.35 | 1.67 ( 1.29 - 2.15 ) | 8.26E-05 |
| rs61591337 | 10 | 27678461 | T/C | *ARMC4P1 - PTCHD3* | 10p12.1 | 0.418 | 0.35 | 1.67 ( 1.3 - 2.16 ) | 7.76E-05 |
| rs7100677 | 10 | 27681526 | A/G | *ARMC4P1 - PTCHD3* | 10p12.1 | 0.428 | 0.359 | 1.66 ( 1.29 - 2.15 ) | 8.75E-05 |
| rs11015740 | 10 | 27690072 | G/T | *PTCHD3* | 10p12.1 | 0.428 | 0.351 | 1.7 ( 1.32 - 2.19 ) | 4.52E-05 |
| rs2768693 | 10 | 29578351 | C/T | *LYZL1* | 10p12.1 | 0.645 | 0.539 | 1.72 ( 1.32 - 2.24 ) | 6.20E-05 |
| rs111812553 | 10 | 76624101 | C/T | *KAT6B* | 10q22.2 | 0.935 | 0.886 | 2.47 ( 1.57 - 3.88 ) | 8.61E-05 |
| rs10509352 | 10 | 76637516 | A/G | *KAT6B* | 10q22.2 | 0.935 | 0.885 | 2.55 ( 1.63 - 4 ) | 4.28E-05 |

| | | | | | | | | | |
|---|---|---|---|---|---|---|---|---|---|
| rs58330860 | 10 | 76638350 | T/G | *KAT6B* | 10q22.2 | 0.933 | 0.886 | 2.48 ( 1.59 - 3.86 ) | 6.53E-05 |
| rs11813058 | 10 | 76643027 | C/A | *KAT6B* | 10q22.2 | 0.933 | 0.884 | 2.48 ( 1.59 - 3.86 ) | 6.53E-05 |
| rs11813774 | 10 | 76644547 | G/T | *KAT6B* | 10q22.2 | 0.933 | 0.884 | 2.48 ( 1.59 - 3.86 ) | 6.53E-05 |
| rs58255635 | 10 | 76652783 | C/T | *KAT6B* | 10q22.2 | 0.933 | 0.884 | 2.48 ( 1.59 - 3.86 ) | 6.53E-05 |
| rs4746246 | 10 | 76658548 | A/G | *KAT6B* | 10q22.2 | 0.935 | 0.884 | 2.5 ( 1.6 - 3.91 ) | 5.57E-05 |
| rs75595613 | 10 | 76679000 | A/T | *LOC101929165* | 10q22.2 | 0.935 | 0.881 | 2.67 ( 1.71 - 4.17 ) | 1.60E-05 |
| rs150163015 | 10 | 76698564 | G/A | *KAT6B* | 10q22.2 | 0.935 | 0.885 | 2.5 ( 1.6 - 3.91 ) | 5.57E-05 |
| rs7905775 | 10 | 76704830 | A/G | *KAT6B* | 10q22.2 | 0.935 | 0.885 | 2.5 ( 1.6 - 3.91 ) | 5.57E-05 |
| rs529246328 | 10 | 111169490 | A/* | . | . | 0.857 | 0.815 | 2.23 ( 1.54 - 3.24 ) | 2.28E-05 |
| rs9630104 | 10 | 117051755 | C/T | *ATRNL1* | 10q25.3 | 0.743 | 0.649 | 1.74 ( 1.34 - 2.26 ) | 3.50E-05 |
| rs615905 | 10 | 117139057 | A/G | *ATRNL1* | 10q25.3 | 0.778 | 0.678 | 1.77 ( 1.33 - 2.35 ) | 9.27E-05 |
| rs2485971 | 10 | 117141509 | A/G | *ATRNL1* | 10q25.3 | 0.776 | 0.676 | 1.79 ( 1.34 - 2.38 ) | 7.18E-05 |
| rs1565833 | 10 | 117142374 | A/G | *ATRNL1* | 10q25.3 | 0.778 | 0.677 | 1.79 ( 1.34 - 2.38 ) | 6.97E-05 |
| rs2485966 | 10 | 117157219 | T/C | *ATRNL1* | 10q25.3 | 0.774 | 0.674 | 1.76 ( 1.32 - 2.34 ) | 1.00E-04 |
| rs594160 | 10 | 117163566 | T/C | *ATRNL1* | 10q25.3 | 0.776 | 0.675 | 1.79 ( 1.34 - 2.38 ) | 7.18E-05 |
| rs621652 | 10 | 117180598 | A/T | *ATRNL1* | 10q25.3 | 0.787 | 0.686 | 1.83 ( 1.36 - 2.45 ) | 5.61E-05 |
| rs877023 | 10 | 132170008 | G/A | *GLRX3 - MIR378C* | 10q26.3 | 0.335 | 0.265 | 1.81 ( 1.36 - 2.41 ) | 3.95E-05 |
| rs378120 | 10 | 132174169 | G/A | *GLRX3 - MIR378C* | 10q26.3 | 0.463 | 0.4 | 1.79 ( 1.36 - 2.35 ) | 3.70E-05 |
| rs394356 | 10 | 132174428 | C/A | *GLRX3 - MIR378C* | 10q26.3 | 0.461 | 0.4 | 1.76 ( 1.34 - 2.32 ) | 6.09E-05 |
| rs2013870 | 10 | 132174491 | A/G | *GLRX3 - MIR378C* | 10q26.3 | 0.461 | 0.398 | 1.78 ( 1.35 - 2.35 ) | 3.90E-05 |
| rs1872315 | 10 | 132174628 | G/A | *GLRX3 - MIR378C* | 10q26.3 | 0.463 | 0.4 | 1.77 ( 1.35 - 2.34 ) | 4.66E-05 |
| rs1872316 | 10 | 132174704 | A/G | *GLRX3 - MIR378C* | 10q26.3 | 0.461 | 0.401 | 1.73 ( 1.32 - 2.28 ) | 8.67E-05 |
| rs10764951 | 10 | 132174900 | G/T | *GLRX3 - MIR378C* | 10q26.3 | 0.461 | 0.397 | 1.78 ( 1.36 - 2.34 ) | 3.64E-05 |
| rs11017223 | 10 | 132174977 | C/T | *GLRX3 - MIR378C* | 10q26.3 | 0.461 | 0.4 | 1.76 ( 1.34 - 2.32 ) | 6.09E-05 |
| rs11017224 | 10 | 132175036 | G/A | *GLRX3 - MIR378C* | 10q26.3 | 0.459 | 0.401 | 1.74 ( 1.32 - 2.29 ) | 8.03E-05 |
| rs4751198 | 10 | 132175048 | G/A | *GLRX3 - MIR378C* | 10q26.3 | 0.459 | 0.401 | 1.74 ( 1.32 - 2.29 ) | 8.03E-05 |

| | | | | | | | | | |
|---|---|---|---|---|---|---|---|---|---|
| rs612028 | 10 | 132177303 | A/C | *GLRX3 - MIR378C* | 10q26.3 | 0.459 | 0.4 | 1.74 ( 1.32 - 2.29 ) | 7.89E-05 |
| rs594754 | 10 | 132177344 | G/A | *GLRX3 - MIR378C* | 10q26.3 | 0.459 | 0.401 | 1.74 ( 1.32 - 2.29 ) | 7.89E-05 |
| rs624248 | 10 | 132177712 | C/T | *GLRX3 - MIR378C* | 10q26.3 | 0.459 | 0.401 | 1.73 ( 1.31 - 2.27 ) | 9.43E-05 |
| rs624276 | 10 | 132177733 | C/G | *GLRX3 - MIR378C* | 10q26.3 | 0.459 | 0.4 | 1.74 ( 1.32 - 2.29 ) | 7.89E-05 |
| rs529603 | 10 | 132178240 | C/T | *GLRX3 - MIR378C* | 10q26.3 | 0.459 | 0.4 | 1.74 ( 1.32 - 2.29 ) | 7.89E-05 |
| rs626919 | 10 | 132178340 | G/A | *GLRX3 - MIR378C* | 10q26.3 | 0.459 | 0.401 | 1.74 ( 1.32 - 2.29 ) | 7.89E-05 |
| rs367259 | 10 | 132179257 | C/G | *GLRX3 - MIR378C* | 10q26.3 | 0.459 | 0.4 | 1.74 ( 1.32 - 2.29 ) | 7.89E-05 |
| rs7084088 | 10 | 132179475 | T/G | *GLRX3 - MIR378C* | 10q26.3 | 0.459 | 0.398 | 1.77 ( 1.34 - 2.33 ) | 5.08E-05 |
| rs652855 | 10 | 132179539 | G/A | *GLRX3 - MIR378C* | 10q26.3 | 0.459 | 0.4 | 1.74 ( 1.32 - 2.29 ) | 7.89E-05 |
| rs405494 | 10 | 132179651 | A/G | *GLRX3 - MIR378C* | 10q26.3 | 0.457 | 0.401 | 1.73 ( 1.31 - 2.27 ) | 9.92E-05 |
| rs444289 | 10 | 132179689 | C/T | *GLRX3 - MIR378C* | 10q26.3 | 0.461 | 0.401 | 1.75 ( 1.33 - 2.31 ) | 6.45E-05 |
| rs7092943 | 10 | 132180956 | C/G | *GLRX3 - MIR378C* | 10q26.3 | 0.459 | 0.398 | 1.75 ( 1.33 - 2.3 ) | 6.73E-05 |
| rs407020 | 10 | 132187990 | G/A | *GLRX3 - MIR378C* | 10q26.3 | 0.461 | 0.402 | 1.74 ( 1.32 - 2.28 ) | 8.03E-05 |
| rs382014 | 10 | 132191419 | C/T | *GLRX3 - MIR378C* | 10q26.3 | 0.461 | 0.402 | 1.73 ( 1.32 - 2.28 ) | 8.54E-05 |
| rs1785631 | 11 | 66092730 | G/A | *CD248 - RIN1* | 11q13.2 | 0.468 | 0.412 | 1.66 ( 1.3 - 2.13 ) | 6.10E-05 |
| rs1402308385 | 11 | 67467339 | G/C | *ALDH3B2 - FAM86C2P* | 11q13.2 | 0.072 | 0.056 | 3.96 ( 2.01 - 7.82 ) | 7.27E-05 |
| rs4427595 | 11 | 96899508 | T/A | *LOC105369443 - CNTN5* | 11q21 | 0.676 | 0.599 | 1.66 ( 1.29 - 2.15 ) | 9.45E-05 |
| rs4753997 | 11 | 96901174 | C/G | *LOC105369443 - CNTN5* | 11q21 | 0.68 | 0.596 | 1.66 ( 1.3 - 2.12 ) | 5.17E-05 |
| rs147355747 | 12 | 50221860 | G/A | *NCKAP5L* | 12q13.12 | 0.944 | 0.884 | 2.51 ( 1.58 - 3.97 ) | 9.42E-05 |
| rs12813681 | 12 | 69767492 | T/C | *YEATS4* | 12q15 | 0.715 | 0.642 | 1.72 ( 1.31 - 2.25 ) | 8.61E-05 |
| rs117426963 | 12 | 69768344 | G/A | *YEATS4* | 12q15 | 0.717 | 0.641 | 1.74 ( 1.33 - 2.27 ) | 5.83E-05 |
| rs34111034 | 12 | 69779210 | T/G | *YEATS4* | 12q15 | 0.715 | 0.644 | 1.72 ( 1.31 - 2.25 ) | 8.66E-05 |
| rs12816616 | 12 | 69780691 | A/G | *YEATS4* | 12q15 | 0.715 | 0.645 | 1.72 ( 1.31 - 2.25 ) | 8.66E-05 |
| rs12826219 | 12 | 69785576 | A/C | *YEATS4* | 12q15 | 0.715 | 0.643 | 1.71 ( 1.31 - 2.24 ) | 9.25E-05 |
| rs10859907 | 12 | 95995906 | T/C | *USP44 - PGAM1P5* | 12q22 | 0.491 | 0.401 | 1.75 ( 1.35 - 2.27 ) | 2.61E-05 |
| rs11109766 | 12 | 99626644 | G/C | *ANKS1B* | 12q23.1 | 0.602 | 0.507 | 1.79 ( 1.39 - 2.32 ) | 7.52E-06 |

| | | | | | | | | | |
|---|---|---|---|---|---|---|---|---|---|
| rs2220689 | 12 | 99629188 | G/C | *ANKS1B* | 12q23.1 | 0.596 | 0.515 | 1.76 ( 1.36 - 2.28 ) | 1.88E-05 |
| rs73140985 | 12 | 99638989 | G/A | *ANKS1B* | 12q23.1 | 0.617 | 0.528 | 1.76 ( 1.36 - 2.28 ) | 1.69E-05 |
| rs11109773 | 12 | 99639729 | C/T | *ANKS1B* | 12q23.1 | 0.668 | 0.582 | 1.71 ( 1.32 - 2.23 ) | 5.89E-05 |
| rs923724 | 12 | 99639930 | C/G | *ANKS1B* | 12q23.1 | 0.672 | 0.599 | 1.74 ( 1.33 - 2.27 ) | 4.78E-05 |
| rs11109775 | 12 | 99641804 | G/A | *ANKS1B* | 12q23.1 | 0.678 | 0.614 | 1.7 ( 1.3 - 2.21 ) | 9.58E-05 |
| rs2245763 | 12 | 102929425 | G/A | *IGF1 - LINC00485* | 12q23.2 | 0.557 | 0.484 | 1.7 ( 1.3 - 2.22 ) | 9.38E-05 |
| rs2607988 | 12 | 102929883 | G/A | *IGF1 - LINC00485* | 12q23.2 | 0.557 | 0.483 | 1.75 ( 1.33 - 2.28 ) | 4.76E-05 |
| rs703544 | 12 | 102942355 | G/A | *IGF1 - LINC00485* | 12q23.2 | 0.559 | 0.474 | 1.72 ( 1.31 - 2.24 ) | 7.77E-05 |
| rs703545 | 12 | 102943000 | A/G | *IGF1 - LINC00485* | 12q23.2 | 0.556 | 0.473 | 1.71 ( 1.31 - 2.24 ) | 7.92E-05 |
| rs10735400 | 12 | 105689161 | C/T | *APPL2 - KCCAT198* | 12q23.3 | 0.407 | 0.309 | 1.78 ( 1.37 - 2.31 ) | 1.92E-05 |
| rs10746014 | 12 | 105689315 | T/C | *APPL2 - KCCAT198* | 12q23.3 | 0.407 | 0.309 | 1.78 ( 1.37 - 2.31 ) | 1.92E-05 |
| rs10746015 | 12 | 105689407 | C/T | *APPL2 - KCCAT198* | 12q23.3 | 0.407 | 0.309 | 1.78 ( 1.37 - 2.31 ) | 1.92E-05 |
| rs11112453 | 12 | 105689523 | G/T | *APPL2 - KCCAT198* | 12q23.3 | 0.407 | 0.309 | 1.78 ( 1.37 - 2.31 ) | 1.92E-05 |
| rs10861381 | 12 | 105689566 | C/T | *APPL2 - KCCAT198* | 12q23.3 | 0.407 | 0.309 | 1.78 ( 1.37 - 2.31 ) | 1.92E-05 |
| rs7298167 | 12 | 105690259 | T/A | *APPL2 - KCCAT198* | 12q23.3 | 0.407 | 0.308 | 1.79 ( 1.38 - 2.33 ) | 1.50E-05 |
| rs7302933 | 12 | 105691466 | G/C | *APPL2 - KCCAT198* | 12q23.3 | 0.406 | 0.308 | 1.78 ( 1.37 - 2.32 ) | 1.81E-05 |
| rs6539203 | 12 | 105693298 | T/C | *APPL2 - KCCAT198* | 12q23.3 | 0.407 | 0.309 | 1.78 ( 1.37 - 2.31 ) | 1.92E-05 |
| rs7966786 | 12 | 105694513 | C/T | *APPL2 - KCCAT198* | 12q23.3 | 0.407 | 0.309 | 1.78 ( 1.37 - 2.31 ) | 1.92E-05 |
| rs10861382 | 12 | 105694812 | C/T | *APPL2 - KCCAT198* | 12q23.3 | 0.407 | 0.308 | 1.79 ( 1.37 - 2.33 ) | 1.55E-05 |
| rs10861383 | 12 | 105695312 | A/C | *APPL2 - KCCAT198* | 12q23.3 | 0.407 | 0.309 | 1.78 ( 1.37 - 2.31 ) | 1.92E-05 |
| rs10861384 | 12 | 105695459 | G/A | *APPL2 - KCCAT198* | 12q23.3 | 0.409 | 0.309 | 1.79 ( 1.37 - 2.33 ) | 1.52E-05 |
| rs11112457 | 12 | 105695541 | A/T | *APPL2 - KCCAT198* | 12q23.3 | 0.407 | 0.309 | 1.78 ( 1.37 - 2.31 ) | 1.92E-05 |
| rs11112458 | 12 | 105695861 | T/C | *APPL2 - KCCAT198* | 12q23.3 | 0.407 | 0.309 | 1.78 ( 1.37 - 2.31 ) | 1.92E-05 |
| rs10861386 | 12 | 105696895 | A/G | *APPL2 - KCCAT198* | 12q23.3 | 0.404 | 0.306 | 1.76 ( 1.35 - 2.29 ) | 2.54E-05 |
| rs10861387 | 12 | 105696911 | A/T | *APPL2 - KCCAT198* | 12q23.3 | 0.404 | 0.306 | 1.76 ( 1.35 - 2.29 ) | 2.54E-05 |
| rs11112460 | 12 | 105697071 | C/G | *APPL2 - KCCAT198* | 12q23.3 | 0.404 | 0.308 | 1.76 ( 1.35 - 2.29 ) | 2.54E-05 |

| | | | | | | | | | |
|---|---|---|---|---|---|---|---|---|---|
| rs7961016 | 12 | 105697937 | A/T | *KCCAT198* | 12q23.3 | 0.404 | 0.308 | 1.76 ( 1.35 - 2.29 ) | 2.54E-05 |
| rs7300095 | 12 | 105699349 | C/T | *KCCAT198* | 12q23.3 | 0.404 | 0.308 | 1.76 ( 1.35 - 2.29 ) | 2.54E-05 |
| rs2163719 | 12 | 105700039 | G/C | *KCCAT198* | 12q23.3 | 0.404 | 0.307 | 1.77 ( 1.36 - 2.31 ) | 2.00E-05 |
| rs10861390 | 12 | 105701628 | G/A | *KCCAT198* | 12q23.3 | 0.402 | 0.308 | 1.76 ( 1.35 - 2.29 ) | 2.57E-05 |
| rs10861391 | 12 | 105702394 | T/C | *KCCAT198* | 12q23.3 | 0.404 | 0.307 | 1.76 ( 1.35 - 2.29 ) | 2.54E-05 |
| rs7300376 | 12 | 105702824 | G/A | *KCCAT198* | 12q23.3 | 0.404 | 0.308 | 1.76 ( 1.35 - 2.29 ) | 2.54E-05 |
| rs10083056 | 12 | 105702921 | T/C | *KCCAT198* | 12q23.3 | 0.402 | 0.307 | 1.76 ( 1.35 - 2.29 ) | 2.66E-05 |
| rs7300512 | 12 | 105702925 | G/A | *KCCAT198* | 12q23.3 | 0.402 | 0.307 | 1.76 ( 1.35 - 2.29 ) | 2.66E-05 |
| rs10083061 | 12 | 105703041 | A/G | *KCCAT198* | 12q23.3 | 0.404 | 0.308 | 1.76 ( 1.35 - 2.29 ) | 2.54E-05 |
| rs10861393 | 12 | 105703474 | T/C | *KCCAT198* | 12q23.3 | 0.404 | 0.308 | 1.76 ( 1.35 - 2.29 ) | 2.54E-05 |
| rs7305413 | 12 | 105704099 | A/G | *KCCAT198* | 12q23.3 | 0.404 | 0.308 | 1.76 ( 1.35 - 2.29 ) | 2.54E-05 |
| rs10778390 | 12 | 105707106 | T/C | *KCCAT198* | 12q23.3 | 0.404 | 0.307 | 1.76 ( 1.35 - 2.29 ) | 2.54E-05 |
| rs10778391 | 12 | 105707185 | A/G | *KCCAT198* | 12q23.3 | 0.404 | 0.308 | 1.76 ( 1.35 - 2.29 ) | 2.54E-05 |
| rs7299882 | 12 | 105708341 | T/C | *KCCAT198* | 12q23.3 | 0.406 | 0.308 | 1.77 ( 1.36 - 2.3 ) | 2.05E-05 |
| rs10735402 | 12 | 105708888 | G/A | *KCCAT198* | 12q23.3 | 0.409 | 0.308 | 1.8 ( 1.39 - 2.35 ) | 1.17E-05 |
| rs10861397 | 12 | 105714216 | T/C | *KCCAT198* | 12q23.3 | 0.4 | 0.305 | 1.77 ( 1.36 - 2.3 ) | 2.58E-05 |
| rs148792425 | 12 | 105719596 | C/T | *KCCAT198* | 12q23.3 | 0.4 | 0.304 | 1.74 ( 1.34 - 2.27 ) | 3.84E-05 |
| rs838926 | 12 | 125198640 | A/G | *NCOR2 - SCARB1* | 12q24.31 | 0.532 | 0.452 | 1.71 ( 1.33 - 2.2 ) | 3.07E-05 |
| rs11058957 | 12 | 127542092 | A/G | *LINC02405* | 12q24.32 | 0.315 | 0.29 | 1.82 ( 1.36 - 2.44 ) | 5.32E-05 |
| rs116882606 | 12 | 128851012 | T/G | *TMEM132C* | 12q24.32 | 0.093 | 0.058 | 3.33 ( 1.85 - 6.01 ) | 6.46E-05 |
| rs4768982 | 13 | 29159556 | G/A | *FLT1 - POMP* | 13q12.3 | 0.789 | 0.727 | 1.79 ( 1.33 - 2.39 ) | 9.83E-05 |
| rs754304 | 14 | 101358623 | C/G | *MEG8* | 14q32.2 | 0.735 | 0.66 | 1.85 ( 1.41 - 2.44 ) | 1.14E-05 |
| rs74082829 | 14 | 101358831 | G/A | *MEG8* | 14q32.2 | 0.896 | 0.838 | 2.34 ( 1.62 - 3.37 ) | 5.72E-06 |
| rs7160787 | 14 | 101360168 | C/T | *MEG8* | 14q32.2 | 0.906 | 0.845 | 2.43 ( 1.67 - 3.53 ) | 3.67E-06 |
| rs7146460 | 14 | 101361878 | C/T | *MEG8* | 14q32.2 | 0.898 | 0.839 | 2.31 ( 1.6 - 3.33 ) | 8.09E-06 |
| rs55773826 | 15 | 26282223 | C/T | *LINC02346* | 15q12 | 0.938 | 0.896 | 2.57 ( 1.61 - 4.11 ) | 8.18E-05 |

| | | | | | | | | | |
|---|---|---|---|---|---|---|---|---|---|
| rs12908219 | 15 | 46307474 | A/G | *LOC105370802 - SEMA6D* | 15q21.1 | 0.824 | 0.752 | 1.88 ( 1.37 - 2.57 ) | 7.79E-05 |
| rs11074449 | 16 | 20273991 | C/T | *GPR139 - GP2* | 16p12.3 | 0.698 | 0.603 | 1.87 ( 1.43 - 2.43 ) | 3.64E-06 |
| . | 16 | 32833203 | A/G | *TP53TG3 - SLC6A10P* | 16p11.2 | 0.846 | 0.788 | 2.16 ( 1.52 - 3.08 ) | 1.80E-05 |
| rs7215641 | 17 | 16368752 | A/G | *LRRC75A-AS1* | 17p11.2 | 0.822 | 0.752 | 1.91 ( 1.39 - 2.61 ) | 5.38E-05 |
| rs4143830 | 17 | 16370019 | G/A | *LRRC75A-AS1* | 17p11.2 | 0.846 | 0.783 | 1.99 ( 1.43 - 2.77 ) | 4.00E-05 |
| rs11868175 | 17 | 33564479 | A/G | *SLC35G3 - SLFN5* | 17q12 | 0.887 | 0.828 | 2.08 ( 1.44 - 3.01 ) | 8.56E-05 |
| rs11867955 | 17 | 33570468 | T/C | *SLFN5* | 17q12 | 0.889 | 0.829 | 2.14 ( 1.48 - 3.1 ) | 5.03E-05 |
| rs11651968 | 17 | 43333125 | C/T | *SPATA32* | 17q21.31 | 0.689 | 0.619 | 1.72 ( 1.34 - 2.22 ) | 2.77E-05 |
| rs2740757 | 17 | 46647909 | A/G | *HOXB3* | 17q21.32 | 0.494 | 0.423 | 1.65 ( 1.29 - 2.12 ) | 8.46E-05 |
| rs57112168 | 17 | 47994255 | A/C | *FLJ45513 - DLX4* | 17q21.33 | 0.641 | 0.54 | 1.7 ( 1.31 - 2.21 ) | 7.24E-05 |
| rs56011844 | 17 | 47994267 | G/C | *FLJ45513 - DLX4* | 17q21.33 | 0.641 | 0.54 | 1.68 ( 1.29 - 2.18 ) | 9.75E-05 |
| rs72631343 | 17 | 67191270 | C/G | *ABCA10* | 17q24.3 | 0.574 | 0.482 | 1.79 ( 1.38 - 2.34 ) | 1.61E-05 |
| rs60066663 | 17 | 67196143 | T/C | *ABCA10* | 17q24.3 | 0.576 | 0.497 | 1.69 ( 1.3 - 2.2 ) | 9.34E-05 |
| rs12939436 | 17 | 67229514 | T/C | *PRO1804* | 17q24.3 | 0.507 | 0.411 | 1.71 ( 1.31 - 2.22 ) | 6.43E-05 |
| rs817121 | 17 | 67271089 | A/G | *ABCA5* | 17q24.3 | 0.522 | 0.422 | 1.68 ( 1.3 - 2.18 ) | 9.07E-05 |
| rs333937 | 17 | 67319310 | T/C | *ABCA5* | 17q24.3 | 0.6 | 0.501 | 1.77 ( 1.35 - 2.31 ) | 3.07E-05 |
| rs4969282 | 17 | 78828266 | G/T | *RPTOR* | 17q25.3 | 0.841 | 0.784 | 1.92 ( 1.39 - 2.67 ) | 8.17E-05 |
| rs2361221 | 19 | 10638567 | T/G | *S1PR5 - ATG4D* | 19p13.2 | 0.807 | 0.736 | 1.94 ( 1.4 - 2.67 ) | 5.85E-05 |
| rs3093169 | 19 | 15996008 | C/T | *CYP4F2* | 19p13.12 | 0.93 | 0.87 | 2.35 ( 1.53 - 3.6 ) | 8.74E-05 |
| rs2074901 | 19 | 15997422 | A/C | *CYP4F2* | 19p13.12 | 0.931 | 0.871 | 2.42 ( 1.57 - 3.73 ) | 6.36E-05 |
| rs60052980 | 19 | 15999627 | C/T | *CYP4F2* | 19p13.12 | 0.938 | 0.871 | 2.46 ( 1.58 - 3.84 ) | 6.99E-05 |
| rs4646500 | 19 | 16003651 | T/C | *CYP4F2* | 19p13.12 | 0.93 | 0.872 | 2.35 ( 1.53 - 3.61 ) | 9.78E-05 |
| rs3093135 | 19 | 16004371 | A/T | *CYP4F2* | 19p13.12 | 0.93 | 0.872 | 2.35 ( 1.53 - 3.61 ) | 9.78E-05 |
| rs3093134 | 19 | 16004396 | C/G | *CYP4F2* | 19p13.12 | 0.93 | 0.872 | 2.35 ( 1.53 - 3.61 ) | 9.78E-05 |
| rs2365178 | 19 | 16005003 | C/T | *CYP4F2* | 19p13.12 | 0.93 | 0.872 | 2.35 ( 1.53 - 3.61 ) | 9.78E-05 |
| rs3093124 | 19 | 16005232 | A/G | *CYP4F2* | 19p13.12 | 0.93 | 0.872 | 2.35 ( 1.53 - 3.61 ) | 9.78E-05 |

| | | | | | | | | | |
|---|---|---|---|---|---|---|---|---|---|
| rs3093122 | 19 | 16005276 | G/C | *CYP4F2* | 19p13.12 | 0.93 | 0.872 | 2.35 ( 1.53 - 3.61 ) | 9.78E-05 |
| rs3093121 | 19 | 16005277 | G/T | *CYP4F2* | 19p13.12 | 0.93 | 0.872 | 2.35 ( 1.53 - 3.61 ) | 9.78E-05 |
| rs3093120 | 19 | 16005296 | C/T | *CYP4F2* | 19p13.12 | 0.93 | 0.872 | 2.35 ( 1.53 - 3.61 ) | 9.78E-05 |
| rs3093115 | 19 | 16005615 | A/T | *CYP4F2* | 19p13.12 | 0.93 | 0.873 | 2.35 ( 1.53 - 3.61 ) | 9.78E-05 |
| rs984692 | 19 | 16006074 | T/A | *CYP4F2* | 19p13.12 | 0.93 | 0.872 | 2.35 ( 1.53 - 3.61 ) | 9.78E-05 |
| rs2215093 | 19 | 16006101 | T/C | *CYP4F2* | 19p13.12 | 0.93 | 0.872 | 2.35 ( 1.53 - 3.61 ) | 9.78E-05 |
| rs8109064 | 19 | 16006178 | A/G | *CYP4F2* | 19p13.12 | 0.93 | 0.872 | 2.35 ( 1.53 - 3.61 ) | 9.78E-05 |
| rs3093114 | 19 | 16006413 | G/A | *CYP4F2* | 19p13.12 | 0.93 | 0.872 | 2.35 ( 1.53 - 3.61 ) | 9.78E-05 |
| rs3093112 | 19 | 16006611 | T/C | *CYP4F2* | 19p13.12 | 0.931 | 0.872 | 2.42 ( 1.58 - 3.72 ) | 5.52E-05 |
| rs8112642 | 19 | 16006854 | A/C | *CYP4F2* | 19p13.12 | 0.933 | 0.872 | 2.43 ( 1.57 - 3.74 ) | 6.20E-05 |
| rs2074902 | 19 | 16008099 | T/C | *CYP4F2* | 19p13.12 | 0.93 | 0.872 | 2.35 ( 1.53 - 3.61 ) | 9.78E-05 |
| rs3093100 | 19 | 16008469 | C/G | *CYP4F2* | 19p13.12 | 0.93 | 0.872 | 2.35 ( 1.53 - 3.61 ) | 9.78E-05 |
| rs114635268 | 19 | 16077628 | A/G | *OR10H4 - LINC00661* | 19p13.12 | 0.922 | 0.872 | 2.33 ( 1.53 - 3.54 ) | 7.54E-05 |
| rs6051348 | 20 | 2717526 | T/C | *EBF4* | 20p13 | 0.213 | 0.15 | 2.01 ( 1.42 - 2.83 ) | 7.13E-05 |
| rs2875959 | 20 | 4593976 | A/G | *ADRA1D - PRNP* | 20p13 | 0.615 | 0.563 | 1.63 ( 1.27 - 2.08 ) | 9.51E-05 |
| rs117624310 | 20 | 39153161 | T/C | *LINC01370 - MAFB* | 20q12 | 0.933 | 0.875 | 2.6 ( 1.69 - 3.98 ) | 1.23E-05 |
| rs12625267 | 20 | 45803936 | C/G | *EYA2* | 20q13.12 | 0.298 | 0.217 | 1.8 ( 1.34 - 2.42 ) | 9.27E-05 |
| rs2281366 | 20 | 45808926 | A/G | *EYA2* | 20q13.12 | 0.302 | 0.219 | 1.84 ( 1.37 - 2.47 ) | 5.69E-05 |
| rs2073171 | 20 | 45809082 | C/T | *EYA2* | 20q13.12 | 0.302 | 0.217 | 1.83 ( 1.36 - 2.45 ) | 6.04E-05 |
| rs80071852 | 20 | 48957588 | C/G | *LINC01271 - PTPN1* | 20q13.13 | 0.117 | 0.058 | 2.57 ( 1.61 - 4.11 ) | 7.80E-05 |
| rs79969681 | 20 | 48964930 | A/G | *LINC01271 - PTPN1* | 20q13.13 | 0.115 | 0.057 | 2.59 ( 1.62 - 4.17 ) | 8.11E-05 |
| rs116760543 | 20 | 48966330 | A/G | *LINC01271 - PTPN1* | 20q13.13 | 0.117 | 0.057 | 2.65 ( 1.64 - 4.26 ) | 6.04E-05 |
| rs138000473 | 20 | 48968450 | T/C | *LINC01271 - PTPN1* | 20q13.13 | 0.115 | 0.057 | 2.52 ( 1.58 - 4.03 ) | 9.82E-05 |
| rs72626599 | 20 | 56320267 | G/A | *NKILA - MIR4532* | 20q13.31 | 0.826 | 0.757 | 1.86 ( 1.37 - 2.52 ) | 6.21E-05 |
| rs11088462 | 21 | 40354531 | T/G | *LINC01700 - PSMG1* | 21q22.2 | 0.374 | 0.32 | 1.73 ( 1.32 - 2.27 ) | 6.95E-05 |
| rs423158 | 22 | 18053496 | A/G | *SLC25A18* | 22q11.21 | 0.268 | 0.207 | 1.92 ( 1.4 - 2.64 ) | 6.04E-05 |

| | | | | | | | | | |
|---|---|---|---|---|---|---|---|---|---|
| rs7284558 | 22 | 48885052 | C/T | *FAM19A5* | 22q13.32 | 0.433 | 0.328 | 1.87 ( 1.45 - 2.42 ) | 1.72E-06 |
| rs12858236 | 23 | 4846592 | A/G | *NONE - NONE* | . | 0.844 | 0.721 | 2.52 ( 1.64 - 3.88 ) | 2.71E-05 |
| rs140372616 | 23 | 146675912 | A/G | *NONE - NONE* | . | 0.115 | 0.059 | 5.27 ( 2.32 - 11.98 ) | 7.38E-05 |

[a] The positions of SNVs are based on NCBI human genome reference sequence Build 37

[b] A1 is risk-associated allele, and A2 is non-risk-associated allele.

**Appendix Table 2 SNVs with $P < 1\times10^{-4}$ in the G-H group of GWAS discovery stage**

| | | Position | | | | Frequency of A1 | | | |
|---|---|---|---|---|---|---|---|---|---|
| **Rs id** | **Chr** | **Bp[a]** | **A1/A2[b]** | **Gene** | **Locus** | **Cases** | **Controls** | **OR (95% CI)** | **P value** |
| rs367520 | 1 | 25399781 | T/C | *MIR4425 - SYF2* | 1p36.11 | 0.411 | 0.292 | 1.84 ( 1.36 - 2.49 ) | 8.84E-05 |
| rs16830099 | 1 | 43406701 | A/G | *SLC2A1* | 1p34.2 | 0.196 | 0.107 | 2.37 ( 1.54 - 3.63 ) | 8.26E-05 |
| rs57247989 | 1 | 43414370 | G/C | *SLC2A1* | 1p34.2 | 0.211 | 0.117 | 2.36 ( 1.54 - 3.6 ) | 6.91E-05 |
| rs58062906 | 1 | 43438399 | T/C | *SLC2A1 - AS1* | 1p34.2 | 0.185 | 0.101 | 2.46 ( 1.57 - 3.87 ) | 9.54E-05 |
| rs879688782 | 1 | 52348092 | C/A | *NRDC - RAB3B* | 1p32.3 | 0.795 | 0.683 | 2.49 ( 1.67 - 3.71 ) | 6.60E-06 |
| rs12129986 | 1 | 94399210 | A/G | *GCLM - ABCA4* | 1p22.1 | 0.9 | 0.798 | 2.25 ( 1.51 - 3.35 ) | 6.82E-05 |
| rs1190101854 | 1 | 1.55E+08 | T/* | . | . | 0.821 | 0.708 | 2.4 ( 1.6 - 3.59 ) | 2.10E-05 |
| rs58836406 | 1 | 2.39E+08 | C/T | *LINC01139 - CHRM3* | 1q43 | 0.528 | 0.391 | 1.84 ( 1.37 - 2.47 ) | 4.67E-05 |
| rs6697193 | 1 | 2.49E+08 | C/T | *ZNF692 - PGBD2* | 1q44 | 0.824 | 0.688 | 2.04 ( 1.46 - 2.83 ) | 2.36E-05 |
| rs4335411 | 1 | 2.49E+08 | A/G | *ZNF692 - PGBD2* | 1q44 | 0.809 | 0.667 | 2.1 ( 1.52 - 2.92 ) | 8.37E-06 |
| rs62158971 | 2 | 57622282 | C/T | *CCDC85A - VRK2* | 2p16.1 | 0.924 | 0.828 | 2.58 ( 1.62 - 4.12 ) | 6.59E-05 |
| rs146136095 | 2 | 57627526 | G/T | *CCDC85A - VRK2* | 2p16.1 | 0.924 | 0.831 | 2.56 ( 1.6 - 4.11 ) | 8.85E-05 |
| rs4416200 | 2 | 57636894 | A/C | *CCDC85A - VRK2* | 2p16.1 | 0.926 | 0.828 | 2.68 ( 1.67 - 4.28 ) | 3.99E-05 |
| rs12623520 | 2 | 57668554 | C/T | *CCDC85A - VRK2* | 2p16.1 | 0.924 | 0.828 | 2.61 ( 1.63 - 4.16 ) | 5.82E-05 |
| rs17048827 | 2 | 57669359 | G/C | *CCDC85A - VRK2* | 2p16.1 | 0.924 | 0.828 | 2.61 ( 1.63 - 4.16 ) | 5.82E-05 |
| rs17048834 | 2 | 57689942 | C/T | *CCDC85A - VRK2* | 2p16.1 | 0.924 | 0.828 | 2.61 ( 1.63 - 4.16 ) | 5.82E-05 |
| rs72816507 | 2 | 57693832 | A/C | *CCDC85A - VRK2* | 2p16.1 | 0.924 | 0.828 | 2.61 ( 1.63 - 4.16 ) | 5.82E-05 |

| | | | | | | | | | |
|---|---|---|---|---|---|---|---|---|---|
| rs1914755 | 2 | 57706827 | C/T | *CCDC85A - VRK2* | 2p16.1 | 0.906 | 0.798 | 2.38 ( 1.56 - 3.64 ) | 6.17E-05 |
| rs3927303 | 2 | 2.03E+08 | T/C | *BMPR2* | 2q33.2 | 0.88 | 0.794 | 2.33 ( 1.52 - 3.57 ) | 9.50E-05 |
| rs163579 | 3 | 3101065 | C/T | *CNTN4-AS1* | 3p26.2 | 0.833 | 0.73 | 2.1 ( 1.47 - 3 ) | 4.81E-05 |
| rs9876659 | 3 | 3101111 | A/G | *CNTN4-AS1* | 3p26.2 | 0.828 | 0.716 | 2.16 ( 1.52 - 3.08 ) | 2.02E-05 |
| rs6776625 | 3 | 12864664 | C/T | *CAND2* | 3p25.2 | 0.611 | 0.483 | 1.84 ( 1.37 - 2.46 ) | 4.81E-05 |
| rs929397437 | 3 | 22321858 | T/* | . | . | 0.933 | 0.85 | 2.8 ( 1.7 - 4.59 ) | 4.90E-05 |
| rs112586249 | 3 | 54156146 | G/T | *CACNA2D3* | 3p21.1 | 0.567 | 0.421 | 1.76 ( 1.35 - 2.3 ) | 2.76E-05 |
| rs6777548 | 3 | 81427224 | G/A | *LINC02027 - GBE1* | 3p12.2 | 0.706 | 0.566 | 1.79 ( 1.34 - 2.38 ) | 7.91E-05 |
| rs28625332 | 4 | 17778944 | G/C | *FAM184B* | 4p15.32 | 0.744 | 0.62 | 1.87 ( 1.38 - 2.53 ) | 5.72E-05 |
| rs887778 | 4 | 27295829 | A/G | *LINC02261 - MIR4275* | 4p15.2 | 0.763 | 0.648 | 1.91 ( 1.38 - 2.63 ) | 8.08E-05 |
| rs2080110 | 4 | 27303327 | A/G | *LINC02261 - MIR4275* | 4p15.2 | 0.767 | 0.65 | 1.92 ( 1.39 - 2.65 ) | 6.66E-05 |
| rs16879151 | 4 | 27303909 | C/G | *LINC02261 - MIR4275* | 4p15.2 | 0.767 | 0.65 | 1.9 ( 1.38 - 2.62 ) | 8.08E-05 |
| rs3846339 | 4 | 27305166 | T/C | *LINC02261 - MIR4275* | 4p15.2 | 0.763 | 0.648 | 1.9 ( 1.38 - 2.61 ) | 8.53E-05 |
| rs6814847 | 4 | 39770834 | */T | . | . | 0.819 | 0.7 | 2.11 ( 1.48 - 3 ) | 3.76E-05 |
| rs2725220 | 4 | 88959922 | C/G | *PKD2* | 4q22.1 | 0.42 | 0.262 | 1.97 ( 1.45 - 2.68 ) | 1.61E-05 |
| rs2725219 | 4 | 88959961 | C/T | *PKD2* | 4q22.1 | 0.42 | 0.26 | 2 ( 1.47 - 2.73 ) | 1.07E-05 |
| rs2725218 | 4 | 88959968 | C/G | *PKD2* | 4q22.1 | 0.422 | 0.262 | 1.98 ( 1.45 - 2.69 ) | 1.39E-05 |
| rs2725217 | 4 | 88960258 | T/A | *PKD2* | 4q22.1 | 0.428 | 0.262 | 2.08 ( 1.53 - 2.85 ) | 3.95E-06 |
| rs2725216 | 4 | 88960579 | G/A | *PKD2* | 4q22.1 | 0.437 | 0.295 | 1.85 ( 1.36 - 2.5 ) | 8.36E-05 |
| rs2725215 | 4 | 88961571 | T/C | *PKD2* | 4q22.1 | 0.43 | 0.262 | 2.11 ( 1.54 - 2.88 ) | 3.08E-06 |
| rs2728108 | 4 | 88961736 | C/A | *PKD2* | 4q22.1 | 0.437 | 0.295 | 1.85 ( 1.36 - 2.5 ) | 8.36E-05 |
| rs17013739 | 4 | 88963255 | G/A | *PKD2* | 4q22.1 | 0.904 | 0.787 | 2.67 ( 1.76 - 4.04 ) | 3.94E-06 |
| rs10031265 | 4 | 88968343 | A/G | *PKD2* | 4q22.1 | 0.898 | 0.74 | 3.29 ( 2.2 - 4.93 ) | 6.78E-09 |
| rs2728107 | 4 | 88968488 | T/C | *PKD2* | 4q22.1 | 0.435 | 0.265 | 2.17 ( 1.58 - 2.98 ) | 1.49E-06 |
| rs2725211 | 4 | 88970375 | T/C | *PKD2* | 4q22.1 | 0.435 | 0.265 | 2.17 ( 1.58 - 2.98 ) | 1.49E-06 |
| rs2728104 | 4 | 88973006 | C/T | *PKD2* | 4q22.1 | 0.441 | 0.292 | 1.95 ( 1.43 - 2.66 ) | 2.29E-05 |

| | | | | | | | | | |
|---|---|---|---|---|---|---|---|---|---|
| rs2728099 | 4 | 88975738 | C/T | *PKD2* | 4q22.1 | 0.433 | 0.265 | 2.13 ( 1.56 - 2.92 ) | 2.07E-06 |
| rs2728134 | 4 | 88978139 | G/T | *PKD2* | 4q22.1 | 0.433 | 0.264 | 2.13 ( 1.56 - 2.91 ) | 2.22E-06 |
| rs74901820 | 4 | 88981537 | G/A | *PKD2* | 4q22.1 | 0.906 | 0.792 | 2.7 ( 1.77 - 4.11 ) | 3.63E-06 |
| rs74668466 | 4 | 88981837 | A/G | *PKD2* | 4q22.1 | 0.906 | 0.787 | 2.76 ( 1.82 - 4.2 ) | 1.99E-06 |
| rs117007073 | 4 | 88987915 | C/T | *PKD2* | 4q22.1 | 0.906 | 0.784 | 2.82 ( 1.85 - 4.29 ) | 1.23E-06 |
| rs74933714 | 4 | 88990386 | T/G | *PKD2* | 4q22.1 | 0.906 | 0.79 | 2.69 ( 1.78 - 4.07 ) | 2.91E-06 |
| rs2725204 | 4 | 88992813 | G/T | *PKD2* | 4q22.1 | 0.437 | 0.27 | 2.09 ( 1.53 - 2.85 ) | 3.29E-06 |
| rs2725202 | 4 | 88997876 | A/G | *PKD2* | 4q22.1 | 0.437 | 0.27 | 2.09 ( 1.53 - 2.85 ) | 3.36E-06 |
| rs2728126 | 4 | 88999222 | A/T | *PKD2* | 4q22.1 | 0.443 | 0.27 | 2.12 ( 1.56 - 2.89 ) | 1.83E-06 |
| rs17013802 | 4 | 89001185 | T/A | *PKD2 - ABCG2* | 4q22.1 | 0.904 | 0.792 | 2.62 ( 1.73 - 3.97 ) | 5.60E-06 |
| rs2728125 | 4 | 89001893 | G/A | *PKD2 - ABCG2* | 4q22.1 | 0.443 | 0.273 | 2.12 ( 1.55 - 2.89 ) | 2.20E-06 |
| rs2728124 | 4 | 89006160 | A/T | *PKD2 - ABCG2* | 4q22.1 | 0.456 | 0.292 | 2.05 ( 1.51 - 2.78 ) | 3.85E-06 |
| rs17013810 | 4 | 89008506 | A/G | *PKD2 - ABCG2* | 4q22.1 | 0.909 | 0.792 | 2.97 ( 1.93 - 4.57 ) | 7.17E-07 |
| rs2725269 | 4 | 89009006 | T/C | *PKD2 - ABCG2* | 4q22.1 | 0.435 | 0.284 | 1.87 ( 1.39 - 2.51 ) | 3.28E-05 |
| rs142474948 | 4 | 89010900 | A/G | *ABCG2* | 4q22.1 | 0.915 | 0.8 | 3 ( 1.94 - 4.65 ) | 8.07E-07 |
| rs2725268 | 4 | 89010983 | G/A | *ABCG2* | 4q22.1 | 0.444 | 0.284 | 1.99 ( 1.47 - 2.68 ) | 7.55E-06 |
| rs1448784 | 4 | 89012320 | A/G | *ABCG2* | 4q22.1 | 0.878 | 0.757 | 2.69 ( 1.8 - 4 ) | 1.17E-06 |
| rs4148160 | 4 | 89015090 | C/T | *ABCG2* | 4q22.1 | 0.909 | 0.787 | 3.15 ( 2.02 - 4.9 ) | 3.85E-07 |
| rs2231164 | 4 | 89015857 | C/T | *ABCG2* | 4q22.1 | 0.646 | 0.486 | 2.04 ( 1.5 - 2.78 ) | 5.02E-06 |
| rs1383585 | 4 | 89019735 | G/A | *ABCG2* | 4q22.1 | 0.472 | 0.273 | 2.29 ( 1.7 - 3.11 ) | 7.55E-08 |
| rs2231156 | 4 | 89020427 | A/C | *ABCG2* | 4q22.1 | 0.472 | 0.29 | 2.19 ( 1.61 - 2.96 ) | 4.76E-07 |
| rs4148157 | 4 | 89020934 | A/G | *ABCG2* | 4q22.1 | 0.472 | 0.29 | 2.19 ( 1.61 - 2.96 ) | 4.76E-07 |
| rs4693924 | 4 | 89023224 | A/G | *ABCG2* | 4q22.1 | 0.472 | 0.29 | 2.19 ( 1.61 - 2.96 ) | 4.76E-07 |
| rs34455506 | 4 | 89024220 | G/A | *ABCG2* | 4q22.1 | 0.909 | 0.787 | 3.15 ( 2.02 - 4.9 ) | 3.85E-07 |
| rs76979899 | 4 | 89025241 | T/C | *ABCG2* | 4q22.1 | 0.47 | 0.284 | 2.22 ( 1.64 - 3 ) | 2.92E-07 |
| rs2725263 | 4 | 89026428 | C/A | *ABCG2* | 4q22.1 | 0.737 | 0.59 | 1.98 ( 1.46 - 2.69 ) | 1.11E-05 |

| | | | | | | | | | |
|---|---|---|---|---|---|---|---|---|---|
| rs7681519 | 4 | 89027840 | G/C | *ABCG2* | 4q22.1 | 0.909 | 0.787 | 3.15 ( 2.02 - 4.9 ) | 3.85E-07 |
| rs2231148 | 4 | 89028478 | T/A | *ABCG2* | 4q22.1 | 0.909 | 0.787 | 3.15 ( 2.02 - 4.9 ) | 3.85E-07 |
| rs2054576 | 4 | 89028775 | G/A | *ABCG2* | 4q22.1 | 0.47 | 0.29 | 2.16 ( 1.6 - 2.93 ) | 6.28E-07 |
| rs34472643 | 4 | 89029866 | G/A | *ABCG2* | 4q22.1 | 0.894 | 0.781 | 2.72 ( 1.78 - 4.16 ) | 4.07E-06 |
| rs12505410 | 4 | 89030841 | T/G | *ABCG2* | 4q22.1 | 0.876 | 0.727 | 2.68 ( 1.84 - 3.89 ) | 2.33E-07 |
| rs2622621 | 4 | 89030920 | G/C | *ABCG2* | 4q22.1 | 0.778 | 0.607 | 2.3 ( 1.67 - 3.17 ) | 3.21E-07 |
| rs11943824 | 4 | 89031519 | A/G | *ABCG2* | 4q22.1 | 0.876 | 0.73 | 2.61 ( 1.8 - 3.79 ) | 4.34E-07 |
| rs11939579 | 4 | 89031586 | T/G | *ABCG2* | 4q22.1 | 0.876 | 0.732 | 2.59 ( 1.78 - 3.76 ) | 5.59E-07 |
| rs1481013 | 4 | 89032679 | C/T | *ABCG2* | 4q22.1 | 0.818 | 0.66 | 2.43 ( 1.71 - 3.46 ) | 7.49E-07 |
| rs2725261 | 4 | 89036353 | T/C | *ABCG2* | 4q22.1 | 0.786 | 0.607 | 2.16 ( 1.6 - 2.92 ) | 5.33E-07 |
| rs1481012 | 4 | 89039082 | G/A | *ABCG2* | 4q22.1 | 0.598 | 0.336 | 3.23 ( 2.33 - 4.5 ) | 3.16E-12 |
| rs2231146 | 4 | 89039500 | T/C | *ABCG2* | 4q22.1 | 0.93 | 0.806 | 3.42 ( 2.16 - 5.41 ) | 1.70E-07 |
| rs2231145 | 4 | 89039584 | T/C | *ABCG2* | 4q22.1 | 0.93 | 0.806 | 3.42 ( 2.16 - 5.41 ) | 1.70E-07 |
| rs1871744 | 4 | 89039629 | T/C | *ABCG2* | 4q22.1 | 0.9 | 0.751 | 3.16 ( 2.1 - 4.75 ) | 3.79E-08 |
| rs2448795 | 4 | 89040850 | A/G | *ABCG2* | 4q22.1 | 0.83 | 0.691 | 2.04 ( 1.46 - 2.83 ) | 2.30E-05 |
| rs117195876 | 4 | 89041399 | T/C | *ABCG2* | 4q22.1 | 0.93 | 0.806 | 3.42 ( 2.16 - 5.41 ) | 1.70E-07 |
| rs117815474 | 4 | 89041878 | C/T | *ABCG2* | 4q22.1 | 0.93 | 0.806 | 3.42 ( 2.16 - 5.41 ) | 1.70E-07 |
| rs45499402 | 4 | 89043634 | C/G | *ABCG2* | 4q22.1 | 0.613 | 0.344 | 3.33 ( 2.4 - 4.63 ) | 8.41E-13 |
| rs2170290 | 4 | 89044067 | T/C | *ABCG2* | 4q22.1 | 0.9 | 0.751 | 3.16 ( 2.1 - 4.75 ) | 3.79E-08 |
| rs149027545 | 4 | 89044180 | C/G | *ABCG2* | 4q22.1 | 0.613 | 0.344 | 3.33 ( 2.4 - 4.63 ) | 8.41E-13 |
| rs138409370 | 4 | 89044312 | T/A | *ABCG2* | 4q22.1 | 0.613 | 0.344 | 3.33 ( 2.4 - 4.63 ) | 8.41E-13 |
| rs75544042 | 4 | 89045331 | A/G | *ABCG2* | 4q22.1 | 0.613 | 0.344 | 3.33 ( 2.4 - 4.63 ) | 8.41E-13 |
| rs3102038 | 4 | 89045426 | G/A | *ABCG2* | 4q22.1 | 0.83 | 0.691 | 2.04 ( 1.46 - 2.83 ) | 2.30E-05 |
| rs141471965 | 4 | 89046202 | T/C | *ABCG2* | 4q22.1 | 0.613 | 0.344 | 3.33 ( 2.4 - 4.63 ) | 8.41E-13 |
| rs117607915 | 4 | 89046617 | G/A | *ABCG2* | 4q22.1 | 0.931 | 0.809 | 3.44 ( 2.17 - 5.47 ) | 1.69E-07 |
| rs74904971 | 4 | 89050026 | A/C | *ABCG2* | 4q22.1 | 0.613 | 0.344 | 3.33 ( 2.4 - 4.63 ) | 8.41E-13 |

| | | | | | | | | | |
|---|---|---|---|---|---|---|---|---|---|
| rs2725256 | 4 | 89050998 | A/G | *ABCG2* | 4q22.1 | 0.83 | 0.691 | 2.04 ( 1.46 - 2.83 ) | 2.30E-05 |
| rs2231142 | 4 | 89052323 | T/G | *ABCG2* | 4q22.1 | 0.613 | 0.344 | 3.33 ( 2.4 - 4.63 ) | 8.41E-13 |
| rs4148155 | 4 | 89054667 | G/A | *ABCG2* | 4q22.1 | 0.611 | 0.342 | 3.32 ( 2.39 - 4.61 ) | 7.89E-13 |
| rs3114017 | 4 | 89055194 | C/T | *ABCG2* | 4q22.1 | 0.824 | 0.678 | 2.08 ( 1.51 - 2.88 ) | 9.01E-06 |
| rs4148153 | 4 | 89056715 | G/A | *ABCG2* | 4q22.1 | 0.928 | 0.8 | 3.43 ( 2.17 - 5.41 ) | 1.20E-07 |
| rs2725254 | 4 | 89057664 | C/T | *ABCG2* | 4q22.1 | 0.832 | 0.697 | 2.03 ( 1.46 - 2.83 ) | 2.43E-05 |
| rs2929060 | 4 | 89058220 | A/C | *ABCG2* | 4q22.1 | 0.832 | 0.691 | 2.06 ( 1.48 - 2.85 ) | 1.57E-05 |
| rs1564481 | 4 | 89061265 | C/T | *ABCG2* | 4q22.1 | 0.83 | 0.694 | 2.05 ( 1.48 - 2.85 ) | 1.95E-05 |
| rs77377473 | 4 | 89061802 | A/G | *ABCG2* | 4q22.1 | 0.937 | 0.828 | 3.31 ( 2.05 - 5.34 ) | 9.20E-07 |
| rs2725252 | 4 | 89061910 | C/A | *ABCG2* | 4q22.1 | 0.793 | 0.615 | 2.24 ( 1.64 - 3.08 ) | 5.08E-07 |
| rs72554039 | 4 | 89063354 | G/A | *ABCG2* | 4q22.1 | 0.937 | 0.831 | 3.23 ( 2 - 5.21 ) | 1.53E-06 |
| rs3109823 | 4 | 89064602 | T/C | *ABCG2* | 4q22.1 | 0.926 | 0.814 | 2.93 ( 1.86 - 4.61 ) | 3.63E-06 |
| rs2725248 | 4 | 89068007 | A/C | *ABCG2* | 4q22.1 | 0.926 | 0.814 | 2.6 ( 1.68 - 4.02 ) | 1.62E-05 |
| rs2622625 | 4 | 89068737 | C/T | *ABCG2* | 4q22.1 | 0.924 | 0.822 | 2.64 ( 1.68 - 4.14 ) | 2.37E-05 |
| rs6821607 | 4 | 89074808 | G/A | *ABCG2* | 4q22.1 | 0.922 | 0.809 | 2.91 ( 1.86 - 4.55 ) | 2.82E-06 |
| rs3109822 | 4 | 89075223 | C/T | *ABCG2* | 4q22.1 | 0.92 | 0.806 | 2.77 ( 1.79 - 4.29 ) | 4.75E-06 |
| rs145778965 | 4 | 89075239 | C/T | *ABCG2* | 4q22.1 | 0.422 | 0.224 | 2.6 ( 1.87 - 3.63 ) | 1.82E-08 |
| rs2622604 | 4 | 89078924 | C/T | *ABCG2* | 4q22.1 | 0.922 | 0.814 | 2.76 ( 1.77 - 4.32 ) | 8.35E-06 |
| rs3114019 | 4 | 89081441 | T/C | *ABCG2* | 4q22.1 | 0.922 | 0.812 | 2.84 ( 1.82 - 4.44 ) | 4.56E-06 |
| rs2622606 | 4 | 89084381 | T/A | *ABCG2* | 4q22.1 | 0.922 | 0.812 | 2.84 ( 1.82 - 4.44 ) | 4.56E-06 |
| rs76368528 | 4 | 89092524 | G/T | *ABCG2* | 4q22.1 | 0.9 | 0.8 | 2.29 ( 1.51 - 3.47 ) | 8.67E-05 |
| rs183997620 | 4 | 89100289 | C/G | *ABCG2* | 4q22.1 | 0.328 | 0.18 | 2.21 ( 1.57 - 3.12 ) | 5.82E-06 |
| rs114086764 | 4 | 89118728 | G/A | *ABCG2* | 4q22.1 | 0.878 | 0.757 | 2.42 ( 1.64 - 3.56 ) | 7.62E-06 |
| rs143861620 | 4 | 89121226 | C/T | *ABCG2* | 4q22.1 | 0.837 | 0.694 | 2.28 ( 1.6 - 3.25 ) | 4.92E-06 |
| rs115232777 | 4 | 89121335 | A/C | *ABCG2* | 4q22.1 | 0.837 | 0.7 | 2.2 ( 1.54 - 3.13 ) | 1.24E-05 |
| rs116549154 | 4 | 89121743 | C/T | *ABCG2* | 4q22.1 | 0.837 | 0.697 | 2.26 ( 1.58 - 3.21 ) | 6.77E-06 |

| | | | | | | | | | |
|---|---|---|---|---|---|---|---|---|---|
| rs4546214 | 4 | 89122311 | G/C | *ABCG2* | 4q22.1 | 0.857 | 0.71 | 2.27 ( 1.59 - 3.23 ) | 5.43E-06 |
| rs4693930 | 4 | 89122833 | A/G | *ABCG2* | 4q22.1 | 0.835 | 0.691 | 2.3 ( 1.61 - 3.27 ) | 4.27E-06 |
| rs13108819 | 4 | 89123925 | T/C | *ABCG2* | 4q22.1 | 0.835 | 0.691 | 2.3 ( 1.61 - 3.27 ) | 4.27E-06 |
| rs12500185 | 4 | 89124563 | T/C | *ABCG2* | 4q22.1 | 0.835 | 0.691 | 2.3 ( 1.61 - 3.27 ) | 4.27E-06 |
| rs11097184 | 4 | 89125311 | C/T | *ABCG2* | 4q22.1 | 0.835 | 0.691 | 2.3 ( 1.61 - 3.27 ) | 4.27E-06 |
| rs144292084 | 4 | 89125424 | T/A | *ABCG2* | 4q22.1 | 0.837 | 0.694 | 2.28 ( 1.6 - 3.25 ) | 4.92E-06 |
| rs74943514 | 4 | 89125932 | G/C | *ABCG2* | 4q22.1 | 0.837 | 0.694 | 2.28 ( 1.6 - 3.25 ) | 4.92E-06 |
| rs12511059 | 4 | 89126193 | T/C | *ABCG2* | 4q22.1 | 0.833 | 0.691 | 2.24 ( 1.57 - 3.18 ) | 7.20E-06 |
| rs77180571 | 4 | 89126434 | C/T | *ABCG2* | 4q22.1 | 0.837 | 0.694 | 2.28 ( 1.6 - 3.25 ) | 4.92E-06 |
| rs7672396 | 4 | 89126767 | T/C | *ABCG2* | 4q22.1 | 0.835 | 0.691 | 2.28 ( 1.6 - 3.23 ) | 4.44E-06 |
| rs66508773 | 4 | 89127082 | T/C | *ABCG2* | 4q22.1 | 0.835 | 0.691 | 2.3 ( 1.61 - 3.27 ) | 4.27E-06 |
| rs118013835 | 4 | 89128183 | A/G | *ABCG2* | 4q22.1 | 0.174 | 0.066 | 2.85 ( 1.75 - 4.66 ) | 2.91E-05 |
| rs11935352 | 4 | 89128413 | C/T | *ABCG2* | 4q22.1 | 0.835 | 0.691 | 2.3 ( 1.61 - 3.27 ) | 4.27E-06 |
| rs12646307 | 4 | 89129256 | A/T | *ABCG2* | 4q22.1 | 0.837 | 0.694 | 2.28 ( 1.6 - 3.25 ) | 4.92E-06 |
| rs13127017 | 4 | 89135288 | T/C | *ABCG2* | 4q22.1 | 0.839 | 0.683 | 2.41 ( 1.7 - 3.42 ) | 9.29E-07 |
| rs1986013 | 4 | 89137901 | G/T | *ABCG2* | 4q22.1 | 0.839 | 0.688 | 2.34 ( 1.65 - 3.34 ) | 2.26E-06 |
| rs4693935 | 4 | 89139275 | G/A | *ABCG2* | 4q22.1 | 0.839 | 0.688 | 2.34 ( 1.65 - 3.34 ) | 2.26E-06 |
| rs4693207 | 4 | 89139659 | T/A | *ABCG2* | 4q22.1 | 0.841 | 0.691 | 2.36 ( 1.65 - 3.37 ) | 2.18E-06 |
| rs1383587 | 4 | 89141574 | T/C | *ABCG2* | 4q22.1 | 0.791 | 0.626 | 2.32 ( 1.66 - 3.23 ) | 6.61E-07 |
| rs13119417 | 4 | 89143112 | G/A | *ABCG2* | 4q22.1 | 0.793 | 0.626 | 2.34 ( 1.68 - 3.26 ) | 5.13E-07 |
| rs1904903 | 4 | 89145274 | T/A | *ABCG2* | 4q22.1 | 0.791 | 0.626 | 2.32 ( 1.66 - 3.23 ) | 6.75E-07 |
| rs1074839 | 4 | 89145548 | T/C | *ABCG2* | 4q22.1 | 0.789 | 0.626 | 2.27 ( 1.63 - 3.15 ) | 1.09E-06 |
| rs2046132 | 4 | 89149026 | G/C | *ABCG2* | 4q22.1 | 0.789 | 0.626 | 2.27 ( 1.64 - 3.16 ) | 1.01E-06 |
| rs13107048 | 4 | 89150075 | C/T | *ABCG2* | 4q22.1 | 0.791 | 0.623 | 2.33 ( 1.68 - 3.25 ) | 4.89E-07 |
| rs111955262 | 4 | 89150226 | T/C | *ABCG2* | 4q22.1 | 0.832 | 0.675 | 2.31 ( 1.64 - 3.26 ) | 1.85E-06 |
| rs72554040 | 4 | 89152324 | G/A | *ABCG2* | 4q22.1 | 0.804 | 0.648 | 2.23 ( 1.61 - 3.11 ) | 1.87E-06 |

| | | | | | | | | | |
|---|---|---|---|---|---|---|---|---|---|
| rs61046194 | 4 | 89154025 | A/T | *ABCG2 - PPM1K* | 4q22.1 | 0.943 | 0.855 | 2.75 ( 1.67 - 4.53 ) | 7.24E-05 |
| rs117104615 | 4 | 89154065 | G/T | *ABCG2 - PPM1K* | 4q22.1 | 0.839 | 0.688 | 2.21 ( 1.57 - 3.11 ) | 6.11E-06 |
| rs6846256 | 4 | 89154111 | G/T | *ABCG2 - PPM1K* | 4q22.1 | 0.809 | 0.653 | 2.24 ( 1.6 - 3.12 ) | 2.08E-06 |
| rs12641384 | 4 | 89155237 | C/G | *ABCG2 - PPM1K* | 4q22.1 | 0.837 | 0.667 | 2.45 ( 1.74 - 3.45 ) | 3.05E-07 |
| rs13328043 | 4 | 89155971 | A/C | *ABCG2 - PPM1K* | 4q22.1 | 0.788 | 0.609 | 2.3 ( 1.67 - 3.16 ) | 3.27E-07 |
| rs28883356 | 4 | 89156061 | C/T | *ABCG2 - PPM1K* | 4q22.1 | 0.794 | 0.62 | 2.38 ( 1.72 - 3.32 ) | 2.35E-07 |
| rs4693941 | 4 | 89156899 | T/A | *ABCG2 - PPM1K* | 4q22.1 | 0.793 | 0.62 | 2.33 ( 1.68 - 3.23 ) | 3.69E-07 |
| rs60190393 | 4 | 89156914 | T/A | *ABCG2 - PPM1K* | 4q22.1 | 0.837 | 0.667 | 2.44 ( 1.74 - 3.44 ) | 3.08E-07 |
| rs7437679 | 4 | 89157124 | A/C | *ABCG2 - PPM1K* | 4q22.1 | 0.811 | 0.642 | 2.37 ( 1.7 - 3.3 ) | 3.47E-07 |
| rs13120254 | 4 | 89160561 | A/C | *ABCG2 - PPM1K* | 4q22.1 | 0.793 | 0.617 | 2.37 ( 1.71 - 3.29 ) | 2.49E-07 |
| rs13120819 | 4 | 89160677 | A/G | *ABCG2 - PPM1K* | 4q22.1 | 0.793 | 0.617 | 2.34 ( 1.69 - 3.24 ) | 2.90E-07 |
| rs79659488 | 4 | 89161140 | G/T | *ABCG2 - PPM1K* | 4q22.1 | 0.837 | 0.664 | 2.45 ( 1.74 - 3.45 ) | 2.48E-07 |
| rs77787810 | 4 | 89161195 | A/G | *ABCG2 - PPM1K* | 4q22.1 | 0.837 | 0.664 | 2.45 ( 1.74 - 3.45 ) | 2.48E-07 |
| rs78685931 | 4 | 89161399 | A/T | *ABCG2 - PPM1K* | 4q22.1 | 0.839 | 0.664 | 2.48 ( 1.76 - 3.5 ) | 1.85E-07 |
| rs1481018 | 4 | 89162388 | A/G | *ABCG2 - PPM1K* | 4q22.1 | 0.794 | 0.617 | 2.39 ( 1.72 - 3.32 ) | 1.93E-07 |
| rs7664939 | 4 | 89162700 | A/G | *ABCG2 - PPM1K* | 4q22.1 | 0.837 | 0.664 | 2.45 ( 1.74 - 3.45 ) | 2.48E-07 |
| rs80179435 | 4 | 89163127 | C/T | *ABCG2 - PPM1K* | 4q22.1 | 0.837 | 0.664 | 2.45 ( 1.74 - 3.45 ) | 2.48E-07 |
| rs997630 | 4 | 89163853 | G/A | *ABCG2 - PPM1K* | 4q22.1 | 0.794 | 0.62 | 2.34 ( 1.69 - 3.24 ) | 3.40E-07 |
| rs922674 | 4 | 89164722 | G/A | *ABCG2 - PPM1K* | 4q22.1 | 0.794 | 0.617 | 2.39 ( 1.72 - 3.32 ) | 1.93E-07 |
| rs6532060 | 4 | 89165494 | T/C | *ABCG2 - PPM1K* | 4q22.1 | 0.783 | 0.612 | 2.32 ( 1.67 - 3.22 ) | 4.44E-07 |
| rs6532061 | 4 | 89165613 | T/C | *ABCG2 - PPM1K* | 4q22.1 | 0.782 | 0.612 | 2.28 ( 1.65 - 3.16 ) | 7.15E-07 |
| rs7656113 | 4 | 89166536 | C/A | *ABCG2 - PPM1K* | 4q22.1 | 0.832 | 0.688 | 2.1 ( 1.5 - 2.96 ) | 1.94E-05 |
| rs219473 | 4 | 1.09E+08 | T/C | *LEF1-AS1 - RPL34-AS1* | 4q25 | 0.882 | 0.79 | 2.19 ( 1.48 - 3.24 ) | 8.15E-05 |
| rs10434071 | 4 | 1.36E+08 | G/C | *LINC02462 - LINC02485* | 4q28.3 | 0.435 | 0.328 | 1.84 ( 1.36 - 2.48 ) | 6.83E-05 |
| rs4395564 | 4 | 1.36E+08 | G/A | *LINC02462 - LINC02485* | 4q28.3 | 0.47 | 0.361 | 1.83 ( 1.36 - 2.47 ) | 6.85E-05 |
| rs7656273 | 4 | 1.36E+08 | A/G | *LINC02462 - LINC02485* | 4q28.3 | 0.478 | 0.366 | 1.8 ( 1.34 - 2.42 ) | 9.13E-05 |

| | | | | | | | | | |
|---|---|---|---|---|---|---|---|---|---|
| rs13134190 | 4 | 1.36E+08 | C/T | *LINC02462 - LINC02485* | 4q28.3 | 0.482 | 0.369 | 1.81 ( 1.35 - 2.44 ) | 8.04E-05 |
| rs6822196 | 4 | 1.36E+08 | A/T | *LINC02462 - LINC02485* | 4q28.3 | 0.482 | 0.366 | 1.83 ( 1.36 - 2.46 ) | 6.12E-05 |
| rs7693757 | 4 | 1.36E+08 | T/C | *LINC02462 - LINC02485* | 4q28.3 | 0.482 | 0.369 | 1.81 ( 1.35 - 2.44 ) | 8.04E-05 |
| rs4864240 | 4 | 1.36E+08 | T/C | *LINC02462 - LINC02485* | 4q28.3 | 0.482 | 0.369 | 1.81 ( 1.35 - 2.44 ) | 8.04E-05 |
| rs4280794 | 4 | 1.36E+08 | C/T | *LINC02462 - LINC02485* | 4q28.3 | 0.482 | 0.366 | 1.83 ( 1.36 - 2.46 ) | 6.40E-05 |
| rs4280795 | 4 | 1.36E+08 | A/T | *LINC02462 - LINC02485* | 4q28.3 | 0.482 | 0.369 | 1.81 ( 1.35 - 2.44 ) | 8.04E-05 |
| rs13129178 | 4 | 1.36E+08 | C/A | *LINC02462 - LINC02485* | 4q28.3 | 0.482 | 0.369 | 1.81 ( 1.35 - 2.44 ) | 8.04E-05 |
| rs13129208 | 4 | 1.36E+08 | G/A | *LINC02462 - LINC02485* | 4q28.3 | 0.482 | 0.369 | 1.81 ( 1.35 - 2.44 ) | 8.04E-05 |
| rs12643821 | 4 | 1.36E+08 | A/G | *LINC02462 - LINC02485* | 4q28.3 | 0.485 | 0.372 | 1.8 ( 1.34 - 2.41 ) | 9.55E-05 |
| rs55967588 | 4 | 1.36E+08 | A/G | *LINC02462 - LINC02485* | 4q28.3 | 0.482 | 0.369 | 1.81 ( 1.35 - 2.44 ) | 8.04E-05 |
| rs13126242 | 4 | 1.36E+08 | C/T | *LINC02462 - LINC02485* | 4q28.3 | 0.482 | 0.366 | 1.82 ( 1.36 - 2.45 ) | 6.98E-05 |
| rs4535381 | 4 | 1.36E+08 | T/A | *LINC02462 - LINC02485* | 4q28.3 | 0.482 | 0.369 | 1.81 ( 1.35 - 2.44 ) | 8.04E-05 |
| rs4437307 | 4 | 1.36E+08 | C/T | *LINC02462 - LINC02485* | 4q28.3 | 0.482 | 0.369 | 1.81 ( 1.35 - 2.44 ) | 8.04E-05 |
| rs11730310 | 4 | 1.36E+08 | G/A | *LINC02462 - LINC02485* | 4q28.3 | 0.481 | 0.366 | 1.82 ( 1.36 - 2.45 ) | 6.70E-05 |
| rs4597893 | 4 | 1.36E+08 | C/T | *LINC02462 - LINC02485* | 4q28.3 | 0.482 | 0.369 | 1.81 ( 1.35 - 2.44 ) | 8.04E-05 |
| rs13149380 | 4 | 1.36E+08 | C/A | *LINC02462 - LINC02485* | 4q28.3 | 0.48 | 0.366 | 1.81 ( 1.35 - 2.42 ) | 7.97E-05 |
| rs4479760 | 4 | 1.36E+08 | C/T | *LINC02462 - LINC02485* | 4q28.3 | 0.482 | 0.369 | 1.81 ( 1.35 - 2.44 ) | 8.04E-05 |
| rs12499856 | 4 | 1.36E+08 | T/C | *LINC02462 - LINC02485* | 4q28.3 | 0.494 | 0.376 | 1.79 ( 1.34 - 2.4 ) | 9.75E-05 |
| rs4328967 | 4 | 1.36E+08 | G/A | *LINC02462 - LINC02485* | 4q28.3 | 0.482 | 0.369 | 1.81 ( 1.35 - 2.44 ) | 8.04E-05 |
| rs9998499 | 4 | 1.36E+08 | G/A | *LINC02462 - LINC02485* | 4q28.3 | 0.489 | 0.369 | 1.85 ( 1.38 - 2.48 ) | 4.05E-05 |
| rs7656631 | 4 | 1.36E+08 | C/T | *LINC02462 - LINC02485* | 4q28.3 | 0.483 | 0.369 | 1.82 ( 1.36 - 2.45 ) | 6.90E-05 |
| rs13142779 | 4 | 1.36E+08 | T/G | *LINC02462 - LINC02485* | 4q28.3 | 0.48 | 0.366 | 1.81 ( 1.35 - 2.43 ) | 7.82E-05 |
| rs6535050 | 4 | 1.36E+08 | A/C | *LINC02462 - LINC02485* | 4q28.3 | 0.483 | 0.369 | 1.83 ( 1.36 - 2.46 ) | 6.25E-05 |
| rs6535051 | 4 | 1.36E+08 | G/A | *LINC02462 - LINC02485* | 4q28.3 | 0.482 | 0.369 | 1.81 ( 1.35 - 2.44 ) | 8.04E-05 |
| rs7656209 | 4 | 1.36E+08 | A/G | *LINC02462 - LINC02485* | 4q28.3 | 0.482 | 0.369 | 1.81 ( 1.35 - 2.44 ) | 8.04E-05 |
| rs6838419 | 4 | 1.36E+08 | T/C | *LINC02462 - LINC02485* | 4q28.3 | 0.482 | 0.369 | 1.81 ( 1.35 - 2.44 ) | 8.04E-05 |

| | | | | | | | | | |
|---|---|---|---|---|---|---|---|---|---|
| rs7656595 | 4 | 1.36E+08 | A/G | *LINC02462 - LINC02485* | 4q28.3 | 0.482 | 0.366 | 1.84 ( 1.36 - 2.47 ) | 5.81E-05 |
| rs28801464 | 4 | 1.36E+08 | A/G | *LINC02462 - LINC02485* | 4q28.3 | 0.482 | 0.369 | 1.81 ( 1.35 - 2.44 ) | 8.04E-05 |
| rs28826933 | 4 | 1.36E+08 | T/A | *LINC02462 - LINC02485* | 4q28.3 | 0.482 | 0.369 | 1.81 ( 1.35 - 2.44 ) | 8.04E-05 |
| rs12651324 | 4 | 1.63E+08 | A/G | *FSTL5 - MIR4454* | 4q32.2 | 0.754 | 0.623 | 1.86 ( 1.36 - 2.53 ) | 9.16E-05 |
| rs35213808 | 4 | 1.63E+08 | T/A | *FSTL5 - MIR4454* | 4q32.2 | 0.756 | 0.623 | 1.89 ( 1.38 - 2.58 ) | 6.59E-05 |
| rs12642324 | 4 | 1.63E+08 | T/C | *FSTL5 - MIR4454* | 4q32.2 | 0.754 | 0.623 | 1.87 ( 1.37 - 2.55 ) | 8.80E-05 |
| rs17539368 | 4 | 1.63E+08 | T/G | *FSTL5 - MIR4454* | 4q32.2 | 0.754 | 0.623 | 1.87 ( 1.37 - 2.55 ) | 8.80E-05 |
| rs11941389 | 4 | 1.63E+08 | T/C | *FSTL5 - MIR4454* | 4q32.2 | 0.754 | 0.623 | 1.87 ( 1.37 - 2.55 ) | 8.80E-05 |
| rs1001467 | 4 | 1.63E+08 | A/G | *FSTL5 - MIR4454* | 4q32.2 | 0.754 | 0.623 | 1.87 ( 1.37 - 2.55 ) | 8.80E-05 |
| rs36015093 | 4 | 1.63E+08 | C/T | *FSTL5 - MIR4454* | 4q32.2 | 0.754 | 0.623 | 1.87 ( 1.37 - 2.56 ) | 8.03E-05 |
| rs13104331 | 4 | 1.63E+08 | G/A | *FSTL5 - MIR4454* | 4q32.2 | 0.756 | 0.623 | 1.89 ( 1.38 - 2.58 ) | 6.59E-05 |
| rs28456058 | 4 | 1.87E+08 | T/C | *F11-AS1* | 4q35.2 | 0.304 | 0.194 | 2.03 ( 1.43 - 2.87 ) | 6.90E-05 |
| rs143147592 | 5 | 10833027 | C/G | *DAP - CTNND2* | 5p15.2 | 0.969 | 0.893 | 3.57 ( 1.9 - 6.68 ) | 7.34E-05 |
| rs4866089 | 5 | 18287613 | G/A | *LINC02223 - CDH18* | 5p15.1 | 0.893 | 0.803 | 2.2 ( 1.48 - 3.26 ) | 8.79E-05 |
| rs4866327 | 5 | 18333591 | A/G | *LINC02223 - CDH18* | 5p15.1 | 0.854 | 0.76 | 2.12 ( 1.47 - 3.06 ) | 5.74E-05 |
| rs32494 | 5 | 55643774 | C/T | *ANKRD55 - LINC01948* | 5q11.2 | 0.578 | 0.41 | 1.84 ( 1.39 - 2.43 ) | 1.81E-05 |
| rs6895497 | 5 | 85049974 | T/C | *EDIL3 - NBPF22P* | 5q14.3 | 0.572 | 0.434 | 1.76 ( 1.33 - 2.34 ) | 8.45E-05 |
| rs62363123 | 5 | 85051118 | G/T | *EDIL3 - NBPF22P* | 5q14.3 | 0.574 | 0.437 | 1.77 ( 1.33 - 2.35 ) | 8.38E-05 |
| rs7716856 | 5 | 85089289 | A/G | *EDIL3 - NBPF22P* | 5q14.3 | 0.57 | 0.434 | 1.8 ( 1.35 - 2.4 ) | 6.56E-05 |
| rs529598858 | 5 | 1.21E+08 | A/* | . | . | 0.948 | 0.879 | 3.02 ( 1.75 - 5.2 ) | 6.90E-05 |
| rs1421878 | 5 | 1.22E+08 | G/A | *LOC100505841 - SNCAIP* | 5q23.2 | 0.913 | 0.839 | 2.47 ( 1.57 - 3.9 ) | 9.80E-05 |
| rs1421879 | 5 | 1.22E+08 | G/T | *LOC100505841 - SNCAIP* | 5q23.2 | 0.913 | 0.839 | 2.47 ( 1.57 - 3.9 ) | 9.80E-05 |
| rs6864421 | 5 | 1.22E+08 | C/T | *LOC100505841 - SNCAIP* | 5q23.2 | 0.913 | 0.839 | 2.47 ( 1.57 - 3.9 ) | 9.80E-05 |
| rs6885626 | 5 | 1.22E+08 | T/C | *LOC100505841 - SNCAIP* | 5q23.2 | 0.913 | 0.839 | 2.47 ( 1.57 - 3.9 ) | 9.80E-05 |
| rs1363163 | 5 | 1.22E+08 | T/C | *LOC100505841 - SNCAIP* | 5q23.2 | 0.913 | 0.839 | 2.47 ( 1.57 - 3.9 ) | 9.80E-05 |
| rs4895346 | 5 | 1.22E+08 | A/G | *LOC100505841 - SNCAIP* | 5q23.2 | 0.906 | 0.822 | 2.47 ( 1.59 - 3.84 ) | 5.27E-05 |

| | | | | | | | | | |
|---|---|---|---|---|---|---|---|---|---|
| rs6859604 | 5 | 1.22E+08 | C/G | *LOC100505841 - SNCAIP* | 5q23.2 | 0.906 | 0.822 | 2.47 ( 1.59 - 3.84 ) | 5.27E-05 |
| rs6881951 | 5 | 1.22E+08 | T/A | *LOC100505841 - SNCAIP* | 5q23.2 | 0.906 | 0.822 | 2.47 ( 1.59 - 3.84 ) | 5.27E-05 |
| rs6881968 | 5 | 1.22E+08 | T/A | *LOC100505841 - SNCAIP* | 5q23.2 | 0.906 | 0.822 | 2.47 ( 1.59 - 3.84 ) | 5.27E-05 |
| rs2407110 | 5 | 1.22E+08 | G/A | *LOC100505841 - SNCAIP* | 5q23.2 | 0.906 | 0.822 | 2.47 ( 1.59 - 3.84 ) | 5.27E-05 |
| rs6861145 | 5 | 1.22E+08 | G/A | *LOC100505841 - SNCAIP* | 5q23.2 | 0.907 | 0.822 | 2.55 ( 1.64 - 3.97 ) | 3.20E-05 |
| rs6865529 | 5 | 1.22E+08 | G/C | *LOC100505841 - SNCAIP* | 5q23.2 | 0.906 | 0.825 | 2.42 ( 1.56 - 3.76 ) | 8.04E-05 |
| rs11958217 | 5 | 1.22E+08 | A/G | *LOC100505841 - SNCAIP* | 5q23.2 | 0.907 | 0.822 | 2.54 ( 1.63 - 3.95 ) | 3.51E-05 |
| rs12656426 | 5 | 1.22E+08 | T/C | *LOC100505841 - SNCAIP* | 5q23.2 | 0.906 | 0.822 | 2.47 ( 1.59 - 3.84 ) | 5.27E-05 |
| rs6871091 | 5 | 1.22E+08 | G/C | *LOC100505841 - SNCAIP* | 5q23.2 | 0.906 | 0.822 | 2.47 ( 1.59 - 3.84 ) | 5.27E-05 |
| rs2059049 | 5 | 1.22E+08 | T/C | *LOC100505841 - SNCAIP* | 5q23.2 | 0.906 | 0.822 | 2.47 ( 1.59 - 3.84 ) | 5.27E-05 |
| rs11241634 | 5 | 1.22E+08 | A/C | *LOC100505841 - SNCAIP* | 5q23.2 | 0.906 | 0.822 | 2.47 ( 1.59 - 3.84 ) | 5.27E-05 |
| rs10052044 | 5 | 1.22E+08 | G/C | *LOC100505841 - SNCAIP* | 5q23.2 | 0.906 | 0.822 | 2.47 ( 1.59 - 3.84 ) | 5.27E-05 |
| rs2059048 | 5 | 1.22E+08 | T/C | *LOC100505841 - SNCAIP* | 5q23.2 | 0.906 | 0.822 | 2.47 ( 1.59 - 3.84 ) | 5.27E-05 |
| rs918384 | 5 | 1.22E+08 | G/A | *LOC100505841 - SNCAIP* | 5q23.2 | 0.906 | 0.822 | 2.47 ( 1.59 - 3.84 ) | 5.27E-05 |
| rs2193963 | 5 | 1.22E+08 | A/G | *LOC100505841 - SNCAIP* | 5q23.2 | 0.906 | 0.822 | 2.47 ( 1.59 - 3.84 ) | 5.27E-05 |
| rs9327244 | 5 | 1.22E+08 | C/G | *LOC100505841 - SNCAIP* | 5q23.2 | 0.906 | 0.822 | 2.47 ( 1.59 - 3.84 ) | 5.27E-05 |
| rs10058354 | 5 | 1.22E+08 | C/T | *LOC100505841 - SNCAIP* | 5q23.2 | 0.906 | 0.822 | 2.47 ( 1.59 - 3.84 ) | 5.27E-05 |
| rs731312 | 5 | 1.22E+08 | T/C | *LOC100505841 - SNCAIP* | 5q23.2 | 0.906 | 0.822 | 2.47 ( 1.59 - 3.84 ) | 5.27E-05 |
| rs62381663 | 5 | 1.22E+08 | T/C | *LOC100505841 - SNCAIP* | 5q23.2 | 0.906 | 0.82 | 2.48 ( 1.6 - 3.83 ) | 4.40E-05 |
| rs11241636 | 5 | 1.22E+08 | T/C | *LOC100505841 - SNCAIP* | 5q23.2 | 0.907 | 0.825 | 2.47 ( 1.59 - 3.84 ) | 6.18E-05 |
| rs13157506 | 5 | 1.22E+08 | G/A | *LOC100505841 - SNCAIP* | 5q23.2 | 0.911 | 0.828 | 2.53 ( 1.62 - 3.96 ) | 4.93E-05 |
| rs12659215 | 5 | 1.22E+08 | A/G | *LOC100505841 - SNCAIP* | 5q23.2 | 0.917 | 0.818 | 2.58 ( 1.66 - 4.02 ) | 2.64E-05 |
| rs61692601 | 6 | 19227233 | C/A | *LOC101928519 - LOC105374960* | 6p22.3 | 0.906 | 0.817 | 2.29 ( 1.52 - 3.44 ) | 7.00E-05 |
| rs4895485 | 6 | 1.38E+08 | T/C | *IFNGR1 - OLIG3* | 6q23.3 | 0.309 | 0.205 | 2 ( 1.43 - 2.8 ) | 5.92E-05 |
| rs12666668 | 7 | 18068820 | A/T | *PRPS1L1 - HDAC9* | 7p21.1 | 0.261 | 0.148 | 2.17 ( 1.51 - 3.12 ) | 2.77E-05 |
| rs12668788 | 7 | 18068931 | G/A | *PRPS1L1 - HDAC9* | 7p21.1 | 0.263 | 0.15 | 2.11 ( 1.47 - 3.01 ) | 4.39E-05 |

| | | | | | | | | | |
|---|---|---|---|---|---|---|---|---|---|
| rs59972673 | 7 | 18071528 | G/A | *PRPS1L1 - HDAC9* | 7p21.1 | 0.248 | 0.142 | 2.17 ( 1.5 - 3.15 ) | 4.51E-05 |
| rs79605413 | 7 | 18078160 | A/T | *PRPS1L1 - HDAC9* | 7p21.1 | 0.244 | 0.134 | 2.26 ( 1.55 - 3.3 ) | 2.32E-05 |
| rs78253013 | 7 | 18078591 | T/C | *PRPS1L1 - HDAC9* | 7p21.1 | 0.244 | 0.134 | 2.26 ( 1.55 - 3.3 ) | 2.32E-05 |
| rs6977061 | 7 | 18079091 | G/A | *PRPS1L1 - HDAC9* | 7p21.1 | 0.243 | 0.137 | 2.19 ( 1.5 - 3.18 ) | 4.47E-05 |
| rs76938223 | 7 | 18080341 | A/G | *PRPS1L1 - HDAC9* | 7p21.1 | 0.246 | 0.139 | 2.16 ( 1.49 - 3.13 ) | 4.84E-05 |
| rs78217653 | 7 | 18080413 | A/G | *PRPS1L1 - HDAC9* | 7p21.1 | 0.246 | 0.137 | 2.21 ( 1.52 - 3.21 ) | 3.29E-05 |
| rs77379626 | 7 | 18080883 | C/T | *PRPS1L1 - HDAC9* | 7p21.1 | 0.248 | 0.139 | 2.19 ( 1.51 - 3.18 ) | 3.70E-05 |
| rs76458460 | 7 | 18081455 | T/C | *PRPS1L1 - HDAC9* | 7p21.1 | 0.244 | 0.139 | 2.14 ( 1.48 - 3.1 ) | 6.02E-05 |
| rs111881036 | 7 | 18081745 | C/T | *PRPS1L1 - HDAC9* | 7p21.1 | 0.248 | 0.139 | 2.19 ( 1.51 - 3.18 ) | 3.70E-05 |
| rs79631409 | 7 | 18082011 | G/A | *PRPS1L1 - HDAC9* | 7p21.1 | 0.246 | 0.139 | 2.14 ( 1.48 - 3.1 ) | 5.63E-05 |
| rs74443566 | 7 | 18082463 | T/C | *PRPS1L1 - HDAC9* | 7p21.1 | 0.246 | 0.137 | 2.19 ( 1.51 - 3.19 ) | 3.67E-05 |
| rs76755239 | 7 | 18083492 | A/C | *PRPS1L1 - HDAC9* | 7p21.1 | 0.248 | 0.139 | 2.19 ( 1.51 - 3.18 ) | 3.70E-05 |
| rs79978844 | 7 | 18083645 | A/G | *PRPS1L1 - HDAC9* | 7p21.1 | 0.248 | 0.139 | 2.19 ( 1.51 - 3.18 ) | 3.70E-05 |
| rs4326292 | 7 | 18083820 | C/G | *PRPS1L1 - HDAC9* | 7p21.1 | 0.248 | 0.139 | 2.19 ( 1.51 - 3.18 ) | 3.70E-05 |
| rs10499512 | 7 | 18084843 | G/A | *PRPS1L1 - HDAC9* | 7p21.1 | 0.25 | 0.137 | 2.25 ( 1.55 - 3.26 ) | 2.18E-05 |
| rs79654337 | 7 | 18084960 | A/G | *PRPS1L1 - HDAC9* | 7p21.1 | 0.248 | 0.139 | 2.19 ( 1.51 - 3.18 ) | 3.70E-05 |
| rs17138549 | 7 | 18085049 | G/T | *PRPS1L1 - HDAC9* | 7p21.1 | 0.248 | 0.137 | 2.21 ( 1.52 - 3.21 ) | 3.04E-05 |
| rs74892486 | 7 | 18085428 | G/A | *PRPS1L1 - HDAC9* | 7p21.1 | 0.244 | 0.139 | 2.13 ( 1.47 - 3.08 ) | 6.61E-05 |
| rs17138551 | 7 | 18085676 | A/G | *PRPS1L1 - HDAC9* | 7p21.1 | 0.248 | 0.139 | 2.19 ( 1.51 - 3.18 ) | 3.70E-05 |
| rs17138557 | 7 | 18086614 | G/A | *PRPS1L1 - HDAC9* | 7p21.1 | 0.246 | 0.137 | 2.23 ( 1.53 - 3.25 ) | 2.98E-05 |
| rs76494461 | 7 | 18088568 | A/G | *PRPS1L1 - HDAC9* | 7p21.1 | 0.27 | 0.158 | 2.13 ( 1.49 - 3.04 ) | 3.55E-05 |
| rs79702394 | 7 | 18088572 | C/T | *PRPS1L1 - HDAC9* | 7p21.1 | 0.272 | 0.158 | 2.14 ( 1.5 - 3.07 ) | 2.94E-05 |
| rs117858288 | 7 | 18092093 | T/C | *PRPS1L1 - HDAC9* | 7p21.1 | 0.268 | 0.158 | 2.09 ( 1.46 - 2.98 ) | 5.00E-05 |
| rs61388592 | 7 | 88350769 | C/T | *STEAP4 - ZNF804B* | 7q21.13 | 0.382 | 0.268 | 1.88 ( 1.37 - 2.57 ) | 9.68E-05 |
| rs56963120 | 7 | 88351391 | T/A | *STEAP4 - ZNF804B* | 7q21.13 | 0.382 | 0.268 | 1.88 ( 1.37 - 2.57 ) | 9.68E-05 |
| rs17163911 | 7 | 88352832 | C/T | *STEAP4 - ZNF804B* | 7q21.13 | 0.387 | 0.268 | 1.9 ( 1.38 - 2.61 ) | 7.33E-05 |

| | | | | | | | | | |
|---|---|---|---|---|---|---|---|---|---|
| rs10952926 | 7 | 88356414 | C/T | *STEAP4 - ZNF804B* | 7q21.13 | 0.383 | 0.265 | 1.9 ( 1.39 - 2.62 ) | 7.20E-05 |
| rs66590293 | 7 | 88365083 | A/T | *STEAP4 - ZNF804B* | 7q21.13 | 0.498 | 0.366 | 1.86 ( 1.38 - 2.52 ) | 4.61E-05 |
| rs9641035 | 7 | 88421333 | T/C | *ZNF804B* | 7q21.13 | 0.378 | 0.251 | 2.07 ( 1.49 - 2.86 ) | 1.19E-05 |
| rs11769319 | 7 | 95980767 | A/G | *SLC25A13 - SEM1* | 7q21.3 | 0.63 | 0.511 | 1.84 ( 1.36 - 2.51 ) | 9.40E-05 |
| rs79656125 | 7 | 1.08E+08 | A/G | *DNAJB9 - C7orf66* | 7q31.1 | 0.965 | 0.902 | 3.62 ( 1.92 - 6.85 ) | 7.33E-05 |
| rs77631916 | 7 | 1.08E+08 | G/A | *DNAJB9 - C7orf66* | 7q31.1 | 0.967 | 0.902 | 3.96 ( 2.07 - 7.58 ) | 3.34E-05 |
| rs67337609 | 7 | 1.31E+08 | T/C | *PODXL - LOC101928782* | 7q32.3 | 0.957 | 0.852 | 3.65 ( 2.14 - 6.2 ) | 1.78E-06 |
| rs73159568 | 7 | 1.31E+08 | G/T | *PODXL - LOC101928782* | 7q32.3 | 0.957 | 0.855 | 3.65 ( 2.14 - 6.25 ) | 2.23E-06 |
| rs78506532 | 8 | 5476076 | T/G | *CSMD1 - LOC100287015* | 8p23.2 | 0.831 | 0.71 | 1.98 ( 1.42 - 2.77 ) | 6.50E-05 |
| rs1477926733 | 8 | 51993276 | */T | . | . | 0.891 | 0.794 | 2.86 ( 1.83 - 4.46 ) | 3.96E-06 |
| rs66620947 | 9 | 36559533 | G/T | *RNF38 - MELK* | 9p13.2 | 0.959 | 0.888 | 3.23 ( 1.83 - 5.7 ) | 5.30E-05 |
| rs449451 | 9 | 36565685 | A/G | *RNF38 - MELK* | 9p13.2 | 0.959 | 0.891 | 3.27 ( 1.84 - 5.81 ) | 5.59E-05 |
| rs329355 | 9 | 36571473 | A/C | *RNF38 - MELK* | 9p13.2 | 0.961 | 0.896 | 3.25 ( 1.81 - 5.86 ) | 8.44E-05 |
| rs10511963 | 9 | 71421359 | C/T | *PIP5K1B* | 9q21.11 | 0.759 | 0.626 | 1.88 ( 1.39 - 2.54 ) | 4.16E-05 |
| rs10868794 | 9 | 91178471 | A/G | *NXNL2* | 9q22.1 | 0.354 | 0.219 | 1.9 ( 1.38 - 2.62 ) | 7.29E-05 |
| rs647439 | 10 | 6228413 | T/C | *PFKFB3* | 10p15.1 | 0.45 | 0.328 | 1.86 ( 1.36 - 2.54 ) | 9.80E-05 |
| rs646564 | 10 | 6228598 | C/A | *PFKFB3* | 10p15.1 | 0.454 | 0.328 | 1.9 ( 1.39 - 2.59 ) | 5.76E-05 |
| rs645680 | 10 | 6228777 | A/G | *PFKFB3* | 10p15.1 | 0.454 | 0.331 | 1.88 ( 1.37 - 2.56 ) | 8.37E-05 |
| rs645611 | 10 | 6228824 | T/A | *PFKFB3* | 10p15.1 | 0.454 | 0.331 | 1.88 ( 1.37 - 2.56 ) | 8.37E-05 |
| rs653659 | 10 | 6228878 | T/C | *PFKFB3* | 10p15.1 | 0.454 | 0.331 | 1.88 ( 1.37 - 2.56 ) | 8.37E-05 |
| rs634507 | 10 | 6228979 | C/A | *PFKFB3* | 10p15.1 | 0.454 | 0.331 | 1.88 ( 1.37 - 2.56 ) | 8.37E-05 |
| rs634506 | 10 | 6228980 | A/G | *PFKFB3* | 10p15.1 | 0.454 | 0.331 | 1.88 ( 1.37 - 2.56 ) | 8.37E-05 |
| rs652399 | 10 | 6229127 | T/C | *PFKFB3* | 10p15.1 | 0.45 | 0.325 | 1.86 ( 1.36 - 2.54 ) | 8.99E-05 |
| rs10828842 | 10 | 25954464 | A/G | *LINC00836* | 10p12.1 | 0.911 | 0.844 | 2.54 ( 1.61 - 4.02 ) | 6.86E-05 |
| rs482468 | 10 | 27671644 | G/C | *ARMC4P1 - PTCHD3* | 10p12.1 | 0.444 | 0.322 | 1.8 ( 1.35 - 2.42 ) | 7.49E-05 |
| rs11015730 | 10 | 27671847 | T/C | *ARMC4P1 - PTCHD3* | 10p12.1 | 0.418 | 0.3 | 1.8 ( 1.34 - 2.41 ) | 8.37E-05 |

| | | | | | | | | | |
|---|---|---|---|---|---|---|---|---|---|
| rs10829253 | 10 | 27675276 | T/C | *ARMC4P1 - PTCHD3* | 10p12.1 | 0.42 | 0.3 | 1.81 ( 1.35 - 2.43 ) | 6.77E-05 |
| rs61591337 | 10 | 27678461 | T/C | *ARMC4P1 - PTCHD3* | 10p12.1 | 0.418 | 0.298 | 1.83 ( 1.36 - 2.46 ) | 5.57E-05 |
| rs506659 | 10 | 27678534 | T/C | *ARMC4P1 - PTCHD3* | 10p12.1 | 0.444 | 0.322 | 1.8 ( 1.35 - 2.42 ) | 7.49E-05 |
| rs7100677 | 10 | 27681526 | A/G | *ARMC4P1 - PTCHD3* | 10p12.1 | 0.428 | 0.309 | 1.81 ( 1.35 - 2.43 ) | 7.07E-05 |
| rs11015740 | 10 | 27690072 | G/T | *PTCHD3* | 10p12.1 | 0.428 | 0.303 | 1.85 ( 1.38 - 2.48 ) | 3.67E-05 |
| rs111812553 | 10 | 76624101 | C/T | *KAT6B* | 10q22.2 | 0.935 | 0.872 | 2.82 ( 1.69 - 4.7 ) | 7.41E-05 |
| rs10509352 | 10 | 76637516 | A/G | *KAT6B* | 10q22.2 | 0.935 | 0.872 | 2.81 ( 1.69 - 4.68 ) | 7.33E-05 |
| rs75595613 | 10 | 76679000 | A/T | *LOC101929165* | 10q22.2 | 0.935 | 0.869 | 2.78 ( 1.68 - 4.61 ) | 6.76E-05 |
| rs7896390 | 10 | 1.14E+08 | C/T | *ADRA2A - GPAM* | 10q25.2 | 0.224 | 0.109 | 2.28 ( 1.52 - 3.41 ) | 5.84E-05 |
| rs11195714 | 10 | 1.14E+08 | G/A | *ADRA2A - GPAM* | 10q25.2 | 0.224 | 0.109 | 2.28 ( 1.52 - 3.41 ) | 5.84E-05 |
| rs7085622 | 10 | 1.14E+08 | A/G | *ADRA2A - GPAM* | 10q25.2 | 0.222 | 0.109 | 2.25 ( 1.51 - 3.36 ) | 7.57E-05 |
| rs1264803 | 10 | 1.17E+08 | A/T | *ATRNL1* | 10q25.3 | 0.768 | 0.645 | 1.89 ( 1.37 - 2.6 ) | 9.46E-05 |
| rs1264798 | 10 | 1.17E+08 | A/G | *ATRNL1* | 10q25.3 | 0.77 | 0.639 | 2 ( 1.44 - 2.76 ) | 2.90E-05 |
| rs1615076 | 10 | 1.17E+08 | G/A | *ATRNL1* | 10q25.3 | 0.783 | 0.658 | 1.92 ( 1.39 - 2.66 ) | 7.86E-05 |
| rs2165988 | 10 | 1.17E+08 | A/T | *ATRNL1* | 10q25.3 | 0.782 | 0.653 | 1.96 ( 1.41 - 2.71 ) | 5.12E-05 |
| rs2201363 | 10 | 1.17E+08 | T/C | *ATRNL1* | 10q25.3 | 0.75 | 0.628 | 1.87 ( 1.37 - 2.57 ) | 9.80E-05 |
| rs10787567 | 10 | 1.17E+08 | C/T | *ATRNL1* | 10q25.3 | 0.776 | 0.645 | 2.03 ( 1.46 - 2.8 ) | 1.98E-05 |
| rs650060 | 10 | 1.17E+08 | G/C | *ATRNL1* | 10q25.3 | 0.772 | 0.642 | 1.99 ( 1.44 - 2.74 ) | 2.50E-05 |
| rs615905 | 10 | 1.17E+08 | A/G | *ATRNL1* | 10q25.3 | 0.778 | 0.648 | 2.01 ( 1.45 - 2.78 ) | 2.33E-05 |
| rs675816 | 10 | 1.17E+08 | A/G | *ATRNL1* | 10q25.3 | 0.752 | 0.631 | 1.88 ( 1.37 - 2.58 ) | 9.27E-05 |
| rs2485968 | 10 | 1.17E+08 | A/G | *ATRNL1* | 10q25.3 | 0.823 | 0.664 | 2.31 ( 1.66 - 3.2 ) | 5.72E-07 |
| rs2485971 | 10 | 1.17E+08 | A/G | *ATRNL1* | 10q25.3 | 0.776 | 0.642 | 2.06 ( 1.49 - 2.85 ) | 1.31E-05 |
| rs1565833 | 10 | 1.17E+08 | A/G | *ATRNL1* | 10q25.3 | 0.778 | 0.642 | 2.08 ( 1.5 - 2.88 ) | 1.01E-05 |
| rs683117 | 10 | 1.17E+08 | G/A | *ATRNL1* | 10q25.3 | 0.753 | 0.631 | 1.89 ( 1.38 - 2.59 ) | 8.29E-05 |
| rs2485966 | 10 | 1.17E+08 | T/C | *ATRNL1* | 10q25.3 | 0.774 | 0.642 | 2.02 ( 1.47 - 2.79 ) | 1.85E-05 |
| rs594160 | 10 | 1.17E+08 | T/C | *ATRNL1* | 10q25.3 | 0.776 | 0.642 | 2.06 ( 1.49 - 2.85 ) | 1.31E-05 |

| | | | | | | | | | |
|---|---|---|---|---|---|---|---|---|---|
| rs621652 | 10 | 1.17E+08 | A/T | *ATRNL1* | 10q25.3 | 0.787 | 0.653 | 2.08 ( 1.5 - 2.9 ) | 1.39E-05 |
| rs139875711 | 10 | 1.21E+08 | */C | . | . | 0.093 | 0.017 | 6.29 ( 2.58 - 15.37 ) | 5.46E-05 |
| rs117537199 | 11 | 1.1E+08 | A/T | *C11orf87 - ZC3H12C* | 11q22.3 | 0.132 | 0.049 | 3.26 ( 1.85 - 5.76 ) | 4.65E-05 |
| rs2852132 | 11 | 1.1E+08 | C/T | *C11orf87 - ZC3H12C* | 11q22.3 | 0.132 | 0.049 | 3.26 ( 1.85 - 5.76 ) | 4.65E-05 |
| rs667042 | 11 | 1.1E+08 | A/C | *C11orf87 - ZC3H12C* | 11q22.3 | 0.13 | 0.049 | 3.17 ( 1.79 - 5.61 ) | 7.20E-05 |
| rs519898 | 11 | 1.1E+08 | T/C | *C11orf87 - ZC3H12C* | 11q22.3 | 0.13 | 0.049 | 3.17 ( 1.79 - 5.61 ) | 7.20E-05 |
| rs1944915 | 11 | 1.1E+08 | A/T | *C11orf87 - ZC3H12C* | 11q22.3 | 0.13 | 0.049 | 3.17 ( 1.79 - 5.61 ) | 7.20E-05 |
| rs1836098 | 11 | 1.1E+08 | C/G | *C11orf87 - ZC3H12C* | 11q22.3 | 0.13 | 0.049 | 3.17 ( 1.79 - 5.61 ) | 7.20E-05 |
| rs1789818 | 11 | 1.1E+08 | G/T | *C11orf87 - ZC3H12C* | 11q22.3 | 0.128 | 0.046 | 3.34 ( 1.87 - 5.98 ) | 4.70E-05 |
| rs689152 | 11 | 1.1E+08 | T/C | *C11orf87 - ZC3H12C* | 11q22.3 | 0.126 | 0.046 | 3.25 ( 1.82 - 5.82 ) | 7.28E-05 |
| rs559717 | 11 | 1.1E+08 | G/C | *C11orf87 - ZC3H12C* | 11q22.3 | 0.132 | 0.049 | 3.26 ( 1.85 - 5.76 ) | 4.65E-05 |
| rs659793 | 11 | 1.1E+08 | G/A | *C11orf87 - ZC3H12C* | 11q22.3 | 0.132 | 0.049 | 3.26 ( 1.85 - 5.76 ) | 4.65E-05 |
| rs644425 | 11 | 1.1E+08 | C/G | *C11orf87 - ZC3H12C* | 11q22.3 | 0.132 | 0.049 | 3.26 ( 1.85 - 5.76 ) | 4.65E-05 |
| rs1789815 | 11 | 1.1E+08 | T/C | *C11orf87 - ZC3H12C* | 11q22.3 | 0.132 | 0.049 | 3.26 ( 1.85 - 5.76 ) | 4.65E-05 |
| rs546359 | 11 | 1.1E+08 | G/A | *C11orf87 - ZC3H12C* | 11q22.3 | 0.13 | 0.049 | 3.17 ( 1.79 - 5.61 ) | 7.20E-05 |
| rs544555 | 11 | 1.1E+08 | G/C | *C11orf87 - ZC3H12C* | 11q22.3 | 0.13 | 0.049 | 3.17 ( 1.79 - 5.61 ) | 7.20E-05 |
| rs2851174 | 11 | 1.1E+08 | T/C | *C11orf87 - ZC3H12C* | 11q22.3 | 0.13 | 0.049 | 3.17 ( 1.79 - 5.61 ) | 7.20E-05 |
| rs2852136 | 11 | 1.1E+08 | G/A | *C11orf87 - ZC3H12C* | 11q22.3 | 0.13 | 0.049 | 3.17 ( 1.79 - 5.61 ) | 7.20E-05 |
| rs2852134 | 11 | 1.1E+08 | G/A | *C11orf87 - ZC3H12C* | 11q22.3 | 0.13 | 0.049 | 3.17 ( 1.79 - 5.61 ) | 7.20E-05 |
| rs672958 | 11 | 1.1E+08 | G/T | *C11orf87 - ZC3H12C* | 11q22.3 | 0.13 | 0.049 | 3.17 ( 1.79 - 5.61 ) | 7.20E-05 |
| rs694826 | 11 | 1.1E+08 | T/C | *C11orf87 - ZC3H12C* | 11q22.3 | 0.13 | 0.049 | 3.17 ( 1.79 - 5.61 ) | 7.20E-05 |
| rs1789821 | 11 | 1.1E+08 | A/G | *C11orf87 - ZC3H12C* | 11q22.3 | 0.133 | 0.049 | 3.22 ( 1.83 - 5.68 ) | 5.14E-05 |
| rs1893872 | 11 | 1.1E+08 | C/G | *C11orf87 - ZC3H12C* | 11q22.3 | 0.13 | 0.049 | 3.19 ( 1.81 - 5.65 ) | 6.55E-05 |
| rs533037 | 11 | 1.1E+08 | C/T | *C11orf87 - ZC3H12C* | 11q22.3 | 0.13 | 0.049 | 3.17 ( 1.79 - 5.61 ) | 7.20E-05 |
| rs616219 | 11 | 1.1E+08 | T/G | *C11orf87 - ZC3H12C* | 11q22.3 | 0.13 | 0.049 | 3.17 ( 1.79 - 5.61 ) | 7.20E-05 |
| rs4633423 | 11 | 1.1E+08 | C/A | *C11orf87 - ZC3H12C* | 11q22.3 | 0.13 | 0.049 | 3.17 ( 1.79 - 5.61 ) | 7.20E-05 |

| | | | | | | | | | |
|---|---|---|---|---|---|---|---|---|---|
| rs637478 | 11 | 1.1E+08 | G/T | *C11orf87 - ZC3H12C* | 11q22.3 | 0.13 | 0.049 | 3.17 ( 1.79 - 5.61 ) | 7.20E-05 |
| rs471773 | 11 | 1.1E+08 | C/T | *C11orf87 - ZC3H12C* | 11q22.3 | 0.13 | 0.049 | 3.17 ( 1.79 - 5.61 ) | 7.20E-05 |
| rs529544 | 11 | 1.1E+08 | A/G | *C11orf87 - ZC3H12C* | 11q22.3 | 0.13 | 0.049 | 3.17 ( 1.79 - 5.61 ) | 7.20E-05 |
| rs560631 | 11 | 1.1E+08 | C/A | *C11orf87 - ZC3H12C* | 11q22.3 | 0.13 | 0.049 | 3.17 ( 1.79 - 5.61 ) | 7.20E-05 |
| rs564374 | 11 | 1.1E+08 | G/T | *C11orf87 - ZC3H12C* | 11q22.3 | 0.13 | 0.049 | 3.17 ( 1.79 - 5.61 ) | 7.20E-05 |
| rs485166 | 11 | 1.1E+08 | T/C | *C11orf87 - ZC3H12C* | 11q22.3 | 0.13 | 0.049 | 3.17 ( 1.79 - 5.61 ) | 7.20E-05 |
| rs7937607 | 11 | 1.1E+08 | A/G | *C11orf87 - ZC3H12C* | 11q22.3 | 0.148 | 0.06 | 3.07 ( 1.81 - 5.2 ) | 3.26E-05 |
| rs538380 | 11 | 1.1E+08 | T/G | *C11orf87 - ZC3H12C* | 11q22.3 | 0.13 | 0.049 | 3.17 ( 1.79 - 5.61 ) | 7.20E-05 |
| rs594536 | 11 | 1.1E+08 | C/T | *C11orf87 - ZC3H12C* | 11q22.3 | 0.128 | 0.046 | 3.32 ( 1.86 - 5.94 ) | 5.18E-05 |
| rs552700 | 11 | 1.1E+08 | G/A | *C11orf87 - ZC3H12C* | 11q22.3 | 0.128 | 0.046 | 3.32 ( 1.86 - 5.94 ) | 5.18E-05 |
| rs554442 | 11 | 1.1E+08 | G/A | *C11orf87 - ZC3H12C* | 11q22.3 | 0.128 | 0.046 | 3.32 ( 1.86 - 5.94 ) | 5.18E-05 |
| rs685445 | 11 | 1.1E+08 | T/G | *C11orf87 - ZC3H12C* | 11q22.3 | 0.128 | 0.046 | 3.32 ( 1.86 - 5.94 ) | 5.18E-05 |
| rs10891022 | 11 | 1.1E+08 | A/T | *C11orf87 - ZC3H12C* | 11q22.3 | 0.128 | 0.046 | 3.32 ( 1.86 - 5.94 ) | 5.18E-05 |
| rs659402 | 11 | 1.1E+08 | G/T | *C11orf87 - ZC3H12C* | 11q22.3 | 0.128 | 0.046 | 3.32 ( 1.86 - 5.94 ) | 5.18E-05 |
| rs669143 | 11 | 1.1E+08 | T/C | *C11orf87 - ZC3H12C* | 11q22.3 | 0.128 | 0.046 | 3.32 ( 1.86 - 5.94 ) | 5.18E-05 |
| rs624595 | 11 | 1.1E+08 | T/C | *C11orf87 - ZC3H12C* | 11q22.3 | 0.128 | 0.046 | 3.32 ( 1.86 - 5.94 ) | 5.18E-05 |
| rs10891023 | 11 | 1.1E+08 | G/T | *C11orf87 - ZC3H12C* | 11q22.3 | 0.128 | 0.046 | 3.32 ( 1.86 - 5.94 ) | 5.18E-05 |
| rs486909 | 11 | 1.1E+08 | T/C | *C11orf87 - ZC3H12C* | 11q22.3 | 0.128 | 0.046 | 3.32 ( 1.86 - 5.94 ) | 5.18E-05 |
| rs566895 | 11 | 1.1E+08 | T/C | *C11orf87 - ZC3H12C* | 11q22.3 | 0.128 | 0.046 | 3.32 ( 1.86 - 5.94 ) | 5.18E-05 |
| rs548021 | 11 | 1.1E+08 | A/G | *C11orf87 - ZC3H12C* | 11q22.3 | 0.128 | 0.046 | 3.32 ( 1.86 - 5.94 ) | 5.18E-05 |
| rs666248 | 11 | 1.1E+08 | A/C | *C11orf87 - ZC3H12C* | 11q22.3 | 0.128 | 0.046 | 3.32 ( 1.86 - 5.94 ) | 5.18E-05 |
| rs509305 | 11 | 1.1E+08 | G/A | *C11orf87 - ZC3H12C* | 11q22.3 | 0.132 | 0.046 | 3.4 ( 1.9 - 6.06 ) | 3.58E-05 |
| rs503824 | 11 | 1.1E+08 | A/C | *C11orf87 - ZC3H12C* | 11q22.3 | 0.132 | 0.046 | 3.4 ( 1.9 - 6.06 ) | 3.58E-05 |
| rs667582 | 11 | 1.1E+08 | T/C | *C11orf87 - ZC3H12C* | 11q22.3 | 0.13 | 0.044 | 3.49 ( 1.93 - 6.31 ) | 3.43E-05 |
| rs670276 | 11 | 1.1E+08 | C/T | *C11orf87 - ZC3H12C* | 11q22.3 | 0.128 | 0.044 | 3.5 ( 1.94 - 6.34 ) | 3.48E-05 |
| rs560753 | 11 | 1.1E+08 | G/A | *C11orf87 - ZC3H12C* | 11q22.3 | 0.128 | 0.044 | 3.5 ( 1.94 - 6.34 ) | 3.48E-05 |

| | | | | | | | | | |
|---|---|---|---|---|---|---|---|---|---|
| rs117047335 | 11 | 1.1E+08 | A/C | *C11orf87 - ZC3H12C* | 11q22.3 | 0.111 | 0.036 | 3.63 ( 1.91 - 6.91 ) | 8.67E-05 |
| rs114328531 | 11 | 1.1E+08 | A/G | *C11orf87 - ZC3H12C* | 11q22.3 | 0.127 | 0.044 | 3.49 ( 1.92 - 6.32 ) | 3.91E-05 |
| rs6589091 | 11 | 1.1E+08 | G/A | *C11orf87 - ZC3H12C* | 11q22.3 | 0.13 | 0.044 | 3.58 ( 1.98 - 6.47 ) | 2.48E-05 |
| rs7934780 | 11 | 1.1E+08 | A/G | *C11orf87 - ZC3H12C* | 11q22.3 | 0.13 | 0.044 | 3.58 ( 1.98 - 6.47 ) | 2.48E-05 |
| rs2126869 | 11 | 1.1E+08 | A/C | *C11orf87 - ZC3H12C* | 11q22.3 | 0.13 | 0.044 | 3.58 ( 1.98 - 6.47 ) | 2.48E-05 |
| rs4494274 | 11 | 1.1E+08 | C/T | *C11orf87 - ZC3H12C* | 11q22.3 | 0.13 | 0.044 | 3.58 ( 1.98 - 6.47 ) | 2.48E-05 |
| rs77861048 | 11 | 1.1E+08 | A/G | *C11orf87 - ZC3H12C* | 11q22.3 | 0.13 | 0.044 | 3.58 ( 1.98 - 6.47 ) | 2.48E-05 |
| rs1600113 | 11 | 1.1E+08 | C/A | *C11orf87 - ZC3H12C* | 11q22.3 | 0.132 | 0.044 | 3.56 ( 1.98 - 6.41 ) | 2.29E-05 |
| rs6589097 | 11 | 1.1E+08 | T/C | *C11orf87 - ZC3H12C* | 11q22.3 | 0.13 | 0.044 | 3.58 ( 1.98 - 6.47 ) | 2.48E-05 |
| rs1478249 | 11 | 1.1E+08 | A/G | *C11orf87 - ZC3H12C* | 11q22.3 | 0.13 | 0.044 | 3.58 ( 1.98 - 6.47 ) | 2.48E-05 |
| rs1471417 | 11 | 1.1E+08 | G/A | *C11orf87 - ZC3H12C* | 11q22.3 | 0.13 | 0.046 | 3.26 ( 1.83 - 5.8 ) | 5.94E-05 |
| rs7106052 | 11 | 1.1E+08 | G/T | *C11orf87 - ZC3H12C* | 11q22.3 | 0.13 | 0.044 | 3.58 ( 1.98 - 6.47 ) | 2.48E-05 |
| rs1600114 | 11 | 1.1E+08 | C/T | *C11orf87 - ZC3H12C* | 11q22.3 | 0.13 | 0.044 | 3.58 ( 1.98 - 6.47 ) | 2.48E-05 |
| rs11213201 | 11 | 1.1E+08 | A/G | *C11orf87 - ZC3H12C* | 11q22.3 | 0.124 | 0.044 | 3.38 ( 1.86 - 6.15 ) | 6.60E-05 |
| rs2126870 | 11 | 1.1E+08 | C/T | *C11orf87 - ZC3H12C* | 11q22.3 | 0.132 | 0.044 | 3.49 ( 1.94 - 6.26 ) | 2.89E-05 |
| rs6589098 | 11 | 1.1E+08 | T/C | *C11orf87 - ZC3H12C* | 11q22.3 | 0.13 | 0.044 | 3.58 ( 1.98 - 6.47 ) | 2.48E-05 |
| rs1478247 | 11 | 1.1E+08 | C/T | *C11orf87 - ZC3H12C* | 11q22.3 | 0.13 | 0.044 | 3.58 ( 1.98 - 6.47 ) | 2.48E-05 |
| rs1478248 | 11 | 1.1E+08 | C/G | *C11orf87 - ZC3H12C* | 11q22.3 | 0.13 | 0.044 | 3.58 ( 1.98 - 6.47 ) | 2.48E-05 |
| rs7126496 | 11 | 1.1E+08 | C/T | *C11orf87 - ZC3H12C* | 11q22.3 | 0.13 | 0.044 | 3.58 ( 1.98 - 6.47 ) | 2.48E-05 |
| rs4754414 | 11 | 1.1E+08 | C/A | *C11orf87 - ZC3H12C* | 11q22.3 | 0.13 | 0.044 | 3.58 ( 1.98 - 6.47 ) | 2.48E-05 |
| rs961806 | 11 | 1.1E+08 | T/G | *C11orf87 - ZC3H12C* | 11q22.3 | 0.137 | 0.044 | 3.52 ( 1.97 - 6.29 ) | 2.14E-05 |
| rs7128879 | 11 | 1.1E+08 | G/A | *C11orf87 - ZC3H12C* | 11q22.3 | 0.128 | 0.044 | 3.48 ( 1.92 - 6.3 ) | 3.80E-05 |
| rs7950429 | 11 | 1.1E+08 | C/T | *C11orf87 - ZC3H12C* | 11q22.3 | 0.13 | 0.044 | 3.58 ( 1.98 - 6.47 ) | 2.48E-05 |
| rs10789744 | 11 | 1.1E+08 | T/A | *C11orf87 - ZC3H12C* | 11q22.3 | 0.13 | 0.044 | 3.58 ( 1.98 - 6.47 ) | 2.48E-05 |
| rs6589099 | 11 | 1.1E+08 | A/G | *C11orf87 - ZC3H12C* | 11q22.3 | 0.128 | 0.044 | 3.48 ( 1.92 - 6.3 ) | 3.80E-05 |
| rs79827329 | 12 | 20807357 | A/G | *PDE3A* | 12p12.2 | 0.08 | 0.019 | 5.83 ( 2.42 - 14 ) | 8.15E-05 |

| | | | | | | | | | |
|---|---|---|---|---|---|---|---|---|---|
| rs192313556 | 12 | 20807453 | G/C | *PDE3A* | 12p12.2 | 0.078 | 0.019 | 5.84 ( 2.41 - 14.14 ) | 9.04E-05 |
| rs7314545 | 12 | 20810280 | T/C | *PDE3A* | 12p12.2 | 0.08 | 0.019 | 5.83 ( 2.42 - 14 ) | 8.15E-05 |
| rs7965422 | 12 | 20840290 | C/T | *PDE3A - SLCO1C1* | 12p12.2 | 0.085 | 0.025 | 5.02 ( 2.26 - 11.15 ) | 7.51E-05 |
| rs7978760 | 12 | 20840292 | A/C | *PDE3A - SLCO1C1* | 12p12.2 | 0.085 | 0.025 | 5.02 ( 2.26 - 11.15 ) | 7.51E-05 |
| rs117426963 | 12 | 69768344 | G/A | *YEATS4* | 12q15 | 0.717 | 0.593 | 1.82 ( 1.35 - 2.45 ) | 9.86E-05 |
| rs75311705 | 12 | 1.26E+08 | G/A | *AACS - TMEM132B* | 12q24.31 | 0.946 | 0.866 | 2.82 ( 1.69 - 4.71 ) | 7.42E-05 |
| rs8002175 | 13 | 28680073 | G/C | *FLT3 - PAN3-AS1* | 13q12.2 | 0.307 | 0.191 | 1.97 ( 1.41 - 2.76 ) | 6.98E-05 |
| rs55988011 | 13 | 94464404 | C/G | *GPC6* | 13q31.3 | 0.412 | 0.276 | 1.77 ( 1.35 - 2.32 ) | 3.54E-05 |
| rs11158268 | 14 | 60199508 | T/A | *RTN1* | 14q23.1 | 0.53 | 0.382 | 1.74 ( 1.32 - 2.3 ) | 7.67E-05 |
| rs754304 | 14 | 1.01E+08 | C/G | *MEG8* | 14q32.2 | 0.735 | 0.62 | 1.82 ( 1.35 - 2.46 ) | 8.71E-05 |
| rs74082829 | 14 | 1.01E+08 | G/A | *MEG8* | 14q32.2 | 0.896 | 0.787 | 2.33 ( 1.57 - 3.45 ) | 2.78E-05 |
| rs7160787 | 14 | 1.01E+08 | C/T | *MEG8* | 14q32.2 | 0.906 | 0.795 | 2.4 ( 1.6 - 3.59 ) | 2.13E-05 |
| rs7146460 | 14 | 1.01E+08 | C/T | *MEG8* | 14q32.2 | 0.898 | 0.792 | 2.27 ( 1.53 - 3.36 ) | 4.89E-05 |
| rs1815262 | 14 | 1.03E+08 | T/C | *LINC02323 - RCOR1* | 14q32.31 | 0.693 | 0.555 | 1.99 ( 1.46 - 2.72 ) | 1.33E-05 |
| rs749543 | 14 | 1.03E+08 | A/C | *LINC02323 - RCOR1* | 14q32.31 | 0.689 | 0.555 | 1.99 ( 1.45 - 2.72 ) | 1.60E-05 |
| rs4906229 | 14 | 1.03E+08 | C/A | *LINC02323 - RCOR1* | 14q32.31 | 0.689 | 0.557 | 1.95 ( 1.43 - 2.66 ) | 2.33E-05 |
| rs10132160 | 14 | 1.03E+08 | G/A | *RCOR1* | 14q32.31 | 0.683 | 0.555 | 1.9 ( 1.4 - 2.59 ) | 3.84E-05 |
| rs8009475 | 14 | 1.03E+08 | G/A | *RCOR1* | 14q32.31 | 0.686 | 0.544 | 2.02 ( 1.49 - 2.74 ) | 7.31E-06 |
| rs3825565 | 14 | 1.03E+08 | A/G | *RCOR1* | 14q32.31 | 0.674 | 0.546 | 1.91 ( 1.4 - 2.59 ) | 4.15E-05 |
| rs4906231 | 14 | 1.03E+08 | C/T | *RCOR1* | 14q32.31 | 0.678 | 0.544 | 1.95 ( 1.44 - 2.65 ) | 1.99E-05 |
| rs10144778 | 14 | 1.03E+08 | G/C | *RCOR1* | 14q32.31 | 0.674 | 0.549 | 1.9 ( 1.4 - 2.58 ) | 4.29E-05 |
| rs7154538 | 14 | 1.03E+08 | C/T | *RCOR1* | 14q32.31 | 0.68 | 0.549 | 1.94 ( 1.43 - 2.65 ) | 2.51E-05 |
| rs3950014 | 14 | 1.03E+08 | A/G | *RCOR1* | 14q32.31 | 0.676 | 0.546 | 1.9 ( 1.4 - 2.58 ) | 3.88E-05 |
| rs2146429 | 14 | 1.03E+08 | T/A | *RCOR1* | 14q32.31 | 0.676 | 0.546 | 1.92 ( 1.41 - 2.61 ) | 3.37E-05 |
| rs2146430 | 14 | 1.03E+08 | T/G | *RCOR1* | 14q32.31 | 0.676 | 0.546 | 1.92 ( 1.41 - 2.61 ) | 3.37E-05 |
| rs2146431 | 14 | 1.03E+08 | A/G | *RCOR1* | 14q32.31 | 0.676 | 0.546 | 1.92 ( 1.41 - 2.61 ) | 3.37E-05 |

| | | | | | | | | | |
|---|---|---|---|---|---|---|---|---|---|
| rs2403066 | 14 | 1.03E+08 | C/T | *RCOR1* | 14q32.31 | 0.674 | 0.546 | 1.9 ( 1.39 - 2.57 ) | 4.31E-05 |
| rs2403067 | 14 | 1.03E+08 | G/C | *RCOR1* | 14q32.31 | 0.676 | 0.546 | 1.92 ( 1.41 - 2.61 ) | 3.37E-05 |
| rs2403068 | 14 | 1.03E+08 | A/G | *RCOR1* | 14q32.31 | 0.676 | 0.549 | 1.89 ( 1.39 - 2.56 ) | 5.11E-05 |
| rs12185065 | 14 | 1.03E+08 | T/G | *RCOR1* | 14q32.31 | 0.677 | 0.544 | 1.94 ( 1.42 - 2.63 ) | 2.46E-05 |
| rs7144086 | 14 | 1.03E+08 | C/T | *RCOR1* | 14q32.31 | 0.67 | 0.549 | 1.83 ( 1.35 - 2.48 ) | 9.70E-05 |
| rs2896450 | 14 | 1.03E+08 | C/T | *RCOR1* | 14q32.31 | 0.68 | 0.549 | 1.94 ( 1.43 - 2.65 ) | 2.51E-05 |
| rs1951630 | 14 | 1.03E+08 | T/A | *RCOR1* | 14q32.31 | 0.678 | 0.549 | 1.93 ( 1.42 - 2.63 ) | 3.08E-05 |
| rs4906234 | 14 | 1.03E+08 | C/G | *RCOR1* | 14q32.31 | 0.678 | 0.546 | 1.96 ( 1.44 - 2.67 ) | 2.13E-05 |
| rs4906235 | 14 | 1.03E+08 | C/T | *RCOR1* | 14q32.31 | 0.678 | 0.546 | 1.96 ( 1.44 - 2.67 ) | 2.13E-05 |
| rs4906236 | 14 | 1.03E+08 | A/G | *RCOR1* | 14q32.31 | 0.678 | 0.549 | 1.93 ( 1.42 - 2.63 ) | 3.08E-05 |
| rs4906237 | 14 | 1.03E+08 | G/T | *RCOR1* | 14q32.31 | 0.676 | 0.549 | 1.9 ( 1.4 - 2.58 ) | 4.26E-05 |
| rs4906238 | 14 | 1.03E+08 | A/G | *RCOR1* | 14q32.31 | 0.678 | 0.546 | 1.93 ( 1.42 - 2.63 ) | 2.68E-05 |
| rs4906239 | 14 | 1.03E+08 | G/A | *RCOR1* | 14q32.31 | 0.678 | 0.546 | 1.96 ( 1.44 - 2.66 ) | 2.06E-05 |
| rs2403069 | 14 | 1.03E+08 | T/C | *RCOR1* | 14q32.31 | 0.676 | 0.549 | 1.89 ( 1.39 - 2.57 ) | 4.64E-05 |
| rs4509953 | 14 | 1.03E+08 | G/C | *RCOR1* | 14q32.31 | 0.675 | 0.546 | 1.9 ( 1.4 - 2.59 ) | 3.93E-05 |
| rs1174256160 | 14 | 1.03E+08 | */G | . | . | 0.723 | 0.586 | 2.01 ( 1.45 - 2.78 ) | 2.64E-05 |
| rs12887323 | 14 | 1.03E+08 | C/T | *RCOR1* | 14q32.31 | 0.68 | 0.549 | 1.94 ( 1.43 - 2.65 ) | 2.51E-05 |
| rs12887440 | 14 | 1.03E+08 | A/G | *RCOR1* | 14q32.31 | 0.676 | 0.53 | 1.98 ( 1.47 - 2.66 ) | 7.00E-06 |
| rs4906242 | 14 | 1.03E+08 | A/G | *RCOR1* | 14q32.31 | 0.68 | 0.549 | 1.94 ( 1.43 - 2.65 ) | 2.51E-05 |
| rs10144272 | 14 | 1.03E+08 | G/T | *RCOR1* | 14q32.31 | 0.678 | 0.549 | 1.93 ( 1.42 - 2.63 ) | 3.08E-05 |
| rs11626481 | 14 | 1.03E+08 | C/T | *RCOR1* | 14q32.31 | 0.68 | 0.552 | 1.91 ( 1.4 - 2.59 ) | 3.95E-05 |
| rs7153340 | 14 | 1.03E+08 | C/T | *RCOR1* | 14q32.31 | 0.678 | 0.549 | 1.93 ( 1.42 - 2.63 ) | 3.08E-05 |
| rs4906245 | 14 | 1.03E+08 | C/G | *RCOR1* | 14q32.31 | 0.676 | 0.549 | 1.9 ( 1.4 - 2.58 ) | 4.37E-05 |
| rs12588817 | 14 | 1.03E+08 | A/G | *RCOR1* | 14q32.31 | 0.678 | 0.546 | 1.94 ( 1.43 - 2.65 ) | 2.35E-05 |
| rs10150990 | 14 | 1.03E+08 | G/A | *RCOR1* | 14q32.31 | 0.676 | 0.546 | 1.91 ( 1.41 - 2.6 ) | 3.32E-05 |
| rs10131189 | 14 | 1.03E+08 | G/C | *RCOR1* | 14q32.31 | 0.678 | 0.546 | 1.94 ( 1.42 - 2.64 ) | 2.61E-05 |

| | | | | | | | | | |
|---|---|---|---|---|---|---|---|---|---|
| rs2403070 | 14 | 1.03E+08 | C/T | *RCOR1* | 14q32.31 | 0.678 | 0.546 | 1.94 ( 1.42 - 2.64 ) | 2.57E-05 |
| rs57749886 | 14 | 1.03E+08 | T/C | *RCOR1* | 14q32.31 | 0.676 | 0.546 | 1.95 ( 1.43 - 2.66 ) | 2.46E-05 |
| rs12884619 | 14 | 1.03E+08 | A/C | *RCOR1* | 14q32.31 | 0.676 | 0.538 | 2.04 ( 1.49 - 2.79 ) | 7.55E-06 |
| rs9324049 | 14 | 1.03E+08 | C/T | *RCOR1* | 14q32.31 | 0.678 | 0.549 | 1.93 ( 1.42 - 2.63 ) | 3.08E-05 |
| rs11627756 | 14 | 1.03E+08 | T/C | *RCOR1* | 14q32.31 | 0.68 | 0.549 | 1.94 ( 1.42 - 2.64 ) | 2.59E-05 |
| rs1190344 | 14 | 1.03E+08 | G/A | *RCOR1* | 14q32.31 | 0.676 | 0.549 | 1.91 ( 1.4 - 2.61 ) | 3.89E-05 |
| rs1190343 | 14 | 1.03E+08 | A/G | *RCOR1* | 14q32.31 | 0.678 | 0.549 | 1.93 ( 1.42 - 2.63 ) | 3.08E-05 |
| rs1190342 | 14 | 1.03E+08 | T/C | *RCOR1* | 14q32.31 | 0.68 | 0.549 | 1.94 ( 1.43 - 2.65 ) | 2.51E-05 |
| rs1190341 | 14 | 1.03E+08 | G/A | *RCOR1* | 14q32.31 | 0.676 | 0.552 | 1.87 ( 1.38 - 2.55 ) | 6.04E-05 |
| rs1190340 | 14 | 1.03E+08 | G/T | *RCOR1* | 14q32.31 | 0.676 | 0.546 | 1.9 ( 1.4 - 2.58 ) | 3.51E-05 |
| rs1190339 | 14 | 1.03E+08 | T/C | *RCOR1* | 14q32.31 | 0.678 | 0.549 | 1.93 ( 1.42 - 2.63 ) | 3.08E-05 |
| rs1190338 | 14 | 1.03E+08 | G/T | *RCOR1* | 14q32.31 | 0.668 | 0.527 | 1.86 ( 1.39 - 2.49 ) | 2.99E-05 |
| rs1190337 | 14 | 1.03E+08 | C/A | *RCOR1* | 14q32.31 | 0.68 | 0.544 | 1.98 ( 1.46 - 2.69 ) | 1.33E-05 |
| rs1190336 | 14 | 1.03E+08 | T/C | *RCOR1* | 14q32.31 | 0.674 | 0.549 | 1.89 ( 1.39 - 2.58 ) | 5.15E-05 |
| rs942022 | 14 | 1.03E+08 | T/C | *RCOR1* | 14q32.31 | 0.68 | 0.546 | 1.97 ( 1.45 - 2.68 ) | 1.63E-05 |
| rs1190335 | 14 | 1.03E+08 | C/T | *RCOR1* | 14q32.31 | 0.674 | 0.549 | 1.9 ( 1.39 - 2.59 ) | 4.92E-05 |
| rs1190334 | 14 | 1.03E+08 | T/C | *RCOR1* | 14q32.31 | 0.674 | 0.549 | 1.9 ( 1.39 - 2.59 ) | 4.92E-05 |
| rs12892602 | 14 | 1.03E+08 | T/C | *RCOR1* | 14q32.31 | 0.674 | 0.549 | 1.9 ( 1.39 - 2.59 ) | 4.92E-05 |
| rs7155804 | 14 | 1.03E+08 | A/G | *RCOR1* | 14q32.31 | 0.676 | 0.552 | 1.89 ( 1.39 - 2.57 ) | 5.36E-05 |
| rs12050120 | 14 | 1.03E+08 | C/T | *RCOR1* | 14q32.31 | 0.69 | 0.552 | 2 ( 1.47 - 2.72 ) | 1.00E-05 |
| rs8013886 | 14 | 1.03E+08 | A/G | *RCOR1* | 14q32.31 | 0.676 | 0.549 | 1.92 ( 1.41 - 2.61 ) | 3.74E-05 |
| rs7142608 | 14 | 1.03E+08 | G/C | *RCOR1* | 14q32.31 | 0.676 | 0.549 | 1.92 ( 1.41 - 2.61 ) | 3.74E-05 |
| rs7142319 | 14 | 1.03E+08 | C/T | *RCOR1* | 14q32.31 | 0.676 | 0.552 | 1.88 ( 1.38 - 2.56 ) | 5.70E-05 |
| rs7143048 | 14 | 1.03E+08 | C/T | *RCOR1* | 14q32.31 | 0.678 | 0.552 | 1.89 ( 1.39 - 2.58 ) | 4.83E-05 |
| rs8006489 | 14 | 1.03E+08 | G/A | *RCOR1* | 14q32.31 | 0.68 | 0.552 | 1.92 ( 1.41 - 2.61 ) | 3.64E-05 |
| rs7145737 | 14 | 1.03E+08 | T/A | *RCOR1* | 14q32.31 | 0.678 | 0.549 | 1.93 ( 1.42 - 2.63 ) | 3.05E-05 |

| | | | | | | | | | |
|---|---|---|---|---|---|---|---|---|---|
| rs7155883 | 14 | 1.03E+08 | A/C | *RCOR1* | 14q32.31 | 0.68 | 0.549 | 1.94 ( 1.43 - 2.65 ) | 2.51E-05 |
| rs12435280 | 14 | 1.03E+08 | A/C | *RCOR1* | 14q32.31 | 0.678 | 0.549 | 1.93 ( 1.42 - 2.63 ) | 3.08E-05 |
| rs7152144 | 14 | 1.03E+08 | C/T | *RCOR1* | 14q32.31 | 0.682 | 0.549 | 1.96 ( 1.44 - 2.66 ) | 2.05E-05 |
| rs4906248 | 14 | 1.03E+08 | A/T | *RCOR1* | 14q32.31 | 0.678 | 0.549 | 1.93 ( 1.42 - 2.63 ) | 3.08E-05 |
| rs12433517 | 14 | 1.03E+08 | T/G | *RCOR1* | 14q32.31 | 0.678 | 0.549 | 1.93 ( 1.42 - 2.63 ) | 3.08E-05 |
| rs12100848 | 14 | 1.03E+08 | T/C | *RCOR1* | 14q32.31 | 0.678 | 0.549 | 1.93 ( 1.42 - 2.63 ) | 3.08E-05 |
| rs62007925 | 14 | 1.03E+08 | G/A | *RCOR1* | 14q32.31 | 0.683 | 0.549 | 1.97 ( 1.45 - 2.67 ) | 1.61E-05 |
| rs72635192 | 14 | 1.03E+08 | C/T | *RCOR1* | 14q32.31 | 0.682 | 0.549 | 1.96 ( 1.44 - 2.67 ) | 1.96E-05 |
| rs7143783 | 14 | 1.03E+08 | A/G | *RCOR1* | 14q32.31 | 0.682 | 0.549 | 1.96 ( 1.44 - 2.66 ) | 2.06E-05 |
| rs7144340 | 14 | 1.03E+08 | G/T | *RCOR1* | 14q32.31 | 0.678 | 0.549 | 1.93 ( 1.42 - 2.63 ) | 3.08E-05 |
| rs7149667 | 14 | 1.03E+08 | G/A | *RCOR1* | 14q32.31 | 0.68 | 0.549 | 1.94 ( 1.43 - 2.65 ) | 2.51E-05 |
| rs11846451 | 14 | 1.03E+08 | T/C | *RCOR1* | 14q32.31 | 0.68 | 0.549 | 1.94 ( 1.43 - 2.65 ) | 2.48E-05 |
| rs11850368 | 14 | 1.03E+08 | G/C | *RCOR1* | 14q32.31 | 0.678 | 0.549 | 1.93 ( 1.42 - 2.63 ) | 3.08E-05 |
| rs7142819 | 14 | 1.03E+08 | T/C | *RCOR1* | 14q32.31 | 0.68 | 0.549 | 1.94 ( 1.42 - 2.64 ) | 2.59E-05 |
| rs62007927 | 14 | 1.03E+08 | C/T | *RCOR1* | 14q32.31 | 0.68 | 0.549 | 1.94 ( 1.43 - 2.65 ) | 2.51E-05 |
| rs11847793 | 14 | 1.03E+08 | A/G | *RCOR1* | 14q32.31 | 0.682 | 0.549 | 1.97 ( 1.44 - 2.68 ) | 2.00E-05 |
| rs8013957 | 14 | 1.03E+08 | C/T | *RCOR1* | 14q32.31 | 0.676 | 0.549 | 1.91 ( 1.41 - 2.61 ) | 3.84E-05 |
| rs12434778 | 14 | 1.03E+08 | G/T | *RCOR1* | 14q32.31 | 0.678 | 0.549 | 1.93 ( 1.42 - 2.63 ) | 3.08E-05 |
| rs2093259 | 14 | 1.03E+08 | T/A | *RCOR1* | 14q32.31 | 0.68 | 0.549 | 1.94 ( 1.42 - 2.64 ) | 2.59E-05 |
| rs8019811 | 14 | 1.03E+08 | A/G | *RCOR1* | 14q32.31 | 0.678 | 0.549 | 1.93 ( 1.42 - 2.63 ) | 3.08E-05 |
| rs6575929 | 14 | 1.03E+08 | T/G | *RCOR1* | 14q32.31 | 0.678 | 0.549 | 1.93 ( 1.42 - 2.63 ) | 3.08E-05 |
| rs60573537 | 14 | 1.03E+08 | A/G | *RCOR1* | 14q32.31 | 0.678 | 0.552 | 1.9 ( 1.39 - 2.58 ) | 4.71E-05 |
| rs12590775 | 14 | 1.03E+08 | A/G | *RCOR1* | 14q32.31 | 0.678 | 0.549 | 1.93 ( 1.42 - 2.63 ) | 3.08E-05 |
| rs10140498 | 14 | 1.03E+08 | A/G | *RCOR1* | 14q32.31 | 0.68 | 0.552 | 1.91 ( 1.41 - 2.6 ) | 3.60E-05 |
| rs10129838 | 14 | 1.03E+08 | T/C | *RCOR1* | 14q32.31 | 0.683 | 0.555 | 1.91 ( 1.41 - 2.6 ) | 3.41E-05 |
| rs8009257 | 14 | 1.03E+08 | A/G | *RCOR1* | 14q32.31 | 0.676 | 0.549 | 1.91 ( 1.4 - 2.6 ) | 4.12E-05 |

| | | | | | | | | | |
|---|---|---|---|---|---|---|---|---|---|
| rs8010679 | 14 | 1.03E+08 | T/G | *RCOR1* | 14q32.31 | 0.676 | 0.549 | 1.91 ( 1.4 - 2.6 ) | 4.12E-05 |
| rs11850580 | 14 | 1.03E+08 | C/T | *RCOR1* | 14q32.31 | 0.676 | 0.549 | 1.91 ( 1.4 - 2.6 ) | 4.12E-05 |
| rs8010455 | 14 | 1.03E+08 | A/G | *RCOR1* | 14q32.31 | 0.674 | 0.549 | 1.9 ( 1.4 - 2.58 ) | 4.23E-05 |
| rs4906250 | 14 | 1.03E+08 | A/G | *RCOR1* | 14q32.31 | 0.67 | 0.549 | 1.86 ( 1.37 - 2.53 ) | 6.88E-05 |
| rs11847277 | 14 | 1.03E+08 | A/G | *RCOR1* | 14q32.31 | 0.672 | 0.552 | 1.88 ( 1.38 - 2.56 ) | 6.15E-05 |
| rs10138008 | 14 | 1.03E+08 | G/C | *RCOR1 - TRAF3* | 14q32.32 | 0.671 | 0.549 | 1.83 ( 1.36 - 2.46 ) | 7.30E-05 |
| rs55773826 | 15 | 26282223 | C/T | *LINC02346* | 15q12 | 0.938 | 0.85 | 3.26 ( 1.96 - 5.42 ) | 5.29E-06 |
| . | 16 | 32833203 | A/G | *TP53TG3 - SLC6A10P* | 16p11.2 | 0.846 | 0.724 | 2.48 ( 1.68 - 3.68 ) | 5.59E-06 |
| rs4949133 | 16 | 63676226 | A/G | *CDH8 - CDH11* | 16q21 | 0.68 | 0.552 | 1.96 ( 1.43 - 2.67 ) | 2.34E-05 |
| rs12448930 | 16 | 63680366 | C/A | *CDH8 - CDH11* | 16q21 | 0.68 | 0.552 | 1.96 ( 1.43 - 2.67 ) | 2.34E-05 |
| rs7189633 | 16 | 63687513 | T/A | *CDH8 - CDH11* | 16q21 | 0.663 | 0.538 | 1.95 ( 1.43 - 2.65 ) | 2.34E-05 |
| rs1839853 | 16 | 63688705 | A/G | *CDH8 - CDH11* | 16q21 | 0.665 | 0.544 | 1.93 ( 1.41 - 2.64 ) | 3.42E-05 |
| rs1823657 | 16 | 63690757 | T/A | *CDH8 - CDH11* | 16q21 | 0.659 | 0.541 | 1.86 ( 1.37 - 2.52 ) | 6.70E-05 |
| rs28470866 | 17 | 13537630 | A/G | *HS3ST3A1 - CDRT15P1* | 17p12 | 0.35 | 0.227 | 2.02 ( 1.45 - 2.83 ) | 4.11E-05 |
| rs56199331 | 17 | 13538760 | G/T | *HS3ST3A1 - CDRT15P1* | 17p12 | 0.346 | 0.224 | 1.98 ( 1.42 - 2.77 ) | 6.23E-05 |
| rs59576814 | 17 | 13538761 | C/T | *HS3ST3A1 - CDRT15P1* | 17p12 | 0.346 | 0.224 | 1.98 ( 1.42 - 2.77 ) | 6.23E-05 |
| rs8080210 | 17 | 13539465 | A/G | *HS3ST3A1 - CDRT15P1* | 17p12 | 0.348 | 0.23 | 1.97 ( 1.41 - 2.76 ) | 7.29E-05 |
| rs727547 | 17 | 13540397 | G/A | *HS3ST3A1 - CDRT15P1* | 17p12 | 0.35 | 0.23 | 1.99 ( 1.42 - 2.79 ) | 5.91E-05 |
| rs727546 | 17 | 13540518 | G/A | *HS3ST3A1 - CDRT15P1* | 17p12 | 0.35 | 0.23 | 1.99 ( 1.42 - 2.79 ) | 5.91E-05 |
| rs7215641 | 17 | 16368752 | A/G | *LRRC75A-AS1* | 17p11.2 | 0.822 | 0.716 | 2.02 ( 1.42 - 2.88 ) | 9.08E-05 |
| rs4143830 | 17 | 16370019 | G/A | *LRRC75A-AS1* | 17p11.2 | 0.846 | 0.74 | 2.11 ( 1.47 - 3.03 ) | 4.92E-05 |
| rs317326 | 17 | 32161502 | C/A | *ASIC2* | 17q12 | 0.168 | 0.074 | 2.69 ( 1.68 - 4.33 ) | 4.26E-05 |
| rs1881143 | 17 | 48354583 | G/A | *TMEM92* | 17q21.33 | 0.593 | 0.462 | 1.81 ( 1.36 - 2.42 ) | 5.26E-05 |
| rs576604 | 18 | 44365981 | C/G | *ST8SIA5 - PIAS2* | 18q21.1 | 0.535 | 0.407 | 1.79 ( 1.34 - 2.38 ) | 6.78E-05 |
| rs3179015 | 19 | 15347829 | A/* | . | . | 0.879 | 0.783 | 2.61 ( 1.68 - 4.05 ) | 1.92E-05 |
| rs10418981 | 19 | 54100891 | T/C | *ZNF331 - LOC284379* | 19q13.42 | 0.926 | 0.825 | 2.59 ( 1.65 - 4.06 ) | 3.56E-05 |

| | | | | | | | | | |
|---|---|---|---|---|---|---|---|---|---|
| rs7508158 | 19 | 54100947 | G/A | *ZNF331 - LOC284379* | 19q13.42 | 0.924 | 0.828 | 2.5 ( 1.59 - 3.91 ) | 6.60E-05 |
| rs2163841 | 19 | 54101014 | A/G | *ZNF331 - LOC284379* | 19q13.42 | 0.924 | 0.831 | 2.45 ( 1.56 - 3.85 ) | 9.33E-05 |
| rs6029258 | 20 | 39284775 | G/A | *LINC01370 - MAFB* | 20q12 | 0.541 | 0.424 | 1.78 ( 1.33 - 2.37 ) | 9.31E-05 |
| rs2281366 | 20 | 45808926 | A/G | *EYA2* | 20q13.12 | 0.302 | 0.191 | 1.99 ( 1.41 - 2.81 ) | 9.38E-05 |
| rs910248026 | 22 | 38471990 | G/C | *PICK1* | 22q13.1 | 0.911 | 0.831 | 2.62 ( 1.63 - 4.2 ) | 7.12E-05 |
| rs7284558 | 22 | 48885052 | C/T | *FAM19A5* | 22q13.32 | 0.433 | 0.273 | 1.8 ( 1.35 - 2.4 ) | 6.78E-05 |
| rs16999576 | 23 | 1.29E+08 | A/T | *NONE - NONE* | . | 0.763 | 0.599 | 2.46 ( 1.6 - 3.78 ) | 3.86E-05 |
| rs60019963 | 23 | 1.29E+08 | T/C | *NONE - NONE* | . | 0.763 | 0.599 | 2.45 ( 1.6 - 3.77 ) | 4.15E-05 |
| rs113426472 | 23 | 1.29E+08 | A/G | *NONE - NONE* | . | 0.759 | 0.597 | 2.41 ( 1.57 - 3.7 ) | 5.50E-05 |
| rs5920197 | 23 | 1.46E+08 | A/G | *NONE - NONE* | . | 0.87 | 0.727 | 2.76 ( 1.66 - 4.59 ) | 8.71E-05 |
| rs4827722 | 23 | 1.46E+08 | A/G | *NONE - NONE* | . | 0.885 | 0.743 | 2.85 ( 1.68 - 4.83 ) | 9.53E-05 |
| rs4827723 | 23 | 1.46E+08 | T/C | *NONE - NONE* | . | 0.885 | 0.742 | 2.87 ( 1.69 - 4.85 ) | 9.04E-05 |
| rs140546828 | 23 | 1.52E+08 | A/G | *NONE - NONE* | . | 0.922 | 0.797 | 3.35 ( 1.83 - 6.11 ) | 8.45E-05 |

[a] The positions of SNVs are based on NCBI human genome reference sequence Build 37

[b] A1 is risk-associated allele, and A2 is non-risk-associated allele.

### Appendix Table 3 SNVs with *P* < 0.05 in the G-NG group of replication stage

| | | Position | | | | Frequency of A1 | | | |
|---|---|---|---|---|---|---|---|---|---|
| **Rs id** | **Chr** | **Bp[a]** | **A1/A2[b]** | **Gene** | **Locus** | **Cases** | **Controls** | **OR (95% CI)** | **P value** |
| rs879688782 | 1 | 52348092 | A/C | *NRDC - RAB3B* | 1p32.3 | 0.981 | 0.969 | 1.64 ( 1.07 - 2.53 ) | 2.34E-02 |
| rs7546310 | 1 | 232061820 | A/C | *DISC1* | 1q42.2 | 0.754 | 0.73 | 1.16 ( 1 - 1.33 ) | 4.45E-02 |
| rs868933181 | 3 | 51519910 | T/A | *VPRBP* | 3p21.2 | 0.882 | 0.838 | 1.54 ( 1.28 - 1.85 ) | 5.88E-06 |
| rs6777548 | 3 | 81427224 | G/A | *LINC02027 - GBE1* | 3p12.2 | 0.679 | 0.643 | 1.2 ( 1.05 - 1.37 ) | 5.84E-03 |
| rs59961674 | 4 | 88919362 | C/G | *SPP1 - PKD2* | 4q22.1 | 0.649 | 0.573 | 1.74 ( 1.49 - 2.03 ) | 3.56E-12 |
| rs2725263 | 4 | 89026428 | C/A | *ABCG2* | 4q22.1 | 0.672 | 0.545 | 1.85 ( 1.63 - 2.11 ) | 2.34E-20 |
| rs1481012 | 4 | 89039082 | G/A | *ABCG2* | 4q22.1 | 0.512 | 0.324 | 2.52 ( 2.2 - 2.88 ) | 1.24E-41 |

| | | | | | | | | | |
|---|---|---|---|---|---|---|---|---|---|
| rs1871744 | 4 | 89039629 | T/C | *ABCG2* | 4q22.1 | 0.831 | 0.7 | 2.34 ( 2 - 2.74 ) | 1.44E-26 |
| rs45499402 | 4 | 89043634 | C/G | *ABCG2* | 4q22.1 | 0.52 | 0.349 | 2.38 ( 2.08 - 2.72 ) | 3.77E-36 |
| rs4148155 | 4 | 89054667 | G/A | *ABCG2* | 4q22.1 | 0.523 | 0.333 | 2.49 ( 2.18 - 2.85 ) | 9.83E-42 |
| rs12649505 | 4 | 89108106 | A/G | *ABCG2* | 4q22.1 | 0.84 | 0.744 | 1.9 ( 1.62 - 2.22 ) | 1.04E-15 |
| rs118013835 | 4 | 89128183 | A/G | *ABCG2* | 4q22.1 | 0.165 | 0.085 | 2.17 ( 1.82 - 2.59 ) | 5.67E-18 |
| rs147510135 | 4 | 89135089 | T/C | *ABCG2* | 4q22.1 | 0.199 | 0.109 | 2.12 ( 1.8 - 2.5 ) | 2.56E-19 |
| rs35213808 | 4 | 163250472 | T/A | *FSTL5 - MIR4454* | 4q32.2 | 0.714 | 0.677 | 1.23 ( 1.07 - 1.42 ) | 2.87E-03 |
| rs2160000 | 7 | 36513617 | C/A | *ANLN - AOAH* | 7p14.2 | 0.922 | 0.892 | 1.46 ( 1.18 - 1.81 ) | 4.29E-04 |
| rs961806 | 11 | 109804887 | G/T | *C11orf87 - ZC3H12C* | 11q22.3 | 0.911 | 0.89 | 1.27 ( 1.04 - 1.56 ) | 1.76E-02 |
| rs10859907 | 12 | 95995906 | C/T | *USP44 - PGAM1P5* | 12q22 | 0.593 | 0.567 | 1.14 ( 1 - 1.3 ) | 4.77E-02 |
| rs116882606 | 12 | 128851012 | G/T | *TMEM132C* | 12q24.32 | 0.937 | 0.918 | 1.34 ( 1.06 - 1.68 ) | 1.38E-02 |
| rs8009475 | 14 | 103058608 | G/A | *RCOR1* | 14q32.31 | 0.653 | 0.615 | 1.21 ( 1.06 - 1.38 ) | 4.58E-03 |
| rs35785423 | 14 | 103074798 | A/G | *RCOR1* | 14q32.31 | 0.604 | 0.572 | 1.20 (1.05 - 1.38) | 9.08E-03 |
| rs12887440 | 14 | 103083899 | A/G | *RCOR1* | 14q32.31 | 0.646 | 0.609 | 1.22 ( 1.07 - 1.39 ) | 3.76E-03 |
| rs4143830 | 17 | 16370019 | G/A | *LRRC75A-AS1* | 17p11.2 | 0.806 | 0.783 | 1.18 ( 1.01 - 1.37 ) | 3.67E-02 |
| rs11867955 | 17 | 33570468 | T/C | *SLFN5* | 17q12 | 0.862 | 0.839 | 1.21 ( 1.02 - 1.44 ) | 2.49E-02 |
| rs6051348 | 20 | 2717526 | C/T | *EBF4* | 20p13 | 0.77 | 0.742 | 1.22 ( 1.05 - 1.43 ) | 1.12E-02 |
| rs72626599 | 20 | 56320267 | G/A | *NKILA - MIR4532* | 20q13.31 | 0.78 | 0.749 | 1.22 ( 1.05 - 1.42 ) | 7.92E-03 |
| rs7284558 | 22 | 48885052 | T/C | *FAM19A5* | 22q13.32 | 0.617 | 0.582 | 1.19 ( 1.05 - 1.36 ) | 8.35E-03 |

[a] The positions of SNVs are based on NCBI human genome reference sequence Build 37

[b] A1 is risk-associated allele, and A2 is non-risk-associated allele.

### Appendix Table 4 SNVs with $P < 0.05$ in the G-H group of replication stage

| | | Position | | | | Frequency of A1 | | | |
|---|---|---|---|---|---|---|---|---|---|
| Rs id | Chr | Bp[a] | A1/A2[b] | Gene | Locus | Cases | Controls | OR (95% CI) | P value |
| rs879688782 | 1 | 52348092 | C/A | *NRDC - RAB3B* | 1p32.3 | 0.0192 | 0.00894 | 2.19 ( 1.14 - 4.22 ) | 1.85E-02 |
| rs6680315 | 1 | 94370519 | C/G | *GCLM* | 1p22.1 | 0.777 | 0.741 | 1.28 ( 1.08 - 1.52 ) | 5.22E-03 |
| rs755104 | 1 | 221569202 | A/G | *C1orf140 - DUSP10* | 1q41 | 0.733 | 0.699 | 1.23 ( 1.05 - 1.46 ) | 1.30E-02 |
| rs7546310 | 1 | 232061820 | A/C | *DISC1* | 1q42.2 | 0.754 | 0.7 | 1.4 ( 1.18 - 1.65 ) | 7.47E-05 |
| rs6777548 | 3 | 81427224 | G/A | *LINC02027 - GBE1* | 3p12.2 | 0.679 | 0.63 | 1.3 ( 1.11 - 1.51 ) | 9.88E-04 |
| rs59961674 | 4 | 88919362 | C/G | *SPP1 - PKD2* | 4q22.1 | 0.649 | 0.578 | 1.69 ( 1.41 - 2.03 ) | 2.11E-08 |
| rs2725263 | 4 | 89026428 | C/A | *ABCG2* | 4q22.1 | 0.672 | 0.575 | 1.67 ( 1.43 - 1.95 ) | 9.84E-11 |
| rs1481012 | 4 | 89039082 | G/A | *ABCG2* | 4q22.1 | 0.512 | 0.392 | 1.85 ( 1.58 - 2.16 ) | 7.45E-15 |
| rs1871744 | 4 | 89039629 | T/C | *ABCG2* | 4q22.1 | 0.831 | 0.722 | 2.12 ( 1.77 - 2.54 ) | 3.22E-16 |
| rs45499402 | 4 | 89043634 | C/G | *ABCG2* | 4q22.1 | 0.52 | 0.408 | 1.79 ( 1.53 - 2.1 ) | 2.15E-13 |
| rs4148155 | 4 | 89054667 | G/A | *ABCG2* | 4q22.1 | 0.523 | 0.4 | 1.85 ( 1.58 - 2.15 ) | 3.71E-15 |
| rs12649505 | 4 | 89108106 | A/G | *ABCG2* | 4q22.1 | 0.84 | 0.762 | 1.77 ( 1.47 - 2.12 ) | 1.36E-09 |
| rs118013835 | 4 | 89128183 | A/G | *ABCG2* | 4q22.1 | 0.165 | 0.103 | 1.77 ( 1.44 - 2.17 ) | 4.84E-08 |
| rs147510135 | 4 | 89135089 | T/C | *ABCG2* | 4q22.1 | 0.199 | 0.128 | 1.77 ( 1.46 - 2.14 ) | 5.55E-09 |
| rs35213808 | 4 | 163250472 | T/A | *FSTL5 - MIR4454* | 4q32.2 | 0.714 | 0.656 | 1.4 ( 1.19 - 1.65 ) | 3.84E-05 |
| rs141876887 | 4 | 181972398 | T/C | *NONE - LINC00290* | 4q34.3 | 0.908 | 0.881 | 1.37 ( 1.09 - 1.71 ) | 6.55E-03 |
| rs12659215 | 5 | 121605926 | A/G | *LOC100505841 - SNCAIP* | 5q23.2 | 0.894 | 0.872 | 1.25 ( 1.01 - 1.54 ) | 4.22E-02 |
| rs2160000 | 7 | 36513617 | C/A | *ANLN - AOAH* | 7p14.2 | 0.922 | 0.889 | 1.51 ( 1.19 - 1.91 ) | 7.22E-04 |
| rs73158732 | 7 | 136606172 | A/C | *LOC349160* | 7q33 | 0.632 | 0.601 | 1.17 ( 1 - 1.36 ) | 4.30E-02 |
| rs601853 | 10 | 14393558 | C/T | *FRMD4A - MIR4293* | 10p13 | 0.624 | 0.596 | 1.19 ( 1.01 - 1.39 ) | 3.91E-02 |
| rs7896390 | 10 | 113516837 | T/C | *ADRA2A - GPAM* | 10q25.2 | 0.782 | 0.752 | 1.22 ( 1.03 - 1.45 ) | 2.25E-02 |
| rs961806 | 11 | 109804887 | G/T | *C11orf87 - ZC3H12C* | 11q22.3 | 0.911 | 0.882 | 1.4 ( 1.11 - 1.75 ) | 4.04E-03 |
| rs10735402 | 12 | 105708888 | A/G | *KCCAT198* | 12q23.3 | 0.666 | 0.628 | 1.24 ( 1.06 - 1.45 ) | 8.26E-03 |

| | | | | | | | | | |
|---|---|---|---|---|---|---|---|---|---|
| rs116882606 | 12 | 128851012 | G/T | *TMEM132C* | 12q24.32 | 0.937 | 0.905 | 1.59 ( 1.23 - 2.05 ) | 4.03E-04 |
| rs7160787 | 14 | 101360168 | C/T | *MEG8* | 14q32.2 | 0.796 | 0.749 | 1.39 ( 1.16 - 1.66 ) | 2.71E-04 |
| rs8009475 | 14 | 103058608 | G/A | *RCOR1* | 14q32.31 | 0.653 | 0.595 | 1.36 ( 1.17 - 1.6 ) | 1.16E-04 |
| rs35258120 | 14 | 103058769 | A/G | *RCOR1* | 14q32.31 | 0.65 | 0.615 | 1.18 (1.02 - 1.36) | 2.64E-02 |
| rs35785423 | 14 | 103074798 | A/G | *RCOR1* | 14q32.31 | 0.604 | 0.563 | 1.28 (1.08 - 1.5) | 3.13E-03 |
| rs12887440 | 14 | 103083899 | A/G | *RCOR1* | 14q32.31 | 0.646 | 0.59 | 1.37 ( 1.17 - 1.6 ) | 1.15E-04 |
| rs4949133 | 16 | 63676226 | A/G | *CDH8 - CDH11* | 16q21 | 0.648 | 0.608 | 1.24 ( 1.06 - 1.44 ) | 6.37E-03 |
| rs4143830 | 17 | 16370019 | G/A | *LRRC75A-AS1* | 17p11.2 | 0.806 | 0.769 | 1.3 ( 1.09 - 1.55 ) | 3.99E-03 |
| rs10418981 | 19 | 54100891 | T/C | *ZNF331 - LOC284379* | 19q13.42 | 0.875 | 0.85 | 1.26 ( 1.03 - 1.55 ) | 2.49E-02 |
| rs6051348 | 20 | 2717526 | C/T | *EBF4* | 20p13 | 0.77 | 0.733 | 1.3 ( 1.09 - 1.56 ) | 3.67E-03 |
| rs117624310 | 20 | 39153161 | T/C | *LINC01370 - MAFB* | 20q12 | 0.866 | 0.843 | 1.23 ( 1 - 1.5 ) | 4.57E-02 |
| rs72626599 | 20 | 56320267 | G/A | *NKILA - MIR4532* | 20q13.31 | 0.78 | 0.718 | 1.52 ( 1.27 - 1.81 ) | 3.00E-06 |
| rs7284558 | 22 | 48885052 | T/C | *FAM19A5* | 22q13.32 | 0.617 | 0.56 | 1.34 ( 1.15 - 1.55 ) | 1.77E-04 |

[a] The positions of SNVs are based on NCBI human genome reference sequence Build 37

[b] A1 is risk-associated allele, and A2 is non-risk-associated allele.

### Appendix Table 5 SNVs associated with gout at a genome-wide level of significance

| | | | | | **GWAS** | | | | | | | **Replication** | | | | | | | **Meta** | | | |
|---|---|---|---|---|---|---|---|---|---|---|---|---|---|---|---|---|---|---|---|---|---|---|
| | **Position** | | | | **Frequency of A1** | | | **G-NG** | | **G-H** | | **Frequency of A1** | | | **G-NG** | | **G-H** | | **G-NG** | | **G-H** | |
| **Rs id** | **Chr** | **Bp[a]** | **Gene** | **A1/ A2[b]** | **G** | **H** | **NG** | **OR (95% CI)** | **P value** | **OR (95% CI)** | **P value** | **G** | **H** | **NG** | **OR (95% CI)** | **P value** | **OR (95% CI)** | **P value** | **OR (95% CI)** | **P value** | **OR (95% CI)** | **P value** |
| rs868933181 | 3 | 51519910 | *VPRBP* | T/A | 0.918 | 0.842 | 0.890 | 2.46 ( 1.61 - 3.77 ) | 3.41E-05 | 2.32 ( 1.46 - 3.71 ) | 0.000411 | 0.882 | 0.862 | 0.838 | 1.54 ( 1.28 - 1.85 ) | 5.88E-06 | 1.22 ( 0.99 - 1.52 ) | 0.06662 | 1.66 ( 1.4 - 1.96 ) | **6.27E-09** | 1.37 ( 1.13 - 1.67 ) | 0.001654 |

| | | | | | | | | | | | | | | | | | | | | | | |
|---|---|---|---|---|---|---|---|---|---|---|---|---|---|---|---|---|---|---|---|---|---|---|
| rs59961674 | 4 | 88919362 | *SPP1-PKD2* | C/G | 0.715 | 0.607 | 0.571 | 1.80 ( 1.38 - 2.33 ) | 1.21E-05 | 1.60 ( 1.19 - 2.16 ) | 0.001864 | 0.649 | 0.578 | 0.573 | 1.74 ( 1.49 - 2.03 ) | 3.56E-12 | 1.69 ( 1.41 - 2.03 ) | 2.11E-08 | 1.75 ( 1.53 - 2.00 ) | 2.16E-16 | 1.67 ( 1.42 - 1.95 ) | 1.54E-10 |
| rs2725263 | 4 | 89026428 | *ABCG2* | C/A | 0.737 | 0.590 | 0.537 | 2.36 ( 1.80 - 3.09 ) | 3.94E-10 | 1.98 ( 1.46 - 2.69 ) | 1.11E-05 | 0.672 | 0.575 | 0.545 | 1.85 ( 1.62 - 2.11 ) | 2.33E-20 | 1.67 ( 1.43 - 1.95 ) | 9.83E-11 | 1.94 ( 1.72 - 2.18 ) | 2.18E-28 | 1.73 ( 1.51 - 1.99 ) | 8.60E-15 |
| rs1481012 | 4 | 89039082 | *ABCG2* | G/A | 0.598 | 0.336 | 0.284 | 3.92 ( 2.89 - 5.33 ) | 2.49E-18 | 3.23 ( 2.33 - 4.5 ) | 3.16E-12 | 0.512 | 0.392 | 0.324 | 2.52 ( 2.2 - 2.88 ) | 1.24E-41 | 1.85 ( 1.58 - 2.16 ) | 7.45E-15 | 2.7 ( 2.39 - 3.06 ) | 8.70E-57 | 2.04 ( 1.78 - 2.35 ) | 1.52E-23 |
| rs1871744 | 4 | 89039629 | *ABCG2* | T/C | 0.900 | 0.751 | 0.720 | 3.32 ( 2.30 - 4.79 ) | 1.65E-10 | 3.16 ( 2.10 - 4.75 ) | 3.79E-08 | 0.831 | 0.722 | 0.700 | 2.34 ( 2.00 - 2.74 ) | 1.44E-26 | 2.12 ( 1.77 - 2.54 ) | 3.22E-16 | 2.47 ( 2.14 - 2.85 ) | 7.37E-35 | 2.26 ( 1.92 - 2.66 ) | 3.37E-22 |
| rs45499402 | 4 | 89043634 | *ABCG2* | C/G | 0.613 | 0.344 | 0.290 | 4.11 ( 3.02 - 5.59 ) | 2.30E-19 | 3.33 ( 2.40 - 4.63 ) | 8.41E-13 | 0.52 | 0.408 | 0.349 | 2.38 ( 2.08 - 2.72 ) | 3.77E-36 | 1.79 ( 1.53 - 2.10 ) | 2.15E-13 | 2.59 ( 2.29 - 2.94 ) | 1.33E-51 | 2.01 ( 1.74 - 2.31 ) | 3.28E-22 |
| rs4148155 | 4 | 89054667 | *ABCG2* | G/A | 0.611 | 0.342 | 0.290 | 4.02 ( 2.96 - 5.46 ) | 5.14E-19 | 3.32 ( 2.39 - 4.61 ) | 7.89E-13 | 0.523 | 0.400 | 0.333 | 2.49 ( 2.18 - 2.85 ) | 9.83E-42 | 1.85 ( 1.58 - 2.15 ) | 3.71E-15 | 2.69 ( 2.38 - 3.03 ) | 2.40E-57 | 2.05 ( 1.78 - 2.35 ) | 3.25E-24 |
| rs12649505 | 4 | 89108106 | *ABCG2* | A/G | 0.891 | 0.794 | 0.7510 | 2.22 ( 1.58 - 3.13 ) | 4.74E-06 | 1.98 ( 1.35 - 2.92 ) | 0.0005391 | 0.840 | 0.762 | 0.744 | 1.90 ( 1.62 - 2.22 ) | 1.04E-15 | 1.77 ( 1.47 - 2.12 ) | 1.36E-09 | 1.95 ( 1.69 - 2.25 ) | 3.63E-20 | 1.80 ( 1.53 - 2.13 ) | 3.42E-12 |
| rs118013835 | 4 | 89128183 | *ABCG2* | A/G | 0.174 | 0.066 | 0.070 | 2.74 ( 1.80 - 4.17 ) | 2.36E-06 | 2.85 ( 1.75 - 4.66 ) | 2.91E-05 | 0.165 | 0.103 | 0.085 | 2.17 ( 1.82 - 2.59 ) | 5.67E-18 | 1.77 ( 1.44 - 2.17 ) | 4.84E-08 | 2.25 ( 1.91 - 2.65 ) | 1.25E-22 | 1.90 ( 1.57 - 2.29 ) | 3.05E-11 |

| | | | | | | | | | | | | | | | | | | | | | | |
|---|---|---|---|---|---|---|---|---|---|---|---|---|---|---|---|---|---|---|---|---|---|---|
| rs147510135 | 4 | 89135089 | *ABCG2* | T/C | 0.226 | 0.120 | 0.109 | 2.23 ( 1.57 - 3.15 ) | 6.33E-06 | 2.02 ( 1.37 - 2.98 ) | 0.0003529 | 0.199 | 0.128 | 0.109 | 2.12 ( 1.80 - 2.50 ) | 2.56E-19 | 1.77 ( 1.46 - 2.14 ) | 5.55E-09 | 2.14 ( 1.84 - 2.48 ) | 9.05E-24 | 1.81 ( 1.53 - 2.15 ) | 9.65E-12 |
| rs11733577 | 4 | 89174193 | *ABCG2-PPM1K* | T/C | 0.650 | 0.527 | 0.522 | 1.62 ( 1.25 - 2.09 ) | 0.0002256 | 1.59 ( 1.19 - 2.12 ) | 0.001591 | 0.595 | 0.554 | 0.538 | 1.34 ( 1.17 - 1.52 ) | 1.33E-05 | 1.25 ( 1.07 - 1.45 ) | 0.005089 | 1.39 ( 1.24 - 1.56 ) | 2.67E-08 | 1.32 ( 1.15 - 1.51 ) | 7.53E-05 |
| rs35213808 | 4 | 163250472 | *FSTL5-MIR4454* | T/A | 0.756 | 0.623 | 0.691 | 1.58 ( 1.20 - 2.09 ) | 0.001123 | 1.89 ( 1.38 - 2.58 ) | 6.59E-05 | 0.714 | 0.656 | 0.677 | 1.23 ( 1.07 - 1.42 ) | 0.002865 | 1.40 ( 1.19 - 1.65 ) | 3.84E-05 | 1.30 ( 1.15 - 1.47 ) | 3.76E-05 | 1.49 ( 1.29 - 1.72 ) | **4.02E-08** |
| rs8009475 | 14 | 103058608 | *RCOR1* | G/A | 0.686 | 0.544 | 0.597 | 1.66 ( 1.27 - 2.17 ) | 0.0002278 | 2.02 ( 1.49 - 2.74 ) | 7.31E-06 | 0.653 | 0.595 | 0.615 | 1.21 ( 1.06 - 1.38 ) | 0.004586 | 1.36 ( 1.17 - 1.60 ) | 0.0001161 | 1.29 ( 1.14 - 1.45 ) | 2.98E-05 | 1.48 ( 1.29 - 1.71 ) | **4.18E-08** |
| rs12887440 | 14 | 103083899 | *RCOR1* | A/G | 0.676 | 0.53 | 0.592 | 1.61 ( 1.24 - 2.09 ) | 0.0003785 | 1.98 ( 1.47 - 2.66 ) | 7.00E-06 | 0.646 | 0.590 | 0.609 | 1.22 ( 1.07 - 1.39 ) | 0.003757 | 1.37 ( 1.17 - 1.60 ) | 0.0001145 | 1.29 ( 1.15 - 1.45 ) | 2.67E-05 | 1.48 ( 1.29 - 1.71 ) | **3.37E-08** |
| rs72626599 | 20 | 56320267 | *NKILA-MIR4532* | G/A | 0.826 | 0.705 | 0.757 | 1.86 ( 1.37 - 2.52 ) | 6.21E-05 | 1.88 ( 1.33 - 2.64 ) | 0.0003033 | 0.780 | 0.718 | 0.749 | 1.22 ( 1.05 - 1.42 ) | 0.007919 | 1.52 ( 1.27 - 1.81 ) | 3.00E-06 | 1.33 ( 1.16 - 1.52 ) | 3.34E-05 | 1.58 ( 1.36 - 1.85 ) | **6.48E-09** |

[a] The positions of SNVs are based on NCBI human genome reference sequence Build 37

[b] A1 is risk-associated allele, and A2 is non-risk-associated allele.

### Appendix Table 6 Candidate *GCLM* SNVs selected for replication

| | | | | | GWAS | | | | | | | Replication | | | | | | | Meta | | | |
|---|---|---|---|---|---|---|---|---|---|---|---|---|---|---|---|---|---|---|---|---|---|---|
| | Position | | | | Frequency of A1 | | | G-NG | | G-H | | Frequency of A1 | | | G-NG | | G-H | | G-NG | | G-H | |
| Rs id | Chr | Bp[a] | Gene | A1/A2[b] | G | H | NG | OR (95% CI) | P value | OR (95% CI) | P value | G | H | NG | OR (95% CI) | P value | OR (95% CI) | P value | OR (95% CI) | P value | OR (95% CI) | P value |
| rs7515191 | 1 | 94367097 | *GCLM* | G/A | 0.8315 | 0.724 | 0.7525 | 1.67 ( 1.23 - 2.25 ) | 0.0009172 | 1.83 ( 1.31 - 2.57 ) | 0.0004288 | 0.7594 | 0.7565 | 0.7656 | 1.02 ( 0.86 - 1.2 ) | 0.6063 | 0.9818 (0.8315 - 1.159) | 0.8282 | 1.06 ( 0.94 - 1.21 ) | 0.3542 | 1.14 ( 0.98 - 1.33 ) | 0.07982 |
| rs6680315 | 1 | 94370519 | *GCLM* | C/G | 0.8426 | 0.7459 | 0.7673 | 1.64 ( 1.2 - 2.23 ) | 0.001907 | 1.77 ( 1.25 - 2.51 ) | 0.00125 | 0.7773 | 0.7408 | 0.7577 | 1.28 ( 1.08 - 1.52 ) | 0.08535 | 0.7809 (0.6565 - 0.9289) | 0.005216 | 1.22 ( 1.07 - 1.39 ) | 0.003819 | 1.37 ( 1.17 - 1.6 ) | 0.0008086 |
| rs2273406 | 1 | 94374576 | *GCLM* | G/A | 0.9004 | 0.8121 | 0.8153 | 1.91 ( 1.34 - 2.71 ) | 0.0003231 | 1.93 ( 1.31 - 2.86 ) | 0.0009945 | 0.8037 | 0.8155 | 0.8219 | 0.91 ( 0.76 - 1.1 ) | 0.08995 | 1.093 (0.9099 - 1.314) | 0.3405 | 0.99 ( 0.86 - 1.15 ) | 0.9321 | 1.05 ( 0.89 - 1.24 ) | 0.5915 |
| rs41303970 | 1 | 94375309 | *GCLM* | G/A | 0.8981 | 0.8039 | 0.816 | 2 ( 1.39 - 2.87 ) | 0.0001824 | 2.13 ( 1.43 - 3.19 ) | 0.0002268 | 0.7915 | 0.8015 | 0.8117 | 0.93 ( 0.77 - 1.11 ) | 0.05996 | 1.079 (0.8979 - 1.296) | 0.4185 | 0.98 ( 0.85 - 1.14 ) | 0.8143 | 1.07 ( 0.91 - 1.26 ) | 0.43 |
| rs12129986 | 1 | 94399210 | *GCLM - ABCA4* | A/G | 0.9 | 0.7978 | 0.8125 | 2.19 ( 1.53 - 3.14 ) | 0.00002067 | 2.25 ( 1.51 - 3.35 ) | 0.00006818 | 0.8398 | 0.8391 | 0.8486 | 1.01 ( 0.83 - 1.21 ) | 0.3902 | 0.9941 (0.8234 - 1.2) | 0.9509 | 1.08 ( 0.93 - 1.25 ) | 0.3328 | 1.16 ( 0.98 - 1.38 ) | 0.07892 |

[a] The positions of SNVs are based on NCBI human genome reference sequence Build 37

[b] A1 is risk-associated allele, and A2 is non-risk-associated allele.

### Appendix Table 7 SKAT of low frequency variants

| Gene | P value | Rs id | Chr | Bp[a] | Allele[b] | Function | MAF[c] |
|---|---|---|---|---|---|---|---|
| *ABCG2* | $1.81 \times 10^{-8}$ | rs9992204 | 4 | 89010495 | T/A | downstream | 0.020442 |
| | | rs528091868 | 4 | 89010860 | G/A | downstream | 0.00166482 |
| | | rs190754327 | 4 | 89011173 | G/C | downstream | 0.00387168 |
| | | rs201142109 | 4 | 89011275 | A/C | downstream | 0.0177976 |
| | | rs201968848 | 4 | 89011308 | G/A | downstream | 0.017165 |
| | | rs1249531481 | 4 | 89012256 | A/G | UTR3 | 0.00110619 |
| | | rs140657468 | 4 | 89012637 | G/A | UTR3 | 0.00331492 |
| | | rs538078746 | 4 | 89014967 | A/G | intronic | 0.00110619 |
| | | rs150610446 | 4 | 89015583 | A/G | intronic | 0.00995575 |
| | | rs147547385 | 4 | 89015762 | C/T | exonic | 0.00221239 |
| | | rs140044879 | 4 | 89016302 | C/A | intronic | 0.00110619 |
| | | rs548254708 | 4 | 89016686 | A/G | exonic | 0.00331858 |
| | | rs563531614 | 4 | 89018067 | C/G | intronic | 0.00221239 |
| | | rs189817431 | 4 | 89018194 | T/A | intronic | 0.00165929 |
| | | rs182470245 | 4 | 89018245 | A/G | intronic | 0.00497788 |
| | | rs34262876 | 4 | 89018540 | C/A | intronic | 0.00497788 |
| | | rs4148158 | 4 | 89018918 | A/G | intronic | 0.013289 |
| | | rs1485618209 | 4 | 89019211 | T/C | intronic | 0.00221239 |
| | | rs45592144 | 4 | 89019615 | T/C | intronic | 0.00331858 |
| | | rs192169063 | 4 | 89020503 | G/A | exonic | 0.0326327 |
| | | rs80169899 | 4 | 89020735 | T/C | intronic | 0.0392699 |
| | | rs1397014591 | 4 | 89021312 | G/A | intronic | 0.00221239 |
| | | rs2622615 | 4 | 89021361 | C/T | intronic | 0.00166852 |
| | | rs183559271 | 4 | 89021497 | T/C | intronic | 0.00387168 |

| | | | | | |
|---|---|---|---|---|---|
| rs527609867 | 4 | 89021570 | A/G | intronic | 0.00110619 |
| rs13147761 | 4 | 89021642 | A/G | intronic | 0.00276549 |
| rs575847850 | 4 | 89021662 | T/A | intronic | 0.00442478 |
| rs375017772 | 4 | 89021905 | A/G | intronic | 0.00110619 |
| rs140341599 | 4 | 89021971 | T/C | intronic | 0.0132743 |
| rs374493295 | 4 | 89022646 | C/T | intronic | 0.00940265 |
| rs1178324544 | 4 | 89022733 | C/T | intronic | 0.00110619 |
| rs1722446606 | 4 | 89023014 | A/G | intronic | 0.00110619 |
| rs138332371 | 4 | 89023131 | T/C | intronic | 0.00608407 |
| rs113822702 | 4 | 89023515 | T/C | intronic | 0.0204646 |
| rs184992816 | 4 | 89023521 | A/G | intronic | 0.00387168 |
| rs201071772 | 4 | 89023762 | A/T | intronic | 0.0155039 |
| rs1297539383 | 4 | 89024707 | C/G | intronic | 0.00110619 |
| rs191577954 | 4 | 89025024 | C/T | intronic | 0.00331492 |
| rs116774624 | 4 | 89025112 | T/C | intronic | 0.00165929 |
| rs183040957 | 4 | 89025527 | G/T | intronic | 0.00221239 |
| rs1358856563 | 4 | 89025864 | T/C | intronic | 0.00221239 |
| rs28665233 | 4 | 89025882 | A/G | intronic | 0.00110619 |
| rs1160870297 | 4 | 89026127 | A/G | intronic | 0.00221239 |
| rs192562676 | 4 | 89026490 | T/C | intronic | 0.00497788 |
| rs192823174 | 4 | 89027297 | C/T | intronic | 0.00110619 |
| rs754440071 | 4 | 89027592 | T/C | intronic | 0.00110619 |
| rs2231149 | 4 | 89028284 | A/G | intronic | 0.00442478 |
| rs770618556 | 4 | 89028403 | T/C | exonic | 0.00110619 |
| rs1264389772 | 4 | 89028436 | A/T | intronic | 0.00276549 |
| rs1246690166 | 4 | 89028437 | A/G | intronic | 0.00276549 |

| | | | | | |
|---|---|---|---|---|---|
| rs764320552 | 4 | 89028438 | A/C | intronic | 0.00276549 |
| rs41282399 | 4 | 89028544 | C/A | intronic | 0.0204646 |
| rs151266026 | 4 | 89028935 | C/T | intronic | 0.00663717 |
| rs183315559 | 4 | 89028979 | A/G | intronic | 0.00110619 |
| rs189214307 | 4 | 89029111 | T/C | intronic | 0.00940265 |
| rs147070185 | 4 | 89029364 | A/G | intronic | 0.00110619 |
| rs1578182576 | 4 | 89029922 | A/G | intronic | 0.00110619 |
| rs528256971 | 4 | 89030004 | A/C | intronic | 0.00663717 |
| rs1160704302 | 4 | 89030357 | A/G | intronic | 0.00165929 |
| rs117076218 | 4 | 89030702 | C/T | intronic | 0.0436947 |
| rs56395445 | 4 | 89031974 | T/A | intronic | 0.023702 |
| rs59525934 | 4 | 89031976 | T/A | intronic | 0.0252525 |
| rs144990343 | 4 | 89032306 | G/A | intronic | 0.00995575 |
| rs147693758 | 4 | 89032451 | C/T | intronic | 0.00663717 |
| rs1396989866 | 4 | 89032466 | G/A | intronic | 0.00110619 |
| rs143244479 | 4 | 89032893 | C/G | intronic | 0.00774336 |
| rs13120400 | 4 | 89033527 | C/T | intronic | 0.00221239 |
| rs45586132 | 4 | 89033661 | C/G | intronic | 0.022677 |
| rs991855488 | 4 | 89033889 | T/A | intronic | 0.00110619 |
| rs142968221 | 4 | 89033964 | A/C | intronic | 0.00165929 |
| rs35622453 | 4 | 89034551 | T/C | exonic | 0.0149502 |
| rs28465652 | 4 | 89034847 | C/G | intronic | 0.0127212 |
| rs746475944 | 4 | 89035106 | G/A | intronic | 0.00110742 |
| rs35427222 | 4 | 89035198 | C/T | intronic | 0.00221239 |
| rs148411002 | 4 | 89035238 | A/C | intronic | 0.00221484 |
| rs116894666 | 4 | 89035264 | A/G | intronic | 0.0149502 |

| | | | | | |
|---|---|---|---|---|---|
| rs201783299 | 4 | 89035512 | T/C | intronic | 0.00276549 |
| rs1030778295 | 4 | 89035624 | T/C | intronic | 0.00165929 |
| rs1377417149 | 4 | 89036494 | T/G | intronic | 0.00110619 |
| rs149805951 | 4 | 89036687 | T/C | intronic | 0.00663717 |
| rs1723517055 | 4 | 89036744 | G/A | intronic | 0.00110619 |
| rs57892861 | 4 | 89037278 | C/T | intronic | 0.0127212 |
| rs56069342 | 4 | 89038140 | A/G | intronic | 0.0315265 |
| rs138982154 | 4 | 89038395 | A/G | intronic | 0.00165929 |
| rs11097181 | 4 | 89038539 | T/C | intronic | 0.0309735 |
| rs1021238825 | 4 | 89038745 | T/C | intronic | 0.00331858 |
| rs1394980880 | 4 | 89039132 | G/A | intronic | 0.00110742 |
| rs34678167 | 4 | 89039297 | A/G | exonic | 0.00497788 |
| rs140207606 | 4 | 89039396 | A/G | exonic | 0.00110619 |
| rs138944374 | 4 | 89039616 | A/G | intronic | 0.00829646 |
| rs182325642 | 4 | 89039911 | G/T | intronic | 0.00110619 |
| rs1012902106 | 4 | 89040266 | A/G | intronic | 0.00110619 |
| rs1287987262 | 4 | 89040683 | C/T | intronic | 0.00166113 |
| rs183311785 | 4 | 89041016 | A/G | intronic | 0.00663717 |
| rs142334415 | 4 | 89041032 | T/C | intronic | 0.0254425 |
| rs1386975278 | 4 | 89041187 | T/G | intronic | 0.00165929 |
| rs567577891 | 4 | 89041400 | A/G | intronic | 0.00165929 |
| rs118030135 | 4 | 89041877 | A/G | intronic | 0.0176991 |
| rs575707882 | 4 | 89042581 | G/C | intronic | 0.00221239 |
| . | 4 | 89042838 | A/G | exonic | 0.00221239 |
| rs767611090 | 4 | 89042888 | C/T | exonic | 0.00221239 |
| rs2231144 | 4 | 89042960 | C/T | intronic | 0.0121681 |

| | | | | | |
|---|---|---|---|---|---|
| rs183829031 | 4 | 89043319 | T/C | intronic | 0.00165929 |
| rs1038905778 | 4 | 89043320 | A/G | intronic | 0.00221239 |
| rs563506137 | 4 | 89043337 | T/C | intronic | 0.00110619 |
| rs575566297 | 4 | 89043461 | T/C | intronic | 0.00276549 |
| rs6814793 | 4 | 89043484 | T/G | intronic | 0.00221239 |
| rs35155973 | 4 | 89043881 | C/G | intronic | 0.00221239 |
| rs546001614 | 4 | 89043883 | G/C | intronic | 0.00110619 |
| rs113752350 | 4 | 89044014 | C/T | intronic | 0.0121816 |
| rs374839750 | 4 | 89044362 | C/T | intronic | 0.00165929 |
| rs541802187 | 4 | 89044699 | T/C | intronic | 0.00110619 |
| rs113119169 | 4 | 89045187 | T/C | intronic | 0.0127212 |
| rs113877129 | 4 | 89045907 | T/C | intronic | 0.00276243 |
| rs114727724 | 4 | 89046089 | T/C | intronic | 0.00331858 |
| rs183504884 | 4 | 89046213 | A/G | intronic | 0.00165929 |
| rs192491356 | 4 | 89046558 | G/A | intronic | 0.00165929 |
| rs7690904 | 4 | 89047100 | A/G | intronic | 0.0116279 |
| rs193034294 | 4 | 89047149 | C/T | intronic | 0.00221729 |
| rs138651211 | 4 | 89047173 | C/A | intronic | 0.0138581 |
| rs181802958 | 4 | 89047823 | A/G | intronic | 0.00497788 |
| rs2725258 | 4 | 89049055 | T/C | intronic | 0.00111111 |
| rs113033839 | 4 | 89049243 | T/C | intronic | 0.0121681 |
| rs188136279 | 4 | 89050083 | T/A | intronic | 0.00442478 |
| rs1207321662 | 4 | 89050360 | G/C | intronic | 0.00165929 |
| rs201150883 | 4 | 89050530 | C/T | intronic | 0.00110619 |
| rs546670609 | 4 | 89051752 | A/G | intronic | 0.00110619 |
| rs1283087860 | 4 | 89052202 | C/T | intronic | 0.00110619 |

| | | | | | |
|---|---|---|---|---|---|
| rs528655917 | 4 | 89052265 | T/C | exonic | 0.00165929 |
| rs372192400 | 4 | 89052305 | A/G | exonic | 0.00221239 |
| rs770985871 | 4 | 89052340 | A/G | exonic | 0.00221239 |
| rs149106245 | 4 | 89052361 | A/T | exonic | 0.00165929 |
| rs75659660 | 4 | 89052569 | C/T | intronic | 0.00221239 |
| rs1724883716 | 4 | 89052799 | G/A | intronic | 0.00110619 |
| rs72552713 | 4 | 89052957 | A/G | exonic | 0.011615 |
| rs766922517 | 4 | 89052997 | T/C | exonic | 0.00221239 |
| rs192023122 | 4 | 89053658 | C/A | intronic | 0.00165929 |
| rs138876482 | 4 | 89053880 | G/A | intronic | 0.028208 |
| rs187177548 | 4 | 89054220 | A/G | intronic | 0.00110619 |
| rs117319230 | 4 | 89054285 | C/T | intronic | 0.00331858 |
| rs45488400 | 4 | 89054332 | C/T | intronic | 0.0121681 |
| rs17013859 | 4 | 89054486 | T/C | intronic | 0.0121681 |
| rs45555139 | 4 | 89054512 | A/G | intronic | 0.00110619 |
| rs538750835 | 4 | 89054620 | C/G | intronic | 0.00110619 |
| rs193120963 | 4 | 89054892 | A/G | intronic | 0.00940265 |
| rs72554043 | 4 | 89055016 | A/G | intronic | 0.0121681 |
| rs550182016 | 4 | 89055067 | C/T | intronic | 0.00221239 |
| rs191608907 | 4 | 89055263 | C/A | intronic | 0.0110619 |
| rs17731538 | 4 | 89055379 | A/G | intronic | 0.0414823 |
| rs1560699933 | 4 | 89055493 | A/G | intronic | 0.00221239 |
| rs17013870 | 4 | 89056108 | C/T | intronic | 0.0121681 |
| rs72875335 | 4 | 89056187 | A/G | intronic | 0.0121681 |
| rs4148154 | 4 | 89056427 | G/T | intronic | 0.0105088 |
| rs377284700 | 4 | 89056581 | G/T | intronic | 0.00110619 |

| | | | | | |
|---|---|---|---|---|---|
| . | 4 | 89056679 | C/A | intronic | 0.00276549 |
| rs1364852630 | 4 | 89056927 | G/A | intronic | 0.00110742 |
| rs985912657 | 4 | 89057644 | A/G | intronic | 0.00110619 |
| rs113737399 | 4 | 89057843 | A/G | intronic | 0.0121681 |
| rs76351890 | 4 | 89057870 | A/T | intronic | 0.0243363 |
| rs190163000 | 4 | 89057940 | T/G | intronic | 0.00276855 |
| rs568973129 | 4 | 89058010 | C/T | intronic | 0.00940265 |
| rs1423301106 | 4 | 89058215 | A/G | intronic | 0.0144444 |
| rs1163503712 | 4 | 89058219 | A/G | intronic | 0.00332594 |
| rs116914853 | 4 | 89058580 | G/A | intronic | 0.0237832 |
| rs145433166 | 4 | 89059054 | T/C | intronic | 0.00497788 |
| rs2869732 | 4 | 89059087 | G/A | intronic | 0.00221239 |
| rs527465995 | 4 | 89059155 | G/T | intronic | 0.00442478 |
| rs180724363 | 4 | 89060280 | A/G | intronic | 0.00609081 |
| rs185165642 | 4 | 89060397 | G/C | intronic | 0.00110619 |
| rs143730946 | 4 | 89060409 | A/C | intronic | 0.00829646 |
| rs147505061 | 4 | 89061296 | A/G | intronic | 0.00276549 |
| rs140134788 | 4 | 89061303 | T/C | intronic | 0.00774336 |
| rs973977208 | 4 | 89061373 | C/T | intronic | 0.00387168 |
| rs13130891 | 4 | 89062075 | G/A | intronic | 0.0143805 |
| rs13137622 | 4 | 89062513 | T/G | intronic | 0.0143805 |
| rs776265185 | 4 | 89062691 | G/T | intronic | 0.00110619 |
| rs893637148 | 4 | 89062706 | A/G | intronic | 0.00110619 |
| rs186721762 | 4 | 89063042 | G/A | intronic | 0.00553097 |
| rs2046134 | 4 | 89063363 | A/G | intronic | 0.0121681 |
| rs1042167928 | 4 | 89063504 | T/C | intronic | 0.00331858 |

| | | | | | |
|---|---|---|---|---|---|
| rs149129138 | 4 | 89064125 | C/T | intronic | 0.00940265 |
| . | 4 | 89064216 | A/C | intronic | 0.00221239 |
| rs28742177 | 4 | 89065017 | G/T | intronic | 0.00387168 |
| rs190605325 | 4 | 89065748 | T/C | intronic | 0.00221729 |
| rs12641988 | 4 | 89065861 | A/G | intronic | 0.00221484 |
| rs1220874687 | 4 | 89066164 | T/C | intronic | 0.00165929 |
| rs192958180 | 4 | 89066335 | C/A | intronic | 0.00165929 |
| rs184097945 | 4 | 89066337 | A/G | intronic | 0.00165929 |
| rs180732996 | 4 | 89067257 | C/T | intronic | 0.00165929 |
| rs555129847 | 4 | 89067345 | A/G | intronic | 0.00110619 |
| rs913089205 | 4 | 89067444 | C/T | intronic | 0.00110619 |
| rs1416405714 | 4 | 89069526 | A/G | intronic | 0.00221239 |
| rs186311470 | 4 | 89069531 | A/G | intronic | 0.00165929 |
| rs558803064 | 4 | 89069891 | A/G | intronic | 0.00276549 |
| rs552548891 | 4 | 89070286 | C/G | intronic | 0.00387168 |
| rs553545824 | 4 | 89070655 | C/T | intronic | 0.00221239 |
| rs1169652432 | 4 | 89070751 | C/G | intronic | 0.00110619 |
| rs367899264 | 4 | 89071565 | C/G | intronic | 0.00442478 |
| rs151046377 | 4 | 89071971 | G/A | intronic | 0.00331858 |
| rs114916387 | 4 | 89073289 | C/T | intronic | 0.00165929 |
| rs565974346 | 4 | 89074118 | C/A | intronic | 0.00442478 |
| rs547230132 | 4 | 89075759 | A/C | intronic | 0.00110619 |
| rs540103507 | 4 | 89075854 | A/G | intronic | 0.00110742 |
| rs796397692 | 4 | 89076452 | A/G | intronic | 0.00221484 |
| rs138215164 | 4 | 89076778 | T/C | intronic | 0.0138274 |
| rs533391484 | 4 | 89076917 | T/G | intronic | 0.00165929 |

| | | | | | |
|---|---|---|---|---|---|
| rs200697172 | 4 | 89077060 | G/A | intronic | 0.00110619 |
| rs1490810748 | 4 | 89077269 | A/G | intronic | 0.00110619 |
| rs187325658 | 4 | 89078017 | T/G | intronic | 0.0315265 |
| rs184518520 | 4 | 89078298 | C/T | intronic | 0.00221239 |
| rs189370293 | 4 | 89078616 | G/A | intronic | 0.00110619 |
| rs1397248107 | 4 | 89078804 | T/C | intronic | 0.00110619 |
| rs907561164 | 4 | 89079316 | A/T | intronic | 0.00553097 |
| rs1201964734 | 4 | 89079629 | A/G | UTR5 | 0.00165929 |
| rs111692024 | 4 | 89079647 | A/G | UTR5 | 0.00165929 |
| rs2231136 | 4 | 89079780 | A/G | UTR5 | 0.0188053 |
| rs76656413 | 4 | 89080180 | A/G | intronic | 0.00884956 |
| rs962452118 | 4 | 89081770 | A/G | intronic | 0.00995575 |
| rs1305320082 | 4 | 89082195 | T/G | intronic | 0.00110742 |
| rs185832097 | 4 | 89083335 | T/C | intronic | 0.00110619 |
| rs562612257 | 4 | 89083414 | C/T | intronic | 0.00608407 |
| rs181511040 | 4 | 89084189 | A/G | intronic | 0.00387168 |
| rs537724512 | 4 | 89084623 | A/G | intronic | 0.00387168 |
| rs182058119 | 4 | 89084730 | C/A | intronic | 0.00442478 |
| rs544748057 | 4 | 89085006 | C/T | intronic | 0.00110619 |
| rs143524170 | 4 | 89085203 | A/G | intronic | 0.0132743 |
| rs138944612 | 4 | 89085580 | A/G | intronic | 0.00110619 |
| rs183867894 | 4 | 89086013 | C/T | intronic | 0.00110619 |
| rs77778163 | 4 | 89087079 | C/T | intronic | 0.022677 |
| rs376940569 | 4 | 89088651 | A/G | intronic | 0.00110619 |
| rs550190578 | 4 | 89088754 | A/T | intronic | 0.00110619 |
| rs1236617138 | 4 | 89088872 | G/C | intronic | 0.00221239 |

| | | | | | |
|---|---|---|---|---|---|
| rs377598902 | 4 | 89089997 | C/T | intronic | 0.00387597 |
| rs980398905 | 4 | 89090031 | A/T | intronic | 0.00221484 |
| rs142890537 | 4 | 89090415 | G/C | intronic | 0.0370575 |
| rs183713158 | 4 | 89090968 | C/A | intronic | 0.00110619 |
| rs147307831 | 4 | 89091728 | A/G | intronic | 0.0121681 |
| rs1227416276 | 4 | 89092106 | T/G | intronic | 0.00165929 |
| rs1338993728 | 4 | 89093448 | C/A | intronic | 0.00112994 |
| rs537059270 | 4 | 89093471 | G/T | intronic | 0.00110619 |
| rs1243682358 | 4 | 89093582 | T/G | intronic | 0.00110619 |
| rs1210648888 | 4 | 89093728 | G/T | intronic | 0.00227531 |
| rs935373446 | 4 | 89093811 | C/G | intronic | 0.00110619 |
| rs144123347 | 4 | 89095272 | T/G | intronic | 0.00276549 |
| rs75463711 | 4 | 89095333 | A/T | intronic | 0.0221484 |
| rs1303239005 | 4 | 89095445 | A/G | intronic | 0.00165929 |
| rs576043906 | 4 | 89096349 | C/T | intronic | 0.00884956 |
| rs942365429 | 4 | 89096367 | T/C | intronic | 0.00165929 |
| rs13111149 | 4 | 89097819 | G/T | intronic | 0.00228311 |
| rs188633373 | 4 | 89097879 | G/C | intronic | 0.00387597 |
| rs531545314 | 4 | 89097996 | C/T | intronic | 0.00165929 |
| rs189359538 | 4 | 89098425 | C/G | intronic | 0.00608407 |
| rs1241531923 | 4 | 89098813 | T/A | intronic | 0.00110619 |
| rs954950536 | 4 | 89098932 | G/A | intronic | 0.00165929 |
| rs7672821 | 4 | 89099150 | A/G | intronic | 0.0165929 |
| rs1297452206 | 4 | 89100092 | C/T | intronic | 0.00110619 |
| rs62310625 | 4 | 89100336 | T/C | intronic | 0.0127212 |
| rs201076764 | 4 | 89101467 | A/T | intronic | 0.00165929 |

| | | | | | |
|---|---|---|---|---|---|
| rs118159145 | 4 | 89102418 | C/G | intronic | 0.0199115 |
| rs1276948990 | 4 | 89102469 | A/T | intronic | 0.00110619 |
| rs111800277 | 4 | 89102744 | A/G | intronic | 0.00110619 |
| rs34569524 | 4 | 89102785 | G/A | intronic | 0.00332226 |
| rs547117924 | 4 | 89103245 | G/A | intronic | 0.00442478 |
| rs1230141221 | 4 | 89103383 | C/T | intronic | 0.00110619 |
| rs534089006 | 4 | 89103784 | G/A | intronic | 0.00221484 |
| rs6815644 | 4 | 89104065 | G/A | intronic | 0.00665188 |
| rs112804164 | 4 | 89105799 | C/T | intronic | 0.00110619 |
| rs1580353 | 4 | 89105916 | T/C | intronic | 0.00387168 |
| rs368973431 | 4 | 89106126 | T/G | intronic | 0.00165929 |
| rs35677388 | 4 | 89106377 | A/G | intronic | 0.00387168 |
| rs77047724 | 4 | 89106744 | C/T | intronic | 0.00110619 |
| . | 4 | 89107639 | C/T | intronic | 0.00110619 |
| rs529847433 | 4 | 89108539 | T/C | intronic | 0.00166113 |
| rs13147650 | 4 | 89109396 | A/G | intronic | 0.00387168 |
| rs180777251 | 4 | 89109495 | C/G | intronic | 0.00553097 |
| rs76014515 | 4 | 89109510 | T/C | intronic | 0.00110619 |
| rs56892189 | 4 | 89109643 | A/G | intronic | 0.00387168 |
| rs144762183 | 4 | 89109736 | A/G | intronic | 0.00166113 |
| rs184998355 | 4 | 89109827 | T/C | intronic | 0.0132743 |
| rs34160406 | 4 | 89109960 | A/G | intronic | 0.00387168 |
| rs2127862 | 4 | 89110628 | T/C | intronic | 0.00442968 |
| . | 4 | 89110662 | A/C | intronic | 0.00110742 |
| rs113239856 | 4 | 89110708 | G/A | intronic | 0.00110619 |
| rs75889116 | 4 | 89110859 | C/T | intronic | 0.0199115 |

| | | | | | |
|---|---|---|---|---|---|
| rs79496099 | 4 | 89111104 | A/G | intronic | 0.00110619 |
| rs931256121 | 4 | 89111159 | A/T | intronic | 0.00276549 |
| rs75075977 | 4 | 89111947 | G/A | intronic | 0.00110619 |
| rs113972065 | 4 | 89112240 | C/A | intronic | 0.00387168 |
| rs111937630 | 4 | 89112342 | T/C | intronic | 0.00110742 |
| rs371725863 | 4 | 89112360 | T/C | intronic | 0.00221484 |
| rs13117650 | 4 | 89112460 | A/G | intronic | 0.00442478 |
| rs79335697 | 4 | 89112563 | A/T | intronic | 0.00719823 |
| rs6854688 | 4 | 89112833 | G/A | intronic | 0.00387597 |
| rs11097182 | 4 | 89112908 | C/T | intronic | 0.00553097 |
| rs976860655 | 4 | 89113036 | T/C | intronic | 0.00110619 |
| rs574033331 | 4 | 89113199 | T/C | intronic | 0.00221239 |
| rs13152371 | 4 | 89113497 | C/T | intronic | 0.00497788 |
| rs180872136 | 4 | 89113760 | T/C | intronic | 0.00276549 |
| rs112691981 | 4 | 89113855 | A/G | intronic | 0.00110619 |
| rs4693928 | 4 | 89113887 | G/A | intronic | 0.00498891 |
| rs527774222 | 4 | 89114756 | G/A | intronic | 0.00110619 |
| rs1013896737 | 4 | 89115744 | G/T | intronic | 0.00165929 |
| rs1225894466 | 4 | 89116528 | T/G | intronic | 0.00110619 |
| rs1256721860 | 4 | 89116725 | T/C | intronic | 0.00110619 |
| rs9993821 | 4 | 89116978 | A/T | intronic | 0.00497788 |
| rs1258430837 | 4 | 89116981 | A/G | intronic | 0.00110619 |
| rs191157291 | 4 | 89117109 | C/T | intronic | 0.00110619 |
| rs564578785 | 4 | 89117196 | A/G | intronic | 0.00110619 |
| rs1055944586 | 4 | 89117349 | C/T | intronic | 0.00110619 |
| rs35304142 | 4 | 89118445 | G/A | intronic | 0.00221239 |

| | | | | | |
|---|---|---|---|---|---|
| rs11097183 | 4 | 89118545 | G/A | intronic | 0.00110619 |
| rs13123636 | 4 | 89118799 | T/A | intronic | 0.00221239 |
| rs2869733 | 4 | 89119201 | G/A | intronic | 0.00111359 |
| rs12513247 | 4 | 89119258 | T/C | intronic | 0.00331858 |
| rs1156252853 | 4 | 89119382 | A/G | intronic | 0.00110619 |
| rs10856870 | 4 | 89119659 | T/C | intronic | 0.00276855 |
| rs561939396 | 4 | 89119791 | A/G | intronic | 0.00276549 |
| rs7661664 | 4 | 89120147 | G/A | intronic | 0.00165929 |
| rs1307642153 | 4 | 89120181 | T/C | intronic | 0.00165929 |
| rs1382731772 | 4 | 89121252 | G/A | intronic | 0.00165929 |
| rs146883245 | 4 | 89121307 | A/T | intronic | 0.0138274 |
| rs9968365 | 4 | 89121464 | C/A | intronic | 0.00276549 |
| rs545389520 | 4 | 89121785 | T/C | intronic | 0.00996678 |
| rs182069549 | 4 | 89121929 | A/G | intronic | 0.00221239 |
| rs1342066900 | 4 | 89122472 | T/C | intronic | 0.00165929 |
| rs1268644207 | 4 | 89122551 | G/A | intronic | 0.00165929 |
| rs192199098 | 4 | 89122911 | T/C | intronic | 0.00110619 |
| rs147000542 | 4 | 89123337 | G/A | intronic | 0.0166113 |
| rs6845663 | 4 | 89123748 | T/G | intronic | 0.00221484 |
| rs574180936 | 4 | 89125097 | T/C | intronic | 0.00221239 |
| rs28401615 | 4 | 89125779 | C/T | intronic | 0.00221239 |
| rs939191723 | 4 | 89125990 | T/G | intronic | 0.00276549 |
| rs116942732 | 4 | 89126071 | A/G | intronic | 0.0337389 |
| rs183744794 | 4 | 89126083 | G/C | intronic | 0.00276549 |
| rs1157176940 | 4 | 89126397 | C/T | intronic | 0.00165929 |
| rs12512071 | 4 | 89126973 | C/T | intronic | 0.00387168 |

| | | | | | |
|---|---|---|---|---|---|
| rs1253923985 | 4 | 89127338 | T/C | intronic | 0.00110619 |
| rs13149967 | 4 | 89127339 | A/G | intronic | 0.00221239 |
| rs4693931 | 4 | 89127785 | A/T | intronic | 0.00221239 |
| rs187230470 | 4 | 89128075 | A/C | intronic | 0.00221239 |
| rs79309290 | 4 | 89128325 | A/G | intronic | 0.017146 |
| rs59492220 | 4 | 89128435 | A/G | intronic | 0.00221239 |
| rs61055940 | 4 | 89128736 | C/T | intronic | 0.00221239 |
| rs1000039382 | 4 | 89128916 | T/C | intronic | 0.00387168 |
| rs184857710 | 4 | 89129464 | C/G | intronic | 0.00221239 |
| rs11723264 | 4 | 89129484 | G/A | intronic | 0.00221239 |
| rs74624975 | 4 | 89129487 | T/G | intronic | 0.00497788 |
| rs1194572760 | 4 | 89129517 | T/C | intronic | 0.00165929 |
| . | 4 | 89129847 | T/C | intronic | 0.00110619 |
| rs543100299 | 4 | 89129939 | A/G | intronic | 0.00165929 |
| rs1578279587 | 4 | 89129945 | A/C | intronic | 0.00110619 |
| rs535196401 | 4 | 89130131 | A/G | intronic | 0.00110619 |
| rs547971297 | 4 | 89130140 | T/C | intronic | 0.00166113 |
| rs1219909911 | 4 | 89130543 | T/C | intronic | 0.00110988 |
| rs1055865400 | 4 | 89130749 | T/C | intronic | 0.00110619 |
| rs972184785 | 4 | 89130987 | A/G | intronic | 0.00110619 |
| . | 4 | 89131045 | C/A | intronic | 0.00110619 |
| rs537691296 | 4 | 89131505 | G/A | intronic | 0.00220994 |
| rs1445828853 | 4 | 89131857 | G/A | intronic | 0.00110619 |
| rs1457063507 | 4 | 89131910 | G/A | intronic | 0.00110619 |
| rs6842010 | 4 | 89132265 | A/T | intronic | 0.00387168 |
| rs568387623 | 4 | 89133386 | C/G | intronic | 0.00110619 |

| | | | | | |
|---|---|---|---|---|---|
| rs1484892984 | 4 | 89134011 | T/G | intronic | 0.00165929 |
| rs561076689 | 4 | 89135176 | A/G | intronic | 0.00165929 |
| rs201981466 | 4 | 89135506 | G/C | intronic | 0.00110619 |
| rs183072332 | 4 | 89135641 | T/G | intronic | 0.0127212 |
| rs139195406 | 4 | 89136137 | T/G | intronic | 0.0110619 |
| rs141017431 | 4 | 89136246 | C/A | intronic | 0.00110619 |
| rs192433545 | 4 | 89136248 | T/A | intronic | 0.0188053 |
| rs192343384 | 4 | 89136963 | C/T | intronic | 0.00663717 |
| rs4482728 | 4 | 89138075 | T/C | intronic | 0.000553097 |
| rs1030994651 | 4 | 89138197 | A/T | intronic | 0.00497788 |
| rs182653740 | 4 | 89138238 | A/T | intronic | 0.00110619 |
| rs62309287 | 4 | 89138761 | A/C | intronic | 0.00221239 |
| rs60473789 | 4 | 89139219 | T/C | intronic | 0.00110619 |
| . | 4 | 89139516 | G/T | intronic | 0.00110619 |
| rs190696974 | 4 | 89139975 | A/C | intronic | 0.00165929 |
| rs183881719 | 4 | 89140010 | C/T | intronic | 0.00110619 |
| rs367797920 | 4 | 89140379 | C/T | intronic | 0.00110742 |
| rs1374463771 | 4 | 89141223 | G/T | intronic | 0.00165929 |
| rs542581079 | 4 | 89141736 | G/C | intronic | 0.00165929 |
| rs982332474 | 4 | 89141988 | A/C | intronic | 0.00165929 |
| rs1186645537 | 4 | 89142014 | C/T | intronic | 0.00110619 |
| rs116251394 | 4 | 89142440 | C/T | intronic | 0.00165929 |
| rs185543973 | 4 | 89143027 | G/A | intronic | 0.00331492 |
| rs554555755 | 4 | 89143161 | T/G | intronic | 0.00110619 |
| rs959900052 | 4 | 89143261 | T/C | intronic | 0.00110619 |
| rs6811890 | 4 | 89143340 | T/A | intronic | 0.0376106 |

| | | | | | |
|---|---|---|---|---|---|
| rs554775093 | 4 | 89143788 | T/G | intronic | 0.00110619 |
| rs148458422 | 4 | 89144542 | C/T | intronic | 0.00608407 |
| rs563485094 | 4 | 89144822 | A/G | intronic | 0.00110619 |
| rs117961005 | 4 | 89144857 | G/A | intronic | 0.00165929 |
| rs141678006 | 4 | 89145405 | C/G | intronic | 0.0237832 |
| rs752588009 | 4 | 89145778 | A/G | intronic | 0.00165929 |
| rs117294932 | 4 | 89146112 | C/T | intronic | 0.00719027 |
| rs1039887424 | 4 | 89146516 | A/C | intronic | 0.00110619 |
| rs1337157406 | 4 | 89146783 | T/G | intronic | 0.00110742 |
| rs1033518166 | 4 | 89147163 | A/C | intronic | 0.00110619 |
| rs2127864 | 4 | 89147993 | G/A | intronic | 0.0381637 |
| rs139968558 | 4 | 89148182 | T/G | intronic | 0.00110619 |
| rs118136756 | 4 | 89148434 | C/A | intronic | 0.00940265 |
| rs9985959 | 4 | 89148785 | T/C | intronic | 0.00165929 |
| rs7655059 | 4 | 89149144 | G/C | intronic | 0.0381637 |
| rs185089711 | 4 | 89149472 | A/G | intronic | 0.00110619 |
| rs35892152 | 4 | 89149626 | T/C | intronic | 0.00165929 |
| rs549356839 | 4 | 89150135 | A/C | intronic | 0.00719027 |
| rs62308057 | 4 | 89150178 | G/C | intronic | 0.0176991 |
| rs77197787 | 4 | 89150387 | T/C | intronic | 0.0176991 |
| rs1257657271 | 4 | 89150557 | A/G | intronic | 0.00110619 |
| rs149251214 | 4 | 89150931 | A/G | intronic | 0.00110619 |
| rs148056965 | 4 | 89151026 | G/A | intronic | 0.00829646 |
| rs536957456 | 4 | 89151382 | A/T | intronic | 0.00221239 |
| rs11736552 | 4 | 89151397 | G/A | intronic | 0.02051 |
| rs553612098 | 4 | 89151533 | T/C | intronic | 0.00221239 |

| | | | | | | | |
|---|---|---|---|---|---|---|---|
| | | rs183648317 | 4 | 89151928 | C/A | intronic | 0.00719027 |
| | | rs1454402012 | 4 | 89152272 | G/A | UTR5 | 0.00276549 |
| | | rs182367277 | 4 | 89152386 | A/G | UTR5 | 0.00940265 |
| | | rs186907101 | 4 | 89152391 | C/T | UTR5 | 0.00829646 |
| | | rs62309960 | 4 | 89152616 | C/G | UTR5 | 0.00221239 |
| | | rs36111742 | 4 | 89152718 | G/A | UTR5 | 0.0176991 |
| | | rs1254091077 | 4 | 89152741 | A/G | UTR5 | 0.00110619 |
| | | rs557353856 | 11 | 64357589 | C/T | upstream | 0.00331858 |
| | | rs2038880946 | 11 | 64357762 | A/G | upstream | 0.00110619 |
| | | rs1294924027 | 11 | 64357918 | A/G | upstream | 0.00276549 |
| | | rs2038888655 | 11 | 64358048 | C/G | upstream | 0.00110742 |
| | | rs67048806 | 11 | 64358100 | A/G | upstream | 0.00110619 |
| | | rs892916775 | 11 | 64358430 | A/C | UTR5 | 0.00110742 |
| | | rs886048451 | 11 | 64358936 | T/C | UTR5 | 0.00110619 |
| | | rs369153816 | 11 | 64358954 | T/C | UTR5 | 0.00221484 |
| | | rs201365068 | 11 | 64358984 | G/C | UTR5 | 0.00276855 |
| *SLC22A12* | $1.06 \times 10^{-8}$ | rs121907896 | 11 | 64359297 | A/G | exonic | 0.00221729 |
| | | rs549386066 | 11 | 64359643 | T/C | intronic | 0.00110742 |
| | | rs1013199344 | 11 | 64359649 | A/G | intronic | 0.00332226 |
| | | rs187404454 | 11 | 64359693 | T/C | intronic | 0.00110742 |
| | | rs1012855300 | 11 | 64359948 | A/G | intronic | 0.00110742 |
| | | rs76660248 | 11 | 64359994 | A/C | intronic | 0.0199336 |
| | | rs1352610654 | 11 | 64360142 | G/C | intronic | 0.00166297 |
| | | rs371761990 | 11 | 64360724 | T/C | intronic | 0.00442968 |
| | | rs75786299 | 11 | 64361042 | A/G | intronic | 0.0354767 |
| | | rs201136391 | 11 | 64361124 | A/G | exonic | 0.00110742 |

| | | | | | |
|---|---|---|---|---|---|
| rs79933955 | 11 | 64361869 | T/C | intronic | 0.00165929 |
| rs376000964 | 11 | 64362118 | C/G | intronic | 0.00288351 |
| rs12363578 | 11 | 64364866 | T/C | intronic | 0.0194013 |
| rs76945158 | 11 | 64364959 | T/C | intronic | 0.0454042 |
| rs2022048 | 11 | 64365167 | T/C | intronic | 0.0481195 |
| rs1043069143 | 11 | 64365246 | T/C | intronic | 0.00110742 |
| rs1396406487 | 11 | 64365247 | A/G | intronic | 0.00221484 |
| rs373058278 | 11 | 64365251 | T/C | intronic | 0.00166113 |
| rs78160447 | 11 | 64365256 | T/C | intronic | 0.00775194 |
| rs752017508 | 11 | 64366007 | T/C | exonic | 0.00110865 |
| rs144128502 | 11 | 64366451 | T/C | intronic | 0.00110742 |
| rs78880801 | 11 | 64366461 | T/C | intronic | 0.00110742 |
| rs541637819 | 11 | 64366537 | T/C | intronic | 0.00277162 |
| rs1371838063 | 11 | 64366547 | T/C | intronic | 0.00110619 |
| rs117045876 | 11 | 64366608 | C/A | intronic | 0.0248894 |
| rs1297148198 | 11 | 64366664 | T/C | intronic | 0.00110619 |
| rs536803678 | 11 | 64366835 | A/G | intronic | 0.00166113 |
| rs543512307 | 11 | 64367272 | T/C | exonic | 0.00443459 |
| rs557630188 | 11 | 64368131 | T/C | intronic | 0.00110742 |
| rs200072517 | 11 | 64368288 | C/T | exonic | 0.00276855 |
| rs182636416 | 11 | 64368562 | A/G | intronic | 0.00497788 |
| rs117280820 | 11 | 64368664 | A/C | intronic | 0.00277162 |
| rs114845820 | 11 | 64368943 | A/G | intronic | 0.00608407 |
| rs148845071 | 11 | 64369291 | A/G | UTR3 | 0.0254425 |
| . | 11 | 64369450 | G/A | UTR3 | 0.00110497 |
| rs1184307204 | 11 | 64369756 | T/C | UTR3 | 0.00110619 |

| | | | | | |
|---|---|---|---|---|---|
| rs1166925277 | 11 | 64370114 | G/C | downstream | 0.00221484 |
| rs1461763824 | 11 | 64370251 | A/G | downstream | 0.00110742 |
| rs966977560 | 11 | 64370450 | A/G | downstream | 0.00277162 |
| rs916556221 | 11 | 64370671 | T/C | downstream | 0.00165929 |

[a] The positions of SNVs are based on NCBI human genome reference sequence Build 37

[b] Minor allele/ Major allele

[c] Minor allele frequencies

### Appendix Table 8 Uncommon SNVs and indels in gout cases

| Rs id | Chr | Bp[a] | Number of individuals | Frequency of gout | P value | Ref | Alt | Gene | ExonicFunction | AAChange | Locus | ExAC_ALL | ExAC_EAS |
|---|---|---|---|---|---|---|---|---|---|---|---|---|---|
| rs370653261 | 3 | 185252641 | 4 | 0.0071429 | 0.009 | C | A | *LIPH* | nonsynonymous SNV | R110L | 3q27.2 | 6.59E-05 | 0 |
| rs753112355 | 6 | 51889432 | 4 | 0.0071429 | 0.009 | C | A | *PKHD1* | nonsynonymous SNV | A1726S | 6p12.2 | 3.31E-05 | 0.0009 |
| rs753523115 | 16 | 56914054 | 4 | 0.0071429 | 0.009 | G | A | *SLC12A3* | nonsynonymous SNV | D486N | 16q13 | 1.33E-05 | 0 |
| rs200233661 | 1 | 162725022 | 3 | 0.0053571 | 0.0294 | G | A | *DDR2* | nonsynonymous SNV | R165Q | 1q23.3 | 4.12E-05 | . |
| rs564123317 | 1 | 201286739 | 3 | 0.0053571 | 0.0294 | C | T | *PKP1* | nonsynonymous SNV | L296F | 1q32.1 | 8.40E-06 | . |
| rs1006417970 | 16 | 30485579 | 3 | 0.0053571 | 0.0294 | G | C | *ITGAL* | nonsynonymous SNV | G42R | 16p11.2 | . | 0.0005 |
| rs1575982996 | 3 | 38781059 | 3 | 0.0053571 | 0.0294 | C | T | *SCN10A* | nonsynonymous SNV | E743K | 3p22.2 | . | 0.0002 |

| | | | | | | | | | | | | | |
|---|---|---|---|---|---|---|---|---|---|---|---|---|---|
| rs760082663 | 1 | 85498411 | 3 | 0.0053571 | 0.0294 | C | G | *MCOLN3* | nonsynonymous SNV | E234Q | 1p22.3 | . | . |
| . | 1 | 231299484 | 3 | 0.0053571 | 0.0294 | C | CCGA | *TRIM67* | nonframeshift substitution | P257_P258insT | 1q42.2 | . | . |
| . | 2 | 200820543 | 3 | 0.0053571 | 0.0294 | C | CT | *MAIP1* | frameshift substitution | p.P9Tfs*50 | 2q33.1 | . | . |
| rs755807204 | 16 | 30021353 | 3 | 0.0053571 | 0.0294 | G | GGGGGGGCCAGAGCCA | *DOC2A* | nonframeshift substitution | P63_P64insLALAP | 16p11.2 | 1.75E-05 | 0.0002 |

[a] The positions of SNVs and indels are based on NCBI human genome reference sequence Build 37

### Appendix Table 9 Significantly different allele counts of uncommon SNVs located in *RCOR1*

| Rs id | Chr | Bp[a] | Ref | Alt | Function | Gene | Frequency of Alt Allele | | | G-NG | | G-H | |
|---|---|---|---|---|---|---|---|---|---|---|---|---|---|
| | | | | | | | Gout | HUA | Control | P-value | OR (95% CI) | P-value | OR (95% CI) |
| rs1595175942 | 14 | 103028071 | C | G | intergenic | *LINC02323-RCOR1* | 3 (0.54%) | 0 (0.00%) | 0 (0.00%) | 0.03 | Inf | 0.28 | Inf |
| rs115286323 | 14 | 103047948 | C | T | intergenic | *LINC02323-RCOR1* | 26 (4.64%) | 12 (3.14%) | 22 (2.54%) | 4.58E-02 | 1.74 (0.99-3.02) | 0.31 | 1.50 (0.72-3.31) |
| rs923806140 | 14 | 103050657 | C | T | intergenic | *LINC02323-RCOR1* | 3 (0.54%) | 0 (0.00%) | 0 (0.00%) | 0.03 | Inf | 0.28 | Inf |
| . | 14 | 103109792 | T | G | intronic | *RCOR1* | 3 (0.54%) | 0 (0.00%) | 0 (0.00%) | 0.03 | Inf | 0.28 | Inf |
| rs1290583982 | 14 | 103117176 | T | G | intronic | *RCOR1* | 3 (0.54%) | 0 (0.00%) | 0 (0.00%) | 0.03 | Inf | 0.28 | Inf |
| rs746410370 | 14 | 103128943 | G | A | intronic | *RCOR1* | 4 (0.71%) | 0 (0.00%) | 0 (0.00%) | 0.01 | Inf | 0.15 | Inf |
| rs77933057 | 14 | 103190183 | G | A | intronic | *RCOR1* | 36 (6.43%) | 19 (4.97%) | 27 (3.12%) | 0.01 | 1.79 (1.11-2.87) | 0.40 | 1.31 (0.72-2.46) |
| rs569723412 | 14 | 103203627 | G | A | intergenic | *RCOR1-TRAF3* | 3 (0.54%) | 0 (0.00%) | 0 (0.00%) | 0.03 | Inf | 0.28 | Inf |
| rs114476419 | 14 | 103211176 | T | A | intergenic | *RCOR1-TRAF3* | 3 (0.54%) | 0 (0.00%) | 0 (0.00%) | 0.03 | Inf | 0.28 | Inf |
| rs572953510 | 14 | 103211544 | G | A | intergenic | *RCOR1-TRAF3* | 3 (0.54%) | 0 (0.00%) | 0 (0.00%) | 0.03 | Inf | 0.28 | Inf |
| rs188486929 | 14 | 103123362 | T | C | intronic | *RCOR1* | 11 (1.96%) | 1 (0.26%) | 11 (1.27%) | 0.11 | 2.06 (0.82-5.14) | 0.03 | 7.62 (1.10-328.97) |

| | | | | | | | | | | | | | |
|---|---|---|---|---|---|---|---|---|---|---|---|---|---|
| rs575695277 | 14 | 103138196 | C | T | intronic | *RCOR1* | 7 (1.25%) | 0 (0.00%) | 6 (0.69%) | 0.13 | 2.62 (0.75-9.48) | 4.58E-02 | Inf |
| rs552519074 | 14 | 103157992 | A | G | intronic | *RCOR1* | 7 (1.25%) | 0 (0.00%) | 6 (0.69%) | 0.13 | 2.62 (0.75-9.48) | 4.58E-02 | Inf |
| rs181707401 | 14 | 103215090 | T | G | intergenic | *RCOR1-TRAF3* | 0 (0.00%) | 4 (1.05%) | 4 (0.46%) | 0.06 | 0.00 (0.00-1.30) | 0.03 | 0.00 (0.00-1.02) |

[a] The positions of SNVs are based on NCBI human genome reference sequence Build 37

### Appendix Table 10 Significantly different counts of indels located in *RCOR1*

| Rs id | Chr | Bp[a] | Ref | Alt | Type | Function | Gene | Frequency of Alt Allele | | | G-NG | | G-H | |
|---|---|---|---|---|---|---|---|---|---|---|---|---|---|---|
| | | | | | | | | Gout | HUA | Control | P-value | OR (95% CI) | P-value | OR (95% CI) |
| rs36047872 | 14 | 103034540 | A | AAAG | DELINS | intergenic | *LINC02323;RCOR1* | 322 (57.50%) | 181 (47.38%) | 445 (51.39%) | 4.33E-03 | 1.34 (1.09-1.65) | 2.74E-03 | 1.50 (1.15-1.97) |
| rs368679111 | 14 | 103044706 | A | ACATTCC | INS | intergenic | *LINC02323;RCOR1* | 295 (52.68%) | 169 (44.24%) | 406 (46.99%) | 0.01 | 1.30 (1.06-1.59) | 0.01 | 1.40 (1.07-1.84) |
| rs371639695 | 14 | 103044709 | G | GA | INS | intergenic | *LINC02323;RCOR1* | 295 (52.68%) | 169 (44.24%) | 406 (46.99%) | 0.01 | 1.30 (1.06-1.59) | 0.01 | 1.40 (1.07-1.84) |
| rs143296299 | 14 | 103067613 | G | GCCTCCT | INS | intronic | *RCOR1* | 51 (9.11%) | 24 (6.28%) | 40 (4.62%) | 2.35E-03 | 1.85 (1.24-2.76) | 0.14 | 1.49 (0.88-2.59) |
| . | 14 | 103074392 | C | CA | | intronic | *RCOR1* | 39 (6.96%) | 25 (6.54%) | 24 (2.77%) | 8.87E-03 | 1.83 (1.16-2.89) | 0.90 | 1.07 (0.62-1.88) |
| rs35690069 | 14 | 103086725 | A | AG | DELINS | intronic | *RCOR1* | 377 (67.32%) | 210 (55.26%) | 538 (62.12%) | 3.28E-03 | 1.37 (1.11-1.70) | 2.08E-04 | 1.67 (1.26-2.20) |
| rs11447626 | 14 | 103088519 | T | TA | DELINS | intronic | *RCOR1* | 380 (67.86%) | 211 (55.24%) | 540 (62.36%) | 1.92E-03 | 1.40 (1.13-1.74) | 9.17E-05 | 1.71 (1.30-2.26) |
| rs149643159 | 14 | 103093592 | G | GT | DELINS | intronic | *RCOR1* | 49 (8.75%) | 21 (5.50%) | 53 (6.12%) | 0.03 | 1.52 (1.02-2.25) | 0.08 | 1.65 (0.95-2.95) |
| rs3069237 | 14 | 103108580 | C | CTTTG | DELINS | intronic | *RCOR1* | 380 (67.86%) | 211 (55.24%) | 543 (62.70%) | 2.70E-03 | 1.38 (1.12-1.72) | 9.17E-05 | 1.71 (1.30-2.26) |

| | | | | | | | | | | | | | | |
|---|---|---|---|---|---|---|---|---|---|---|---|---|---|---|
| rs3069244 | 14 | 103118823 | G | GTT | DELINS | intronic | *RCOR1* | 180 (32.14%) | 170 (44.50%) | 323 (37.30%) | 2.72E-03 | 0.73 (0.58-0.90) | 1.55E-04 | 0.59 (0.45-0.78) |
| rs10656228 | 14 | 103123488 | T | TAC | DELINS | intronic | *RCOR1* | 180 (32.14%) | 169 (44.24%) | 325 (37.53%) | 2.70E-03 | 0.72 (0.58-0.90) | 2.03E-04 | 0.60 (0.45-0.79) |
| rs111794625 | 14 | 103151399 | T | TTG | INS | intronic | *RCOR1* | 167 (29.82%) | 165 (43.19%) | 306 (35.50%) | 9.63E-04 | 0.70 (0.56-0.87) | 3.00E-05 | 0.56 (0.42-0.74) |

[a] The positions of indels are based on NCBI human genome reference sequence Build 37

**Appendix Table 11 RT-qPCR primers**

| Gene | Forward (5′ to 3′) | Reverse (5′ to 3′) |
|---|---|---|
| *GAPDH* | GCACCGTCAAGGCTGAGAAC | TGGTGAAGACGCCAGTGGA |
| *RCOR1* | AACTGGCAAGACGCAGTCAA | GTTGGGCAAATCAGCCAATGA |

1. Neogi T, Jansen TLTA, Dalbeth N, et al. 2015 gout classification criteria: an American College of Rheumatology/European League Against Rheumatism collaborative initiative. *Arthritis Rheum* 2015; **67**(10): 2557-68.
